# Associations between socioeconomic status and white matter microstructure in children: indirect effects via obesity and cognition

**DOI:** 10.1101/2023.02.09.23285150

**Authors:** Zhaolong Adrian Li, Yuqi Cai, Rita L. Taylor, Sarah A. Eisenstein, Deanna M. Barch, Scott Marek, Tamara Hershey

## Abstract

**Importance:** Both neighborhood and household socioeconomic disadvantage relate to negative health outcomes and altered brain structure in children. It is unclear whether such findings extend to white matter development, and via what mechanisms socioeconomic status (SES) influences the brain.

**Objective:** To test independent associations between neighborhood and household SES indicators and white matter microstructure in children, and examine whether body mass index and cognitive function (a proxy of environmental cognitive/sensory stimulation) may plausibly mediate these associations.

**Design:** This cross-sectional study used baseline data from the Adolescent Brain Cognitive Development (ABCD) Study, an ongoing 10-year cohort study tracking child health.

**Setting:** School-based recruitment at 21 U.S. sites.

**Participants:** Children aged 9 to 11 years and their parents/caregivers completed baseline assessments between October 1^st^, 2016 and October 31^st^, 2018. Data analysis was conducted from July to December 2022.

**Exposures:** Neighborhood disadvantage was derived from area deprivation indices at primary residence. Household SES indicators were total income and the highest parental education attainment.

**Main Outcomes and Measures:** Thirty-one major white matter tracts were segmented from diffusion-weighted images. The Restriction Spectrum Imaging (RSI) model was implemented to measure restricted normalized directional (RND; reflecting oriented myelin organization) and isotropic (RNI; reflecting glial/neuronal cell bodies) diffusion in each tract. Obesity-related measures were body mass index (BMI), BMI *z*-scores, and waist circumference, and cognitive performance was assessed using the NIH Toolbox Cognition Battery. Linear mixed-effects models tested the associations between SES indicators and scanner-harmonized RSI metrics. Structural equation models examined indirect effects of obesity and cognitive performance in the significant associations between SES and white mater microstructure summary principal components. Analyses were adjusted for age, sex, pubertal development stage, intracranial volume, and head motion.

**Results:** The analytical sample included 8842 children (4299 [48.6%] girls; mean age [SD], 9.9 [0.7] years). Greater neighborhood disadvantage and lower parental education were independently associated with lower RSI-RND in forceps major and corticospinal/pyramidal tracts, and had overlapping associations in the superior longitudinal fasciculus. Lower cognition scores and greater obesity-related measures partially accounted for these SES associations with RSI-RND. Lower household income was related to higher RSI-RNI in almost every tract, and greater neighborhood disadvantage had similar effects in primarily frontolimbic tracts. Lower parental education was uniquely linked to higher RSI-RNI in forceps major. Greater obesity-related measures partially accounted for these SES associations with RSI-RNI. Findings were robust in sensitivity analyses and mostly corroborated using traditional diffusion tensor imaging (DTI).

**Conclusions and Relevance:** These cross-sectional results demonstrate that both neighborhood and household contexts are relevant to white matter development in children, and suggest cognitive performance and obesity as possible pathways of influence. Interventions targeting obesity reduction and improving cognition from multiple socioeconomic angles may ameliorate brain health in low-SES children.

**Key Points:** *Question:* Are neighborhood and household socioeconomic levels associated with children’s brain white matter microstructure, and if so, do obesity and cognitive performance (reflecting environmental stimulation) mediate the associations?

*Findings:* In a cohort of 8842 children, higher neighborhood disadvantage, lower household income, and lower parental education had independent and overlapping associations with lower restricted directional diffusion and greater restricted isotropic diffusion in white matter. Greater body mass index and poorer cognitive performance partially mediated these associations.

*Meaning:* Both neighborhood and household poverty may contribute to altered white matter development in children. These effects may be partially explained by obesity incidence and poorer cognitive performance.

## 1. Introduction

Socioeconomic disadvantage (e.g., poverty) during early life, as experienced by nearly 11.8 million U.S. children in 2021^1^, relates robustly to poor physical and mental health that can extend into adulthood^2,3^. Impoverished environments have been shown to affect brain structure and function, particularly during development^4–6^. Further, there is evidence that early neurodevelopmental alterations may mediate the links between low socioeconomic status (SES) and both current and future negative health outcomes^7–10^. Such evidence implicates the brain as a potential target for intervention.

In children, low SES has been associated with lower cortical volumes, surface area, and thickness, as well as lower hippocampal volume^11^. Lower frontal gyri volumes and surface area further related to poorer cognition and more externalizing symptoms^9,12–14^. In contrast to the growing literature documenting SES-related differences in gray matter, studies on white matter remain scarce. While support exists for a link between low SES and compromised white matter microstructure in children^15–20^, findings diverge on which tracts are implicated, likely as a result of limited sample sizes and different SES measurements^8,21^. White matter tracts, which are primarily myelin-ensheathed axonal bundles connecting distal gray matter regions, are integral for long-range information processing^22^. For example, lower integrity in the superior longitudinal fasciculus and cingulum has been respectively associated with poorer working memory and greater psychopathology in youth^23,24^. During childhood and adolescence, white matter undergoes rapid myelination and microstructural organization and continues to mature well into young adulthood^4,25^. Such a protracted developmental window could allow long-lasting influences of low SES on white matter and, due to its functional relevance, warrants in-depth investigation.

A key question is how SES influences brain development. Proximal factors such as nutrition and environmental enrichment, amongst others, are theorized to be influenced by SES and thus could impact brain health^7,8,26,27^. Studies have recently demonstrated that unhealthy weight and poorer cognitive performance mediate associations between low SES and altered brain volumes and functional connectivity^13,28,29^. These studies used obesity measures as a proxy for diet and exercise^13^, and cognitive performance as a proxy for the social, sensory, and cognitive stimulation in the environment^28,29^. Furthermore, there may be multiple components of SES that influence the brain, potentially through different pathways. Recent findings suggest that SES indicators at both neighborhood-level (e.g., area deprivation index) and household-level (e.g., income, parental education), though correlated, do independently relate to children’s brain morphology and functional networks^12,13,29–33^. In terms of SES, families having the same income level may have access to different neighborhoods due to differences in local cost of living and/or structural racism barriers^8^. Proximally, neighborhood SES could reflect environmental (e.g., noise, pollution, crime) and/or social influences (e.g., interactions with teachers and peers), whereas household SES may reflect home characteristics such as material access and parenting practices^7,8,27^. However, it remains unknown how neighborhood and household SES relate to white matter development, and via what mechanisms. Identifying potential mediating factors within this framework may reveal important intervention targets to promote brain health in disadvantaged children.

Leveraging the multi-shell restriction spectrum imaging (RSI) model, we address these knowledge gaps by interrogating associations between SES and white matter microstructure in a large cross-sectional cohort of children aged 9-11 years from the Adolescent Brain Cognitive Development (ABCD) Study. RSI goes beyond traditional diffusion tensor imaging (DTI) by distinguishing restricted (originating intracellularly) from hindered (originating extracellularly) water diffusion within image voxels, thus modeling for greater tissue complexity^34–37^. Specifically, white matter microstructure was assessed using restricted normalized directional (RSI-RND), which models cylindrical intracellular water diffusion and purportedly reflects oriented axons/dendrites; and restricted normalized isotropic (RSI-RNI), which models spherical intracellular water diffusion and purportedly reflects glial/neuronal cell bodies^36–40^ (see **Figure 1**, adapted from Burnor et al. (2021), *JAMA Netw Open*). Given that socioeconomic deprivation has been linked to impaired myelination and heightened microglial activity^6,20^, we expected lower SES to be associated with decreased white matter RSI-RND and increased RSI-RNI. We further expected that greater neighborhood disadvantage and lower household SES (i.e., income, parental education) would *independently* relate to white matter microstructure, paralleling the independent associations that have been seen with gray matter and function. Given known links between obesity, cognition, and white matter microstructure in children^41,42^, we hypothesized that obesity measures and cognition would have indirect effects, *independent for each SES indicator*, on the associations between lower SES and altered white matter microstructure, thereby boosting the plausibility of these proximal factors being causal mediators and potentially motivating their management from both community and household angles.

**Figure 1.**
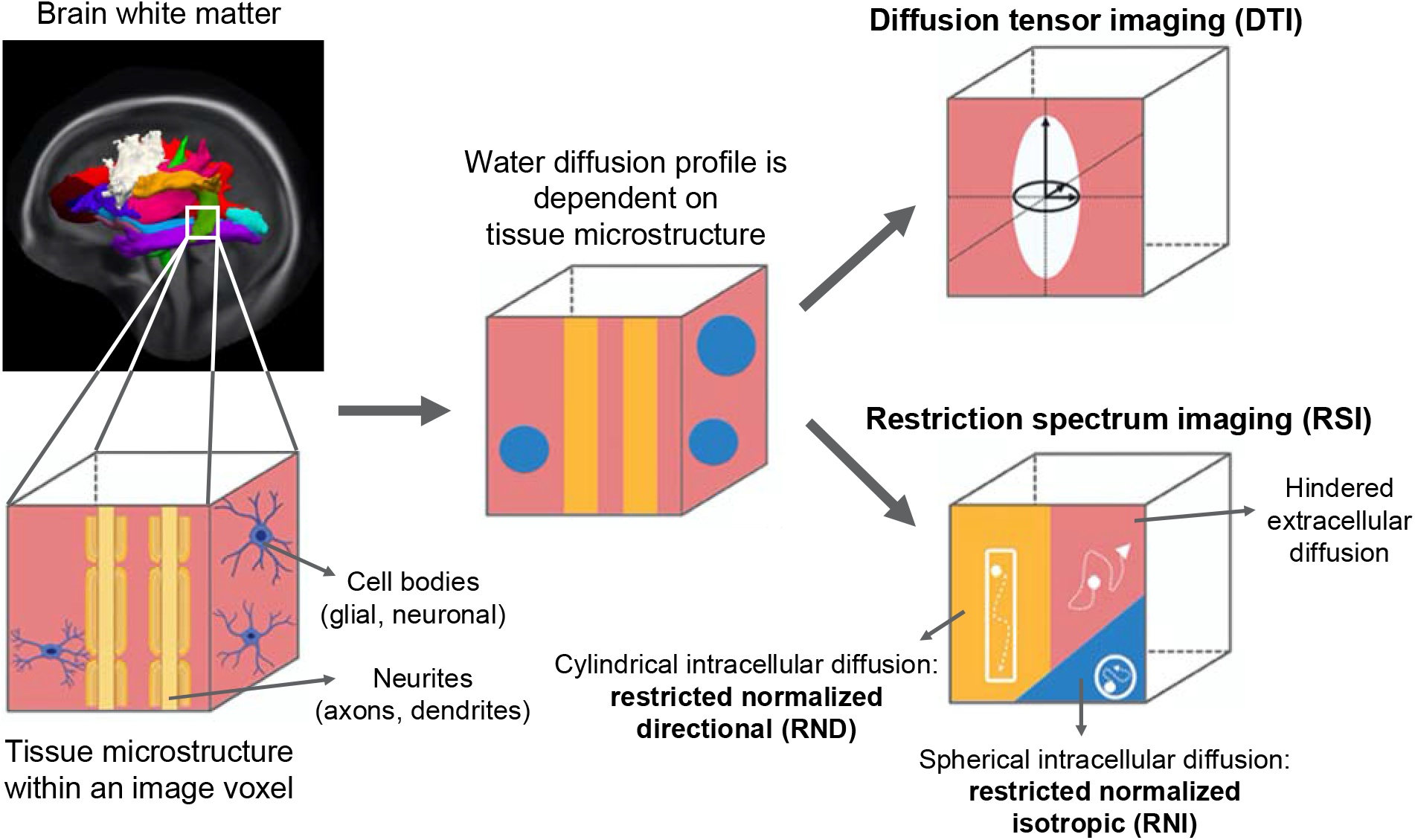
Schematic of RSI and DTI models. *Note*. Water diffusion patterns in white matter, measured in diffusion-weighted magnetic resonance imaging, are shaped by the complex tissue microstructure that may constitute neurites (axons/dendrites), cell bodies (glial/neuronal), and extracellular space. Water diffusion occurs at different scales, subject to the barriers created by these processes. Traditional diffusion tensor imaging (DTI) characterizes the anisotropy of water diffusion in a single 3-dimensional model, and gives metrics of fractional anisotropy (FA) and mean diffusivity (MD). Restriction spectrum imaging (RSI) however models intracellular/restricted diffusion (on a scale less than about 10 μm) and extracellular/hindered diffusion (on a scale greater than about 10 μm) separately. Within the intracellular/restricted compartment, it further separates cylindrical/directional diffusion (RND) from spherical/isotropic diffusion (RNI). These two metrics were normalized to the total diffusion signal, so that greater RSI-RND and RNI would respectively reflect greater relative signal contributions from directional and isotropic diffusion within a voxel. RSI offers greater insight to specific biological processes relative to DTI. In this study, RSI-RND and RNI were the primary white matter microstructure assessments, and DTI-FA and MD were estimated for reference. This image was adapted from Burnor et al. (2021). *JAMA Netw Open*. Under CC-BY license.

## 2. Methods

### 2.1. Participants

Participants were from the ABCD Study, a ten-year cohort study tracking child brain development across 21 U.S. sites representing national demographics^43,44^. Participants receive annual physical, environmental, and behavioral assessments, as well as neuroimaging and cognitive testing every two years^37,45–47^. Institutional review boards at study sites approved procedures; parents/caregivers provided written consent and children gave verbal assent. Our study employed ABCD Study baseline data (release 4.0; collected between October 1^st^, 2016 and October 31^st^, 2018). In addition to the standard ABCD Study inclusion/exclusion criteria^43^, we excluded participants with 1) missing age, sex, or anthropometric data; 2) T1 or diffusion-weighted magnetic resonance imaging (MRI) scans that failed quality control or had clinically significant incidental findings^37^; and 3) history of severe neurological or psychiatric conditions (**eMethods** in the **Supplement**). We also excluded participants with history of diabetes or eating disorders that may confound with obesity-related neurobiological findings^48^. These additional criteria led to a sample of 8842 children from 11875 total. Description of participant selection is shown stepwise in **eFigure 1** in the **Supplement**. This report follows the Strengthening the Reporting of Observational Studies in Epidemiology (STROBE) guidelines.

### 2.2. Measures

Participant age, sex, race/ethnicity, and pubertal development stage (PDS)^49^ were collected; other measures are defined below (see **eTable 1** in the **Supplement** for corresponding ABCD Study instrument names).

#### 2.2.1. Neighborhood SES

The participant’s primary home address was geocoded into a census tract, from which the American Community Survey (2011-2015) Area Deprivation Index (ADI) estimates were extracted^50^. An exploratory factor analysis created a latent neighborhood disadvantage score that included 10 of all 17 ADI values (loadings ≥ 0.63; **eTable 2** in the **Supplement**), consistent with previous reports^12,29^. ADI values that loaded less strongly generally reflected outdated (e.g., percentage of households without telephones) or market-dependent (e.g., median mortgage/rent) instead of contemporary, stable disadvantage and were excluded^12,29,30^. In the current study, *higher* neighborhood disadvantage scores would suggest *lower* education, income, property ownership, and *higher* unemployment, poverty, income disparity, and percentage of single-parent households, all at the community level.

#### 2.2.2. Household SES

Household income was the combined annual income from all family members. As it was assessed in ordinal ranges, we used a reported procedure^30^ that divided the midpoint of each income bracket by $10000, creating a continuous variable. Parental education level was the highest education attainment amongst parents/caregivers, recoded into years of schooling estimated per U.S. convention following previous studies^31,33^ (**eMethods** in the **Supplement**).

#### 2.2.3. Neuroimaging

Participants were scanned following standardized protocols across 3T scanner platforms (Siemens *Prisma* and *Prisma Fit*; GE *Discovery MR750*; and Philips *Achieva dStream* and *Ingenia*) at study sites^47^. T1-weighted structural images and multi-shell diffusion-weighted images (DWIs) were collected (see **eMethods** in the **Supplement** for specifications). Scans were processed centrally at the ABCD Data Analysis, Informatics, & Resource Center^37^. DWIs were corrected for eddy current, head motion, and susceptibility-induced distortions during preprocessing. Participant mean head motion during scanning was inferred from average DWI framewise displacement and was covaried in statistical analyses to partial out residual motion effects. Intracranial volumes (ICVs) were estimated from T1-weighted images. RSI was fitted to DWI-derived fiber orientation density functions to model RSI-RND and RNI, which were each normalized by total diffusion signal and served as the primary assessment of white matter microstructure in the present study. To evaluate the convergent validity of our novel RSI-derived results, we repeated analyses with DTI fractional anisotropy (FA) and mean diffusivity (MD). Previous work suggests a positive correspondence between RSI-RND and DTI-FA, both reflecting anisotropic diffusion, in development^36^, epilepsy^51^, and Alzheimer’s disease^34^. Low SES has been consistently associated with lower DTI-FA^15–19^, but findings have been mixed for DTI-MD. Thus, we expected similar pattern of results between RSI-RND and DTI-FA. Major white matter tracts were delineated by matching prior probabilities and diffusion orientations of fibers from AtlasTrack to individual DWIs^37,52^. RSI and DTI metrics were extracted from 31 tracts^52^, including the corpus callosum (CC) (with forceps major (Fmaj) and minor (Fmin) subregions) and bilateral fornix (Fx), cingulate cingulum (CgC), parahippocampal cingulum (CgH), corticospinal/pyramidal tract (CST), anterior thalamic radiations (ATR), uncinate fasciculus (Unc), inferior longitudinal fasciculus (ILF), inferior frontal-occipital fasciculus (IFOF), superior longitudinal fasciculus (SLF) (with temporal (tSLF; i.e., arcuate fasciculus) and parietal (pSLF) subregions), superior-corticostriatal tract (SCS), striatal to inferior-frontal cortical tract (SIFC), and inferior-frontal to superior-frontal cortical tract (IFSFC). Visualizations of individual tracts are shown in **eFigure 2** in the **Supplement**.

#### 2.2.4. Obesity-related measures

Participant waist circumference, height, and weight were each averaged across up to three measurements. Body mass index (BMI) was calculated (weight_(lbs)_/height_(in)_^2^ × 703). Age and sex-corrected BMI *z*-scores were computed using the 2000 CDC growth charts^53^. We used different obesity-related measures to address their varied accuracy in reflecting adiposity in children^54,55^.

#### 2.2.5. Cognitive performance

We assessed cognitive performance using the age-corrected total cognition score from the NIH Toolbox Cognition Battery, summarized from seven tasks that probed executive functioning, memory, language abilities, and processing speed^46^ (see detailed description in **eMethods** in the **Supplement**). Follow-up analyses using individual task and crystallized and fluid cognition composite scores were conducted to explore any cognitive domain-specific effects.

### 2.3. Statistical analyses

All analyses were performed in R version 4.2.1 (R Project for Statistical Computing).

#### 2.3.1. Handling of outliers and missing data

Outliers ± 4 SD away from the mean were removed. As the missingness in SES indicators seemed related to demographics (**eFigure 3** in the **Supplement**), we imputed missing data 50 times using the “mice” package instead of excluding them^56^. Neuroimaging and cognitive variables, being primary outcomes, were not imputed (**eTable 3** in the **Supplement**). Imputation did not bias data distribution (**eTable 4** in the **Supplement**).

#### 2.3.2. Harmonization of neuroimaging data

Technical differences between scanners accounted for substantial variance in RSI and DTI metrics (*R*^2^’s = 0.06 to 0.55, *p*’s < 0.001) and could contaminate statistical inference^57^. Because SES indicators were unevenly distributed across scanner sites (*R*^2^’s ≥ 0.09, *p*’s < 0.001), the convention of analyzing each scanner separately in multilevel modeling could underestimate meaningful SES-related variance. Instead, we harmonized RSI and DTI data using the batch-adjustment algorithm ComBat^58^, reducing scanner effects (post-harmonization *R*^2^’s ≤ 0.002 (> 30-fold reduction compared to pre-harmonization), *p*’s = 0.007 to 0.99) while retaining inherent associations between neuroimaging metrics and age, sex, race/ethnicity, PDS, and SES (**eTable 5** in the **Supplement**).

#### 2.3.3. Associations with SES

Associations between SES and white matter microstructure were assessed using linear mixed-effects models (“lme4” package^59^) in which neighborhood disadvantage, household income, and parental education were independent variables (IVs) included simultaneously and RSI or DTI metrics were dependent variables (DVs). Age, sex, PDS, ICV, and mean head motion were covaried due to potential confounding^36,39,60^, and family was modeled as a random effect. Associations between SES, obesity-related measures, and cognition were examined using the same procedure but without covarying for ICV and mean head motion. Because SES was highly entangled with race/ethnicity (**eTable 6** in the **Supplement**), we did not adjust for race/ethnicity in models in order to preserve SES-related variance^61^. In follow-up analyses, covarying for race/ethnicity led to more restricted findings (**eTable 10** in the **Supplement**). All models were checked for normality of residuals, homoscedasticity, and low multicollinearity (variance inflation factors were ≤ 1.89). Estimates were standardized β’s with 95% confidence intervals (CIs) pooled across imputed datasets. Multiple comparison correction was performed within each group of models and by each SES indicator using false discovery rate (FDR) at two-tailed *p*_FDR_ ≤ 0.05. As missing neuroimaging and cognition data were not imputed, sample sizes varied across models and are reported in **eTable 7** in the **Supplement**.

#### 2.3.4. Sensitivity analyses

Associations between SES and white matter microstructure were further examined in subsamples that censored potential neuroimaging confounds: 1) participants with mean head motion ≤ 2.5 mm; 2) participants without adverse childhood experiences (e.g., trauma/abuse); 3) participants without common psychiatric diagnoses, including attention-deficit/hyperactivity disorder, depression, bipolar disorder, anxiety, and phobias; and 4) participants with full-term birth.

#### 2.3.5. Testing for indirect effects

We estimated the indirect effects of SES on white matter microstructure through obesity-related measures and cognition using the “lavaan” package^62^. As we employed cross-sectional data, such analyses can establish the plausibility of factors as potential causal mediators that could be confirmed in future longitudinal studies. To reduce data dimensionality, principal component analyses (PCA) were applied to RSI or DTI metrics in tracts that demonstrated significant associations with SES. Structural equation models specified each of the SES indicators as IV, obesity-related measures or cognitive performance as mediator, and white matter microstructure principal components (PCs) as DVs. Covariates included the other SES indicators, age, sex, PDS, ICV, and mean head motion. As multilevel modeling was not feasible in “lavaan”, findings were confirmed in a random subsample of unrelated participants that eliminated possible family confounds (**eTable 20** in the **Supplement**). Lastly, building upon previous research^12,13,33^, we studied additional models with white matter microstructure PCs as mediators and cognition as DV. Standardized estimates were computed with standard errors (SEs) and 95% CIs (from 20000 Monte Carlo simulations). Statistical significance was at two-tailed *p*_FDR_ ≤ 0.05 corrected across all tested models.

## 3. Results

This study included 8842 children (4299 [48.6%] girls; mean age [SD], 9.9 [0.7] years); sample characteristics are detailed in **Table 1**. SES indicators were moderately correlated with each other (**eFigure 4** in the **Supplement**).

**Table 1.**
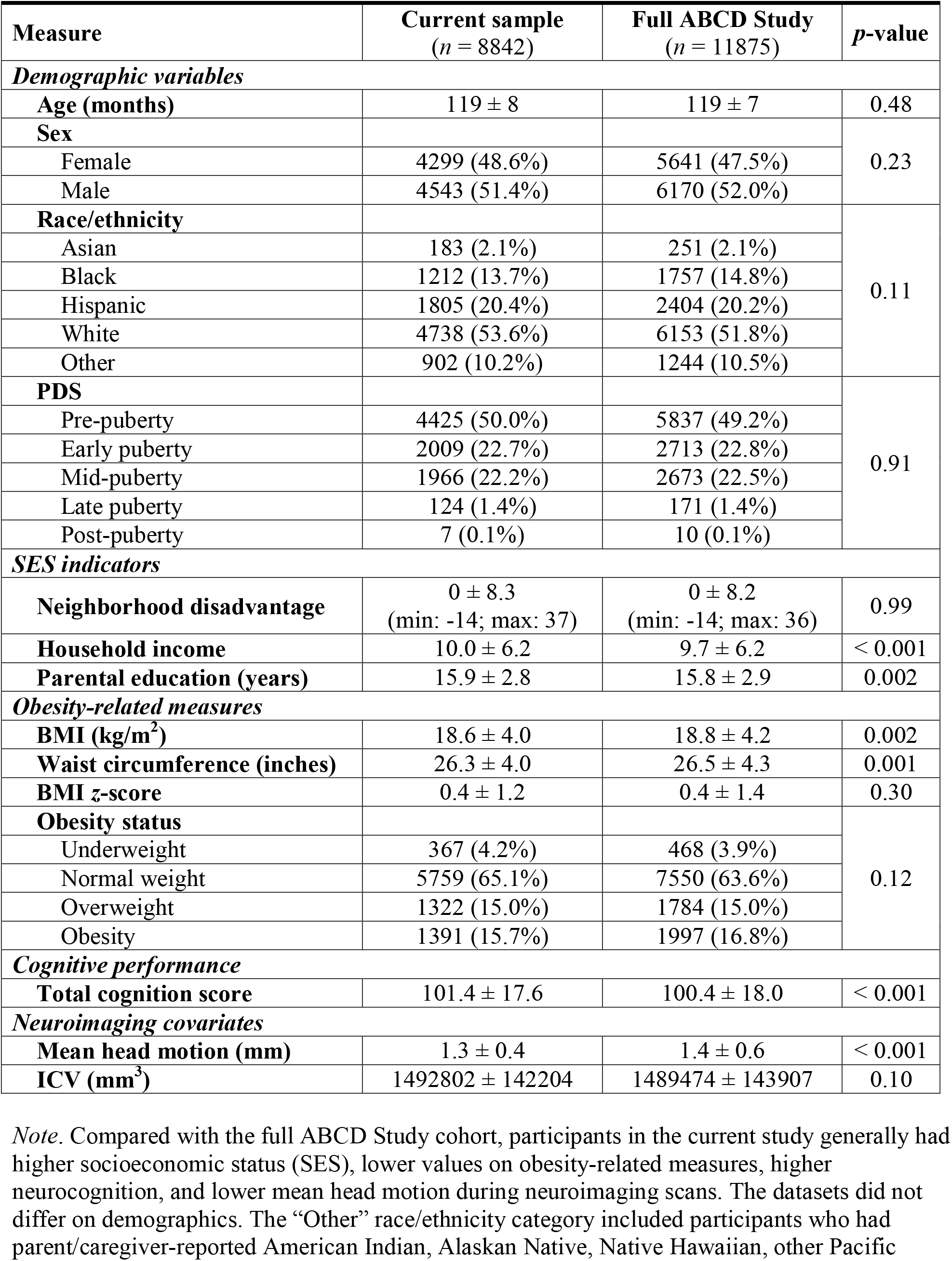
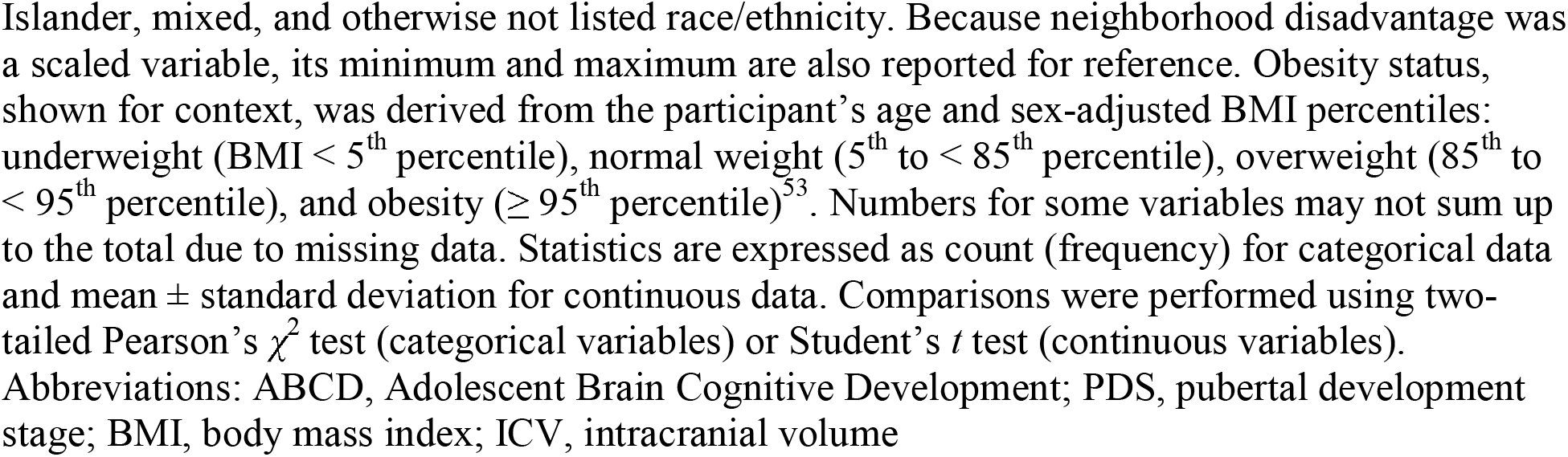
Sample characteristics.

| Measure | Current sample<br>(*n* = 8842) | Full ABCD Study<br>(*n* = 11875) | *p*-value |
| --- | --- | --- | --- |
| *Demographic variables* |  |  |  |
| Age (months) | 119 ± 8 | 119 ± 7 | 0.48 |
| Sex |  |  | 0.23 |
| Female | 4299 (48.6%) | 5641 (47.5%) |  |
| Male | 4543 (51.4%) | 6170 (52.0%) |  |
| Race/ethnicity |  |  | 0.11 |
| Asian | 183 (2.1%) | 251 (2.1%) |  |
| Black | 1212 (13.7%) | 1757 (14.8%) |  |
| Hispanic | 1805 (20.4%) | 2404 (20.2%) |  |
| White | 4738 (53.6%) | 6153 (51.8%) |  |
| Other | 902 (10.2%) | 1244 (10.5%) |  |
| PDS |  |  | 0.91 |
| Pre-puberty | 4425 (50.0%) | 5837 (49.2%) |  |
| Early puberty | 2009 (22.7%) | 2713 (22.8%) |  |
| Mid-puberty | 1966 (22.2%) | 2673 (22.5%) |  |
| Late puberty | 124 (1.4%) | 171 (1.4%) |  |
| Post-puberty | 7 (0.1%) | 10 (0.1%) |  |
| *SES indicators* |  |  |  |
| Neighborhood disadvantage | 0 ± 8.3<br>(min: -14; max: 37) | 0 ± 8.2<br>(min: -14; max: 36) | 0.99 |
| Household income | 10.0 ± 6.2 | 9.7 ± 6.2 | < 0.001 |
| Parental education (years) | 15.9 ± 2.8 | 15.8 ± 2.9 | 0.002 |
| *Obesity-related measures* |  |  |  |
| BMI (kg/m<sup>2</sup>) | 18.6 ± 4.0 | 18.8 ± 4.2 | 0.002 |
| Waist circumference (inches) | 26.3 ± 4.0 | 26.5 ± 4.3 | 0.001 |
| BMI z-score | 0.4 ± 1.2 | 0.4 ± 1.4 | 0.30 |
| Obesity status |  |  | 0.12 |
| Underweight | 367 (4.2%) | 468 (3.9%) |  |
| Normal weight | 5759 (65.1%) | 7550 (63.6%) |  |
| Overweight | 1322 (15.0%) | 1784 (15.0%) |  |
| Obesity | 1391 (15.7%) | 1997 (16.8%) |  |
| *Cognitive performance* |  |  |  |
| Total cognition score | 101.4 ± 17.6 | 100.4 ± 18.0 | < 0.001 |
| *Neuroimaging covariates* |  |  |  |
| Mean head motion (mm) | 1.3 ± 0.4 | 1.4 ± 0.6 | < 0.001 |
| ICV (mm<sup>3</sup>) | 1492802 ± 142204 | 1489474 ± 143907 | 0.10 |

*Note.* Compared with the full ABCD Study cohort, participants in the current study generally had higher socioeconomic status (SES), lower values on obesity-related measures, higher neurocognition, and lower mean head motion during neuroimaging scans. The datasets did not differ on demographics. The “Other” race/ethnicity category included participants who had parent/caregiver-reported American Indian, Alaskan Native, Native Hawaiian, other Pacific
Islander, mixed, and otherwise not listed race/ethnicity. Because neighborhood disadvantage was a scaled variable, its minimum and maximum are also reported for reference. Obesity status, shown for context, was derived from the participant's age and sex-adjusted BMI percentiles: underweight (BMI < 5<sup>th</sup> percentile), normal weight (5<sup>th</sup> to < 85<sup>th</sup> percentile), overweight (85<sup>th</sup> to < 95<sup>th</sup> percentile), and obesity ( ≥ 95<sup>th</sup> percentile)<sup>53</sup>. Numbers for some variables may not sum up to the total due to missing data. Statistics are expressed as count (frequency) for categorical data and mean ± standard deviation for continuous data. Comparisons were performed using two-tailed Pearson's $\chi^2$ test (categorical variables) or Student's *t* test (continuous variables). Abbreviations: ABCD, Adolescent Brain Cognitive Development; PDS, pubertal development stage; BMI, body mass index; ICV, intracranial volume

### 3.1. Associations between SES and white matter microstructure

Higher SES was associated with greater RSI-RND, lower RSI-RNI, and greater DTI-FA in 9, 29, and 10 out of 31 tracts, respectively. Full statistics are shown in **eTable 8** in the **Supplement**; key results are noted below. Findings largely survived sensitivity analyses (**eTable 9** in the **Supplement**).

#### 3.1.1. Associations with RSI-RND

Both neighborhood disadvantage and parental education had independent associations with white matter RSI-RND, overlapping in the SLF (**Figure 2**). Higher neighborhood disadvantage was associated with lower RSI-RND in Fmaj (β = -0.040; 95% CI: -0.067 to -0.013; *p*_FDR_ = 0.03) and left SLF (β = -0.055; 95% CI: -0.081 to -0.028; *p*_FDR_ = 0.001). Higher parental education was associated with greater RSI-RND in bilateral CST (e.g., right β = 0.042; 95% CI: 0.015 to 0.069; *p*_FDR_ = 0.01) and SLF (e.g., right β = 0.053; 95% CI: 0.025 to 0.080; *p*_FDR_ = 0.002). No independent associations were observed for household income.

**Figure 2.**
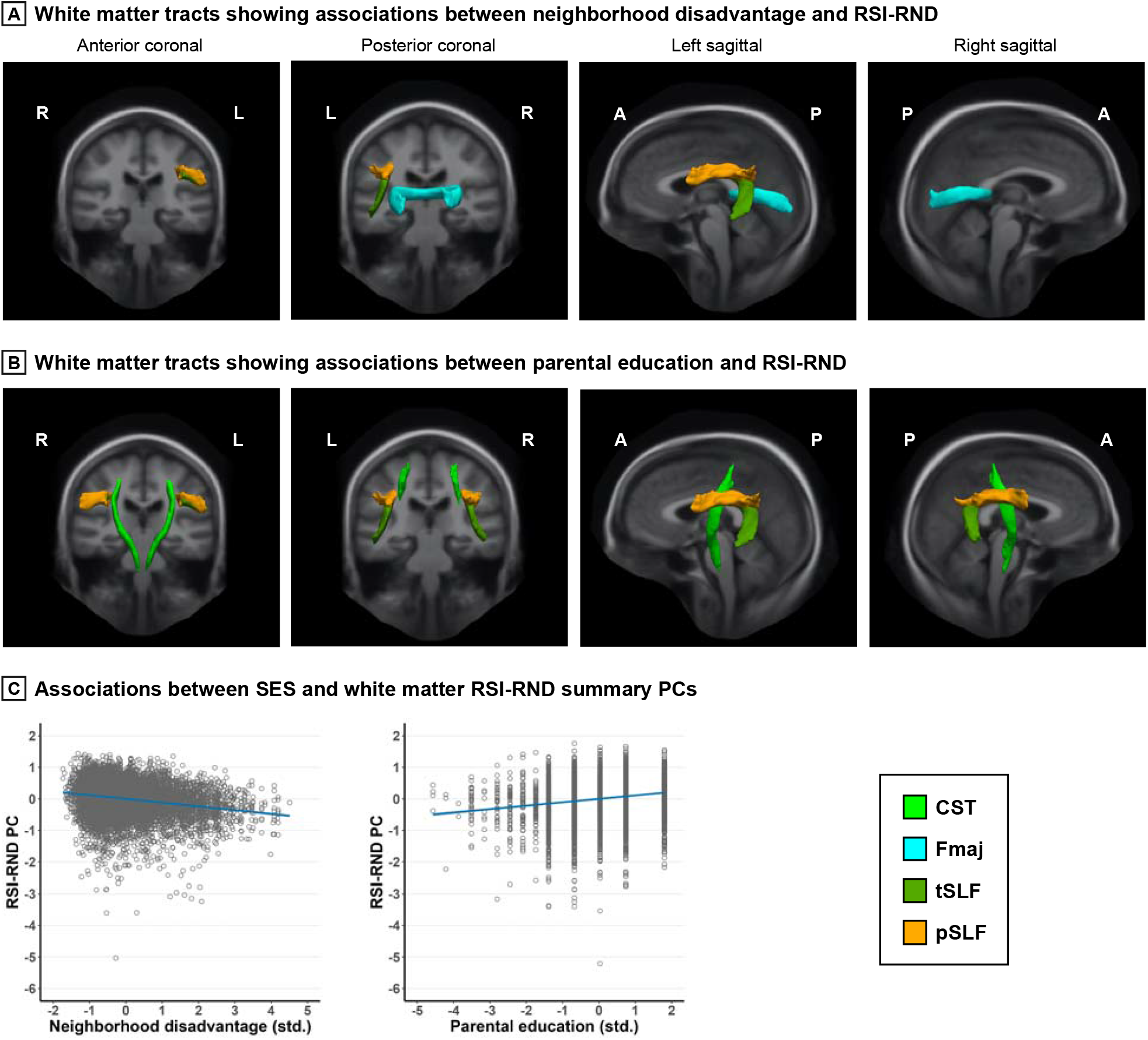
Associations between SES and white matter RSI-RND. *Note*. Greater neighborhood disadvantage and higher parental education were independently associated with lower and greater RSI-RND in specific white matter tracts, respectively. Tracts were visualized using the AtlasTrack atlas. The principal components (PCs) summarized RSI-RND in the relevant tracts, as associations did not differ qualitatively between tracts. In scatterplots, linear regression lines were adjusted for covariates and are flanked by shaded 95% confidence interval. Data points were standardized (std.) residuals extracted from a randomly selected imputed dataset as reference. Covariates included age, sex, pubertal development stage, intracranial volume, and head motion, and family was the random effect. Detailed statistics are shown in **eTable 8** and **12** in the **Supplement**. RSI, restriction spectrum imaging; RND, restricted normalized directional; R, right; L, left; A, anterior; P, posterior; SES, socioeconomic status; CST, corticospinal/pyramidal tract; Fmaj, forceps major; SLF, superior longitudinal fasciculus, including temporal (t) and parietal (p) subregions.

#### 3.1.2. Associations with RSI-RNI

All three SES indicators were independently associated with white matter RSI-RNI, with some spatial overlap of effects between household income and neighborhood disadvantage (**Figure 3**). Higher household income was associated with lower RSI-RNI in almost every tract (β’s = -0.062 to -0.031; *p*_FDR_’s = 0.002 to 0.05), except for bilateral Fx, Fmaj, right tSLF, and right SCS. Higher neighborhood disadvantage was associated with greater RSI-RNI in bilateral Fx (e.g., right β = 0.046; 95% CI: 0.019 to 0.074; *p*_FDR_ = 0.01) and, overlapping with household income, in bilateral CgH (e.g., right β = 0.061; 95% CI: 0.034 to 0.088; *p*_FDR_ < 0.001), CST (e.g., right β = 0.037; 95% CI: 0.010 to 0.065; *p*_FDR_ = 0.03), and ATR (e.g., right β = 0.045; 95% CI: 0.018 to 0.072; *p*_FDR_ = 0.01). Lastly, higher parental education was uniquely associated with lower RSI-RNI in Fmaj (β = -0.048; 95% CI: -0.077 to -0.020; *p*_FDR_ = 0.03).

**Figure 3.**
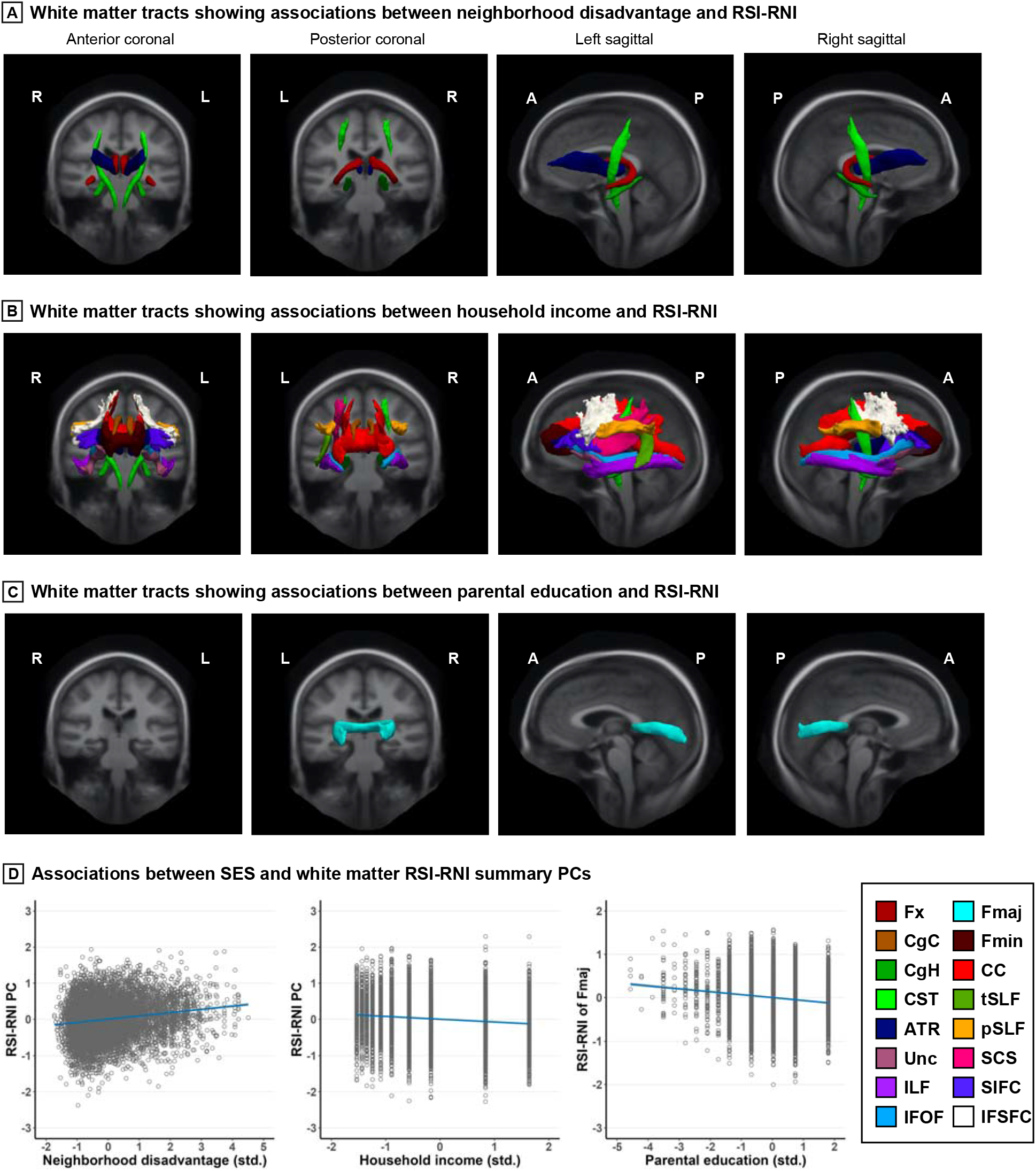
Associations between SES and white matter RSI-RNI. *Note*. Greater neighborhood disadvantage, higher household income, and higher parental education were independently associated with greater, lower, and lower RSI-RNI in white matter tracts, respectively. Widespread effects were seen for household income. Tracts were visualized using the AtlasTrack atlas. The principal components (PCs) summarized RSI-RNI in the relevant tracts, as associations did not differ qualitatively between tracts. In scatterplots, linear regression lines were adjusted for covariates and are flanked by shaded 95% confidence interval. Data points were standardized (std.) residuals extracted from a randomly selected imputed dataset as reference. Covariates included age, sex, pubertal development stage, intracranial volume, and head motion, and family was the random effect. Detailed statistics are shown in **eTable 8** and **12** in the **Supplement**. RSI, restriction spectrum imaging; RNI, restricted normalized isotropic; R, right; L, left; A, anterior; P, posterior; SES, socioeconomic status; Fx, fornix; CgC, cingulate cingulum; CgH, parahippocampal cingulum; CST, corticospinal/pyramidal tract; ATR, anterior thalamic radiations; Unc, uncinate fasciculus; ILF, inferior longitudinal fasciculus; IFOF, inferior frontal-occipital fasciculus; Fmaj, forceps major; Fmin, forceps minor; CC, corpus callosum; SLF, superior longitudinal fasciculus, including temporal (t) and parietal (p) subregions; SCS, superior-corticostriatal tract; SIFC, striatal to inferior-frontal cortical tract; IFSFC, inferior-frontal to superior-frontal cortical tract.

#### 3.1.3. Associations with DTI-FA and MD

Associations between SES and DTI-FA largely resembled those with RSI-RND. Higher neighborhood disadvantage had independent associations with lower RSI-RND in the left SLF, and higher parental education with greater RSI-RND in left Fx, bilateral CST, bilateral SLF, and left SCS (**eFigure 5** in the **Supplement**). Household income was not associated with DTI-FA, nor was any SES indicator with DTI-MD.

### 3.2. Analyses of indirect effects

In each PCA summarizing each independent SES indicator and RSI/DTI metric association, the first PC carried substantial loadings from all involved tracts, captured 58% to 95% of variance, and were related to the SES indicator as individual tracts did (**eTable 11** and **12** in the **Supplement**). These first PCs were thus used in indirect effects models. All models demonstrated good fit (**eTable 17** in the **Supplement**).

#### 3.2.1. Indirect effects via obesity-related measures

Lower SES indexed by all three SES indicators had independent associations with higher values of all obesity-related measures (**eTable 13** in the **Supplement**). Higher BMI had indirect effects on the associations between higher neighborhood disadvantage and lower RSI-RND PC and greater RSI-RNI PC; between lower household income and greater RSI-RNI PC; and between lower parental education and lower RSI-RND PC and greater RSI-RNI in Fmaj (**Figure 4A**). No effect was observed with DTI-FA PCs. Consistent results were seen with waist circumference and BMI *z*-scores. Full statistics are in **eTable 14** in the **Supplement**.

**Figure 4.**
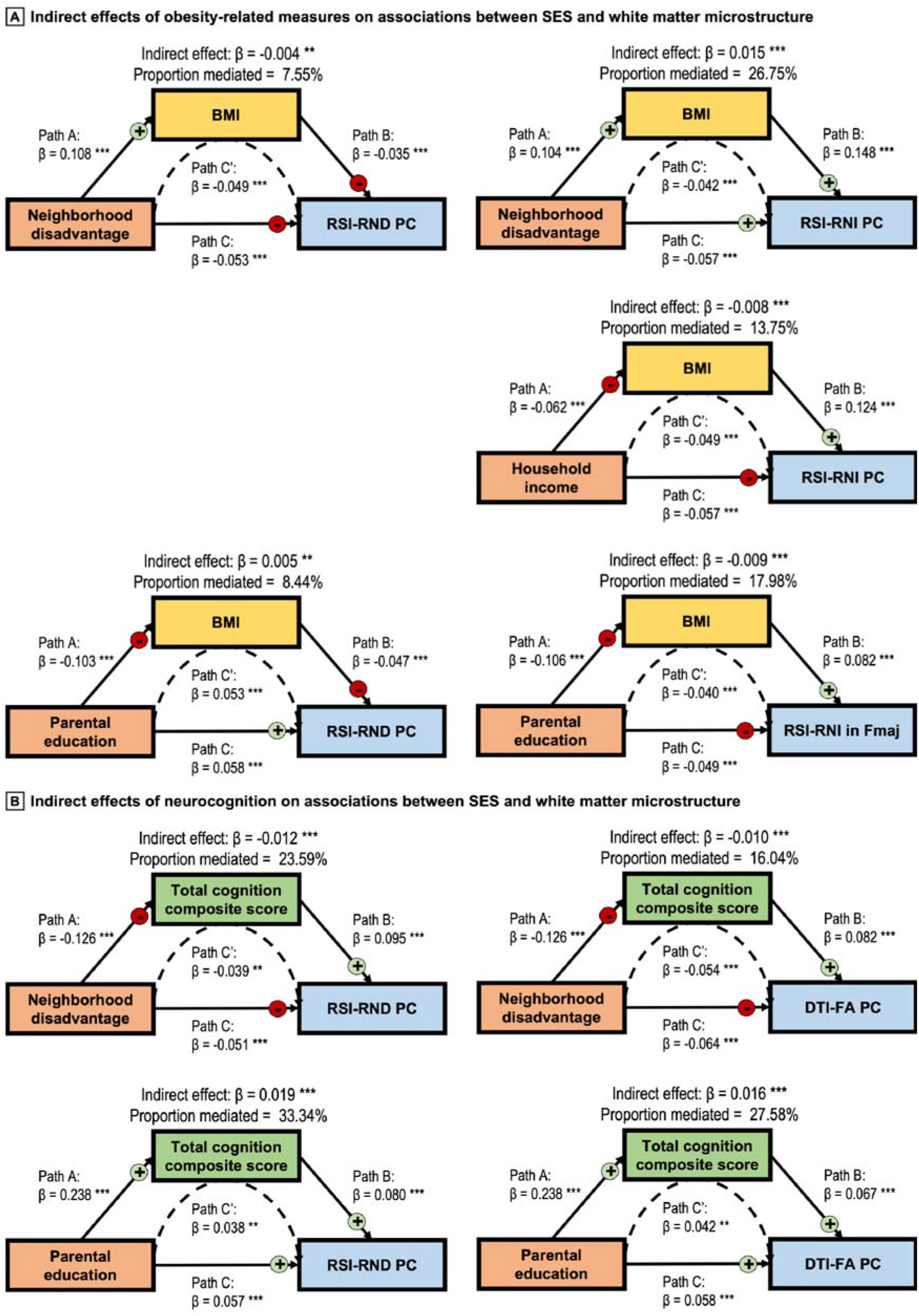
Indirect effects of obesity-related measures and neurocognition in the associations between SES and white matter microstructure. *Note*. Body mass index (BMI) and total cognition score constituted significant partial indirect effects between the associations between socioeconomic status (SES) indicators and white matter microstructure. For BMI, consistent results were seen with BMI *z*-scores and waist circumference. For cognitive performance, findings were mostly similar with individual task and composite scores. In each model, structural equation modeling covariates included age, sex, pubertal development stage, intracranial volume, and head motion, as well as the SES indicators that were not the independent variable (e.g., household income and parental education were covariates in models concerning neighborhood disadvantage). Detailed statistics are shown in **eTable 14** and **16** in the **Supplement**, and model fit indices are in **eTable 17**. **, *p* ≤ 0.01; ***, *p* ≤ 0.001. For indirect effects, *p*-values were corrected using false discovery rate (FDR). RSI, restriction spectrum imaging; RND, restricted normalized directional; RNI, restricted normalized isotropic; PC, principal component; Fmaj, forceps major; DTI, diffusion tensor imaging; FA, fractional anisotropy.

#### 3.2.2. Indirect effects via cognitive performance

Lower SES indexed by all three SES indicators was independently associated with lower total cognition score (**eTable 15** in the **Supplement**). Lower total cognition score had indirect effects on the associations between higher neighborhood disadvantage and lower RSI-RND and DTI-FA PCs. Conversely, higher total cognition score had indirect effects on the associations between higher parental education and greater RSI-RND and DTI-FA PCs (**Figure 4B**; full statistics in **eTable 16** in the **Supplement**). The total cognition score was not related to RSI-RNI PCs and was not tested for indirect effects in these models. Results were consistent across most analyses with composite and individual task scores, suggesting that our observed indirect effects related to general cognition rather than certain domains (**eTable 18** in the **Supplement**). In models where cognition was the DV, we observed indirect effects of greater RSI-RND PCs and greater DTI-FA PCs between higher SES and better cognition, again broadly across cognitive domains (**eTable 19** in the **Supplement**).

## 4. Discussion

In a large group of 9 to 11-year-old children, we found that greater neighborhood disadvantage and lower household SES related independently and robustly with lower RSI-RND, greater RSI-RNI, and lower DTI-FA in partially overlapping white matter regions. Furthermore, there was evidence for indirect effects of SES, whereby greater BMI and related anthropometrics, as well as poorer cognitive performance, partially explained associations between lower SES and altered white matter microstructure. Given the size and demographic diversity of our sample, these results suggest a potentially generalizable pattern that both neighborhood and household SES may be important for white matter development in children. Future research should investigate obesity and cognition as potential mechanistic mediators.

Higher SES was associated with greater RSI-RND in the SLF, Fmaj, and CST. As RSI-RND models anisotropic intracellular water diffusion^35^, these findings might reflect more oriented axonal/dendritic organization in these tracts^4,36,40^. Such interpretation parallels observations with DTI-FA, a traditional indicator of white matter integrity^36^, and is consistent with previous evidence of greater DTI-FA in the SLF^16,18,63,64^ and CST^18,64^ in children from high SES families. The integrity of these tracts have been implicated in cognition: the SLF is linked to language abilities, social cognition, and visuospatial attention^64,65^, and the Fmaj to gross memory functioning^66^. The CST, primarily a motor pathway, finetunes somatosensory-motor information integration necessary for fast processing speed^67^. Correspondingly, we found that higher cognition scores partially explained the links between higher SES and greater RSI-RND. This effect appeared general and not limited to particular cognitive domains, echoing the tracts’ wide functional relevance. This pathway might represent the level of cognitive stimulation a child receives, which has been shown to be higher in high SES families^26,27,68–70^. Nonetheless, our cross-sectional data precludes inference of directionality, and our models with white matter microstructure as the mediator supported the equal plausibility that SES may impact brain outcomes first and in turn contribute to cognition^12,13^. Cognitive performance also likely has both social (i.e., school, family) and neurobiological (i.e., brain-supported functioning) foundations that would position cognition as either a contributor to or consequence of brain development^26–28^, and future research may dissociate these effects using more specific measurements. Additionally, because morphometry in certain regions connected to the SLF (e.g., superior frontal-parietal cortices), CST (e.g., precentral gyrus), and Fmaj (e.g., occipital lobes) has similarly exhibited associations with SES and cognition in children^12–14,28,30,32,33^, our findings in white matter might reflect developmental interactions with gray matter instead of an isolated process^71^.

In comparison to the localized effects seen with RSI-RND, lower SES was associated with greater RSI-RNI in almost every white matter tract. Given that RSI-RNI measured spherical intracellular diffusion^35^, these widespread associations hint that SES might modulate glial/neuronal cell presence in a brain-wide fashion. Our observation that greater BMI and related anthropometrics partially accounted for these associations provides insights to some potential mechanisms. First, obesity induces systemic inflammation, which upregulates circulating pro-inflammatory molecules^72^ that may infiltrate brain tissue through a weakened blood-brain barrier^73,74^. As an immune response, the brain’s astrocytes and microglia undergo “reactive gliosis”, a process marked by their proliferation and enlargement as seen in rodent models of obesity^74–78^. In diffusion MRI models, such neuroinflammatory phenotype may manifest as increases in isotropic intracellular signals, a phenomenon observed in the striatum^39,42,79^ and recently hypothalamus and diffuse white matter tracts^42^ in childhood obesity. Thus, it is possible that low SES might be related to neuroinflammation via obesity incidence. Additionally, microglia dysregulation may cause myelin damage^80,81^, which could explain the observed association between obesity and RSI-RND. Second, because RSI-RNI increases with normative child development^36^, our findings of greater RSI-RNI, which were corrected for age, may represent accelerated white matter maturation with socioeconomic disadvantage. This interpretation echoes reports of higher gray matter-derived brain-predicted age amongst low-SES early adolescents^82–84^ and parallels the stress acceleration hypothesis, which proposes prioritized neuroadaptation to deprivation and adversity^85^. Indeed, the largest RSI-RNI effects were seen in frontolimbic pathways (Fx, CgH, ATR, and Unc) relevant for emotion processing, stress, and depression^86–88^. Notably, obesity has been linked to earlier pubertal onset^89^ and advanced brain aging^90,91^, and thus might embody a way SES impacts brain development. Future animal research comparing RSI to cellular/tissue imaging may help elucidate the exact neurobiological basis underlying our observed SES-related white matter microstructural changes.

The plausibility of obesity and cognitive performance being mediators linking SES to white matter microstructure, if confirmed in future longitudinal studies, may support their intervention to promote brain health in low-SES children. Weight loss has been shown to increase cortical volumes and white matter density^92,93^, attenuate neuroinflammation^94^, and normalize μ-opioid receptor availability^95^. Similar effects were also seen following low-fat dieting^96^ and aerobic training^97^. In the same vein, environments enriched in social and sensory stimuli are known to promote myelination in hypoxic rodents^68^ and aging adults^69^. Critically, because both neighborhood and household SES were independently associated with white matter microstructure, a distinction that continued in our indirect effects models, interventions may need to be conceptualized from multiple socioeconomic angles. At the neighborhood level, improved access to healthy food outlets and playgrounds may limit obesity amongst children living in low-SES areas^98,99^. Household factors such as buying power and health awareness, which might influence the child’s diet choice and participation in sports, are also important for maintaining healthy weight^99,100^. For cognition, sports and music programs in schools have been observed to improve academic achievement and cognitive performance in participating children^101,102^. At home, enriching parent-child interactions, such as reading and play, are known to benefit children’s long-term cognitive and overall well-being^103,104^. Crucially, many of these potential proximal mechanisms, whether through neighborhoods or households, are filtered through structural forces such as occupation, market, allocation of government resources, etc., and thus their management should be considered in terms of broad social policy. Further research is warranted to characterize the specific impact of these factors, together with other unexplored ones such as crime and pollution exposure^7,8,38^, on brain development.

### 4.1. Limitations

This study has limitations. First, the cross-sectional design precludes causal inference, and it is unknown whether our SES-related findings represent temporary or long-term developmental differences. Longitudinal investigations will be particularly meaningful, as our white matter observations, small in magnitude now, may accumulate over time^105^ and relate to future disease status^9,24^. Second, SES indicators, especially education attainment, also have genetic backgrounds^106,107^, and research is needed to dissociate the environmental from genetic contributions to white matter development. However, both genetics and environment influence white matter microstructure, with the former’s influence decreasing and the latter’s increasing from adolescence to adulthood^25,108^. Third, as many low-SES children had greater white matter integrity than their high-SES peers, our SES indicators were not predictors of white matter development at the individual level; rather, their small effects were seen in a large, population-based sample. Future studies are also encouraged to examine other SES-related facets, as well as their potential interactions^32^. Lastly, we only studied major white matter tracts due to the higher reliability of RSI and diffusion-weighted MRI models in these regions, and studies are needed to examine whether our findings extend to superficial/peri-cortical white matter.

## 5. Conclusions

To our knowledge, our study is the first large, multi-site investigation to find that both neighborhood and household socioeconomic adversity are associated with alterations in white matter microstructure in children. These independent associations could be partially explained by obesity and cognition, which may be targets for interventions from multiple socioeconomic angles to improve white matter health in disadvantaged children. Our findings join previous research on gray matter^12–14,28–32,109^ to highlight the complex pathways through which SES might influence brain development.

## Supporting information

All supplementary materials

## Data Availability

Data in this study were from the baseline measurements published in 2021 as part of the ABCD Curated Data Release 4.0, available at DOI: 10.15154/1523041. The ABCD data repository grows and may be modified as more data are collected and processed. This study is available in the NIMH Data Archive (NDA) at DOI: 10.15154/1528516.

## Abbreviations

ABCD: Adolescent Brain Cognitive Development
ADI: area deprivation index
ATR: anterior thalamic radiations
BMI: body mass index
CC: corpus callosum
CgC: cingulate cingulum
CgH: parahippocampal cingulum
CI: confidence interval
CST: corticospinal/pyramidal tract
DTI: diffusion tensor imaging
DV: dependent variable
DWI: diffusion-weighted image
FA: fractional anisotropy
FDR: false discovery rate
Fmaj: forceps major
Fmin: forceps minor
Fx: fornix
ICV: intracranial volumes
IFOF: inferior frontal-occipital fasciculus
IFSFC: inferior-frontal to superior-frontal cortical tract
ILF: inferior longitudinal fasciculus
IV: independent variable
MD: mean diffusivity
MRI: magnetic resonance imaging
PC: principal component
PCA: principal component analyses
PDS: pubertal development stage
pSLF: parietal superior longitudinal fasciculus
RND: restricted normalized directional
RNI: restricted normalized isotropic
RSI: restriction spectrum imaging
SCS: superior-corticostriatal tract
SD: standard deviation
SE: standard error
SES: socioeconomic status
SIFC: striatal to inferior-frontal cortical tract
SLF: superior longitudinal fasciculus
STROBE: Strengthening the Reporting of Observational Studies in Epidemiology
tSLF: temporal superior longitudinal fasciculus
Unc: uncinate fasciculus

## Author Contributions

Mr. Li had full access to all data in the present study and assumes responsibility for data integrity and accuracy of data analysis.

*Concept and design:* Li, Cai, Marek, Hershey.

*Acquisition, analysis, or interpretation of data:* All authors.

*Drafting of the manuscript:* Li.

*Critical revision of the manuscript for important intellectual content:* All authors.

*Statistical analysis:* Li, Marek, Hershey.

*Obtained funding:* Li, Marek, Hershey.

*Administrative, technical, or material support:* Marek, Hershey.

*Supervision:* Marek, Hershey.

## Disclaimer

The content is solely the responsibility of the authors and does not necessarily represent the official views of the National Institutes of Health or other funders.

## Additional Contributions

The authors thank Jerrel Rutlin, BS, Ashley Sanders, PhD, Amjad Samara, MD, and Xuanhe “Dahlia” Qi, BA for the meaningful discussions.

