## Supplementary material for "Associations between socioeconomic status and white matter microstructure in children: indirect effects via obesity and cognition": All supplementary materials

**Table of content**

| **Page** | **Designation** | **Description** |
| --- | --- | --- |
| 2 | **eMethods.** | Participant selection; derivation of SES indicators; neuroimaging specifications; cognitive testing |
| 3 | **eFigure 1.** | Flowchart of participant selection |
| 4 | **eTable 1.** | Lookup list for relevant ABCD Study instrument names |
| 5 | **eTable 2.** | Factor loadings of ADI metrics |
| 6 | **eFigure 2.** | Isosurface renderings of white matter tracts |
| 7-8 | **eFigure 3.** | Relationships between missingness of key variables |
| 9 | **eTable 3.** | Missing data and roles of variables in imputation |
| 10 | **eTable 4.** | Distribution of imputed variables before and after imputation of missing data |
| 11 | **eTable 5.** | Pre- and post-harmonization associations between preserved variables and RSI and DTI metrics |
| 12 | **eTable 6.** | Associations between SES and race/ethnicity |
| 13 | **eTable 7.** | Sample sizes for models testing associations between SES, RSI and DTI metrics, and neurocognition |
| 14 | **eFigure 4.** | Correlations between SES indicators |
| 15-19 | **eTable 8.** | Associations between SES and white matter microstructure |
| 20-26 | **eTable 9.** | Sensitivity analyses on associations between SES and white matter microstructure |
| 27 | **eFigure 5.** | Associations between SES and white matter DTI-FA |
| 28-32 | **eTable 10.** | Associations between SES and white matter microstructure, adjusting for race/ethnicity |
| 33-34 | **eTable 11.** | PCA loadings of RSI and DTI metrics that were significantly associated with SES |
| 35 | **eTable 12.** | Associations between SES and white matter microstructure PCs |
| 36 | **eTable 13.** | Associations between SES and obesity-related measures |
| 37 | **eTable 14.** | Indirect effects of obesity-related measures on associations between SES and white matter microstructure |
| 38 | **eTable 15.** | Associations between SES and cognitive performance |
| 39 | **eTable 16.** | Indirect effects of total cognition score on associations between SES and white matter microstructure |
| 40 | **eTable 17.** | Fit indices for main indirect effects models |
| 41-42 | **eTable 18.** | Indirect effects of cognitive task scores on associations between SES and white matter microstructure |
| 43-44 | **eTable 19.** | Indirect effects of white matter microstructure on associations between SES and cognitive performance |
| 45-46 | **eTable 20.** | Indirect effects model testing in random subsample with one participant per family |
| 47 | **References** | References |

***Co-corresponding authors:**

Scott Marek, PhD

Assistant Professor

Mallinckrodt Institute of Radiology

Washington University in St. Louis School of Medicine

St. Louis, MO 63110

MSC 8134-0070-02

Tamara Hershey, PhD

James S. McDonnell Professor of Cognitive Neuroscience

Department of Psychiatry and Mallinckrodt Institute of Radiology

Washington University in St. Louis School of Medicine

St. Louis, MO 63110

MSC 8134-0070-02

**eMethods.**

**Exclusion of participants with history of severe neurological and psychiatric conditions.** Exclusionary neurological conditions included cerebral palsy, brain tumor, stroke, aneurysm, brain hemorrhage, hematoma, epilepsy, seizures, intellectual disability, traumatic brain injury, lead poisoning, muscular dystrophy, and multiple sclerosis. Exclusionary psychiatric conditions included schizophrenia, autism spectrum disorder, and substance use disorder.

**Derivation of household income.** The Adolescent Brain Cognitive Development (ABCD) Study defined household income as total combined family income in the past 12 months, including pre-tax income, wages, rent, social security, benefits, compensation, help from others, etc. Household income was reported in income brackets at non-regular intervals: Less than $5,000; $5,000 through $11,999; $12,000 through $15,999; $16,000 through $24,999; $25,000 through $34,999; $35,000 through $49,999; $50,000 through $74,999; $75,000 through $99,999; $100,000 through $199,999; and $200,000 and greater. As indirect effects analyses require all variables of interest to be continuous, we followed the procedure by Hackman et al. (2021) that divided the midpoint of each income bracket by $10,000 and scaling the results^1^. The midpoints after division were: 0.25, 0.85, 1.4, 2.05, 3, 4.25, 6.25, 8.75, 15, and 20.

**Derivation of parental education.** The parental education levels reported by the ABCD Study were converted into estimated years of schooling per U.S. conventions, as previously used by Rakesh et al. (2021) amongst others^2^. This procedure was meant to meet the requirement that variables must be continuous in indirect effects analyses.

| **Education level** | **Estimated years** | **Education level** | **Estimated years** | **Education level** | **Estimated years** |
| --- | --- | --- | --- | --- | --- |
| Never attended/  Kindergarten only | 0 | 8^th^ grade | 8 | Associate degree: Occupational | 14 |
| 1^st^ grade | 1 | 9^th^ grade | 9 | Associate degree: Academic Program | 14 |
| 2^nd^ grade | 2 | 10^th^ grade | 10 | Bachelor's degree | 16 |
| 3^rd^ grade | 3 | 11^th^ grade | 11 | Master's degree | 18 |
| 4^th^ grade | 4 | 12^th^ grade | 12 | Professional School degree (ex. MD) | 20 |
| 5^th^ grade | 5 | High school graduate | 12 | Doctoral degree (ex. PhD) | 22 |
| 6^th^ grade | 6 | GED or equivalent Diploma | 12 |  |  |
| 7^th^ grade | 7 | Some college | 14 |  |  |

**Magnetic resonance imaging (MRI) sequence specification**^3^**.** T1-weighted anatomical images were collected as a 3D T1-weighted inversion prepared RF-spoiled gradient echo scan, with voxel resolution of 1 mm^3^ isotropic. Diffusion-weighed images (DWIs) were collected using a multi-shell, multiband echo-planar imaging sequence (acquisition time = 7:31, repetition time = 4100 ms, echo time = 88 ms, matrix size = 140 × 140 × 81, flip angle = 90°, acceleration factor = 3, and voxel resolution = 1.7 mm^3^ isotropic). DWIs were imaged with 7 b = 0 frames and 96 gradient directions (b’s = 500, 1000, 2000, 3000 s/mm^2^ with 6, 15, 15, and 60 directions, respectively).

**Cognitive testing.** Cognitive performance was assessed using the NIH Toolbox Cognition Battery. This suite of seven tests included: flanker inhibitory control test (assessing cognitive control, attention), list sorting working memory test (assessing working memory), dimensional change card sort (assessing executive functioning), oral reading recognition test (assessing reading and language ability), pattern comparison test (assessing processing speed), picture sequencing test (assessing episodic memory), and picture vocabulary test (assessing verbal ability). The crystallized cognition score is a composite of the oral reading recognition and picture vocabulary tests, and assesses learned experience. The fluid cognition score is a composite of the flanker inhibitory control, list sorting working memory, dimension change card sort, pattern comparison, and picture sequencing tests, and assesses ability for new learning and information processing^4,5^. The total cognition score is a composite of all seven individual tests. Age-corrected scores were used in the current analyses.

**eFigure 1. Flowchart of participant selection**

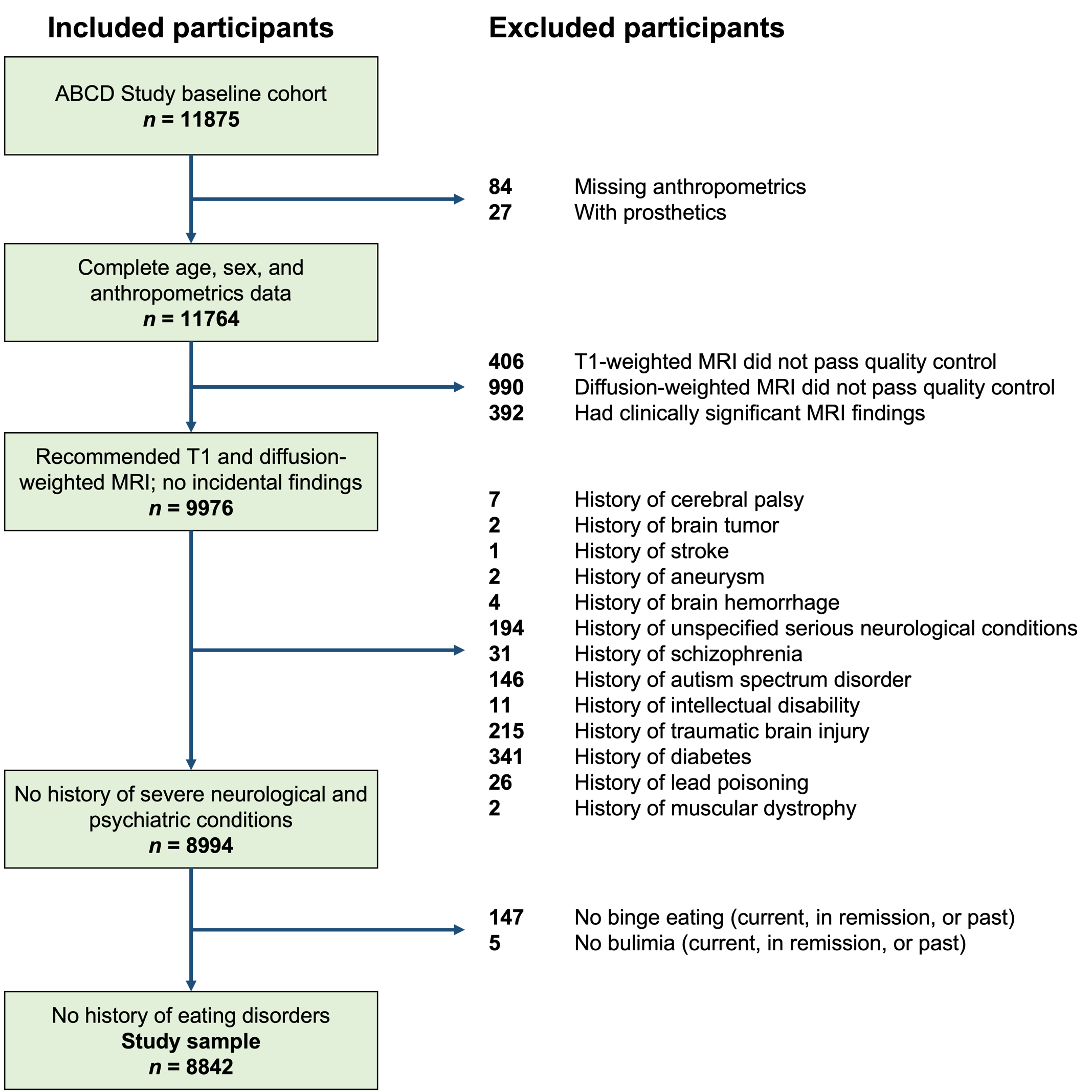

*Note.* Items that were screened (e.g., history of anorexia nervosa) but had no participant for exclusion are omitted. ABCD, Adolescent Brain Cognitive Development; MRI, magnetic resonance imaging

**eTable 1. Lookup list for relevant ABCD Study instrument names**

| **Variable** | **Data structure (instrument)** | **Data element (item)** |
| --- | --- | --- |
| ***Variables included in statistical analyses*** | | |
| **Age** | ABCD Youth Anthropometrics (Modified from PhenX) | interview_age |
| **Sex** |  | sex |
| **Obesity-related measures** |  | anthroheightcalc, anthroweight1lb, anthroweight2lb, anthro_waist_cm, anthroweightcast |
| **Family ID** | ABCD ACS Post Stratification Weights | rel_family_id |
| **Race/ethnicity** |  | race_ethnicity |
| **Pubertal development scale (PDS)** | ABCD Sum Scores Physical Health Parent | pds_p_ss_female_category_2, pds_p_ss_male_category_2 |
| **Intracranial volume (ICV)** | ABCD sMRI Part 1 | smri_vol_scs_intracranialv |
| **Neighborhood disadvantage** | Residential History Derived Scores | reshist_addr1_valid, reshist_addr1_adi_edu_l, reshist_addr1_adi_edu_h, reshist_addr1_adi_work_c, reshist_addr1_adi_income, reshist_addr1_adi_in_dis, reshist_addr1_adi_home_v, reshist_addr1_adi_rent, reshist_addr1_adi_mortg, reshist_addr1_adi_home_o, reshist_addr1_adi_crowd, reshist_addr1_adi_unemp, reshist_addr1_adi_pov, reshist_addr1_adi_b138, reshist_addr1_adi_sp, reshist_addr1_adi_ncar, reshist_addr1_adi_ntel, reshist_addr1_adi_nplumb |
| **Household income** | ABCD Parent Demographics Survey | demo_comb_income_v2 |
| **Parental education** |  | demo_prnt_ed_v2, demo_prtnr_ed_v2 |
| **Mean head motion** | ABCD dMRI RSI Part 1 | dmri_rsi_meanmotion |
| **White matter RSI-RNI** |  | All that starts with dmri_rsirni_fib_... |
| **White matter RSI-RND** | ABCD dMRI RSI Part 2 | All that starts with dmri_rsirnd_fib_... |
| **White matter DTI-FA** | ABCD dMRI DTI Part 1 | All that starts with dmri_dtifa_fiberat_... |
| **White matter DTI-MD** |  | All that starts with dmri_dtimd_fiberat_... |
| **NIH Toolbox cognition** | ABCD Youth NIH TB Summary Scores | nihtbx_picvocab_agecorrected, nihtbx_flanker_agecorrected, nihtbx_list_agecorrected, nihtbx_cardsort_agecorrected, nihtbx_pattern_agecorrected, nihtbx_picture_agecorrected, nihtbx_reading_agecorrected, nihtbx_fluidcomp_agecorrected, nihtbx_cryst_agecorrected, nihtbx_totalcomp_agecorrected |
| **Scanner ID** | ABCD MRI Info | mri_info_deviceserialnumber |
| **Diagnosis of common psychiatric conditions** | ABCD Screener | scrn_commondx |
| **Adverse childhood experience (ACE)** | ABCD Parent Diagnostic Interview for DSM-5 (KSADS) Traumatic Events | ksads_ptsd_raw_754_p, ksads_ptsd_raw_755_p, ksads_ptsd_raw_756_p, ksads_ptsd_raw_757_p, ksads_ptsd_raw_758_p, ksads_ptsd_raw_759_p, ksads_ptsd_raw_760_p, ksads_ptsd_raw_761_p, ksads_ptsd_raw_762_p, ksads_ptsd_raw_763_p, ksads_ptsd_raw_764_p, ksads_ptsd_raw_765_p, ksads_ptsd_raw_766_p, ksads_ptsd_raw_767_p, ksads_ptsd_raw_768_p, ksads_ptsd_raw_769_p, ksads_ptsd_raw_770_p |
| **Preterm birth** | ABCD Developmental History Questionnaire | devhx_12a_p |
| ***Variables as part of inclusion/exclusion criteria*** | | |
| **MRI quality control** | ABCD Recommended Imaging Inclusion | imgincl_t1w_include, imgincl_dmri_include |
| **MRI incidental findings** | ABCD MR Findings | mrif_score |
| **History of severe neurological conditions** | ABCD Screener | scrn_cpalsy, scrn_tumor, scrn_stroke, scrn_aneurysm, scrn_hemorrhage, scrn_hemotoma, scrn_medcond_other, scrn_epls, scrn_seizure, scrn_con_excl, scrn_intdisab, scrn_tbi_loc, scrn_tbi_mem, scrn_tbi_scan |
|  | ABCD Parent Medical History Questionnaire | medhx_2k, medhx_2l, medhx_2m |
| **History of severe psychiatric conditions** | ABCD Screener | scrn_schiz, scrn_asd, scrn_sud |
| **History of obesity comorbidities** | ABCD Parent Medical History Questionnaire | medhx_2g |
|  | ABCD Parent Diagnostic Interview for DSM-5 Full (KSADS-5) | ksads_13_929_p, ksads_13_930_p, ksads_13_931_p, ksads_13_932_p, ksads_13_933_p, ksads_13_934_p, ksads_13_935_p, ksads_13_936_p, ksads_13_937_p, ksads_13_938_p, ksads_13_939_p, ksads_13_940_p |

*Note.* Data elements are shown in the they appear in the ABCD Data Dictionary at <https://nda.nih.gov/data_dictionary.html?source=ABCD%2BRelease%2B4.0>. RSI, restriction spectrum imaging; RNI, restricted normalized isotropic; RND, restricted normalized directional; DTI, diffusion tensor imaging; FA, fractional anisotropy; MD, mean diffusivity; MRI, magnetic resonance imaging

**eTable 2. Factor loadings of ADI metrics**

| **ADI metric description** | **Factor loading** | **Inclusion in neighborhood disadvantage** |
| --- | --- | --- |
| Percentage of population aged ≥ 25 years with < 9 years of education | 0.63 | Yes |
| Percentage of population aged ≥ 25 years with at least a high school diploma | 0.77^a^ | Yes |
| Percentage of employed persons aged ≥ 16 years in white collar occupations | 0.18^a^ | No |
| Median family income | 0.79^a^ | Yes |
| Income disparity, defined by Singh as the log of 100 × ratio of the number of households with < $10000 annual income to the number of households with > $50000 annual income. | 0.86 | Yes |
| Median home value | 0.45^a^ | No |
| Median gross rent | 0.52 | No |
| Median monthly mortgage | 0.51 | No |
| Percentage of homeowners | 0.75^a^ | Yes |
| Percentage of occupied housing units with > 1 person per room (i.e., crowding) | 0.52 | No |
| Percentage of civilian labor force aged ≥ 16 years unemployed (i.e., unemployment rate) | 0.73 | Yes |
| Percentage of families below the poverty level | 0.94 | Yes |
| Percentage of population below 138% of the poverty threshold | 0.98 | Yes |
| Percentage of single parent households | 0.83 | Yes |
| Percentage of occupied housing units without a motor vehicle | 0.70 | Yes |
| Percentage of occupied housing units without a telephone | 0.40 | No |
| Percentage of occupied housing units without complete plumbing | 0.22 | No |

*Note.* An exploratory factor analysis based on 7912 participants with complete data of all 17 Area Deprivation Index (ADI) metrics suggested a one-factor solution; factors with strong loadings (≥ 0.63) were included in the neighborhood disadvantage variable. Kaiser-Meyer-Olkin (KMO) factor adequacy = 0.9; Bartlett’s test of sphericity chi-sq = 131708.7, *p* < 0.001; and positive determinant = 5.802e-08 all suggested factorability. ^a^Metric was reverse-coded to give consistent direction amongst metrics such that higher neighborhood disadvantage reflected greater impoverishment.

**eFigure 2. Isosurface renderings of white matter tracts**

*Note.* Coronal and sagittal views of all (in scale relative to each other) and individual major white matter tracts (each scaled differently for ease of visualization). The forceps major (Fmaj) and forceps minor (Fmin) are portions of the corpus callosum (CC). The superior longitudinal fasciculus (not shown) was partitioned into temporal (tSLF) and parietal (pSLF) subregions for each hemisphere. R, right; L, left; A, anterior; P, posterior.

**eFigure 3. Relationships between missingness of key variables**

**A) Missingness in SES indicators and demographic variables**

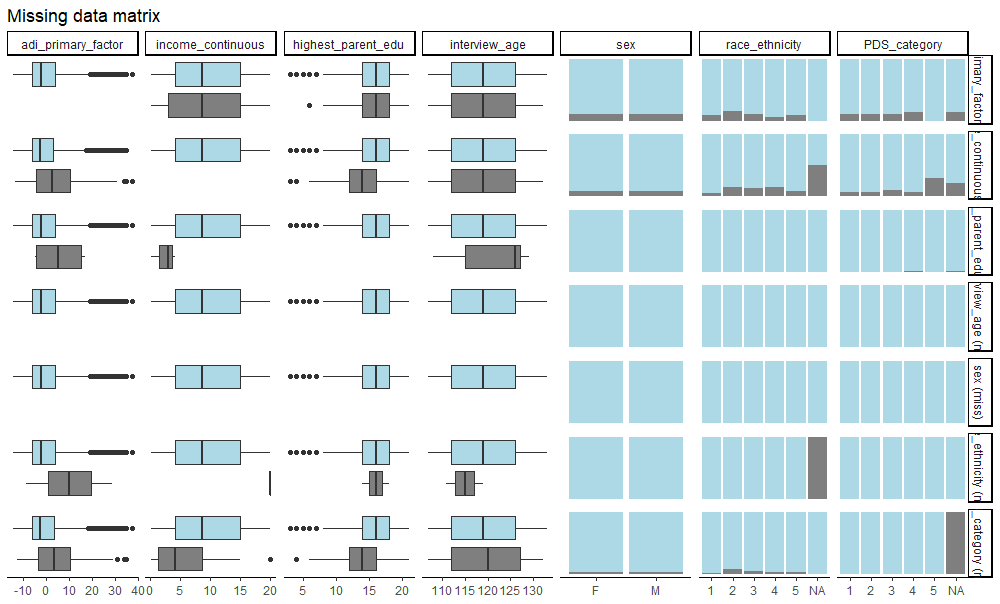

**B) Missingness in SES indicators and obesity-related measures**

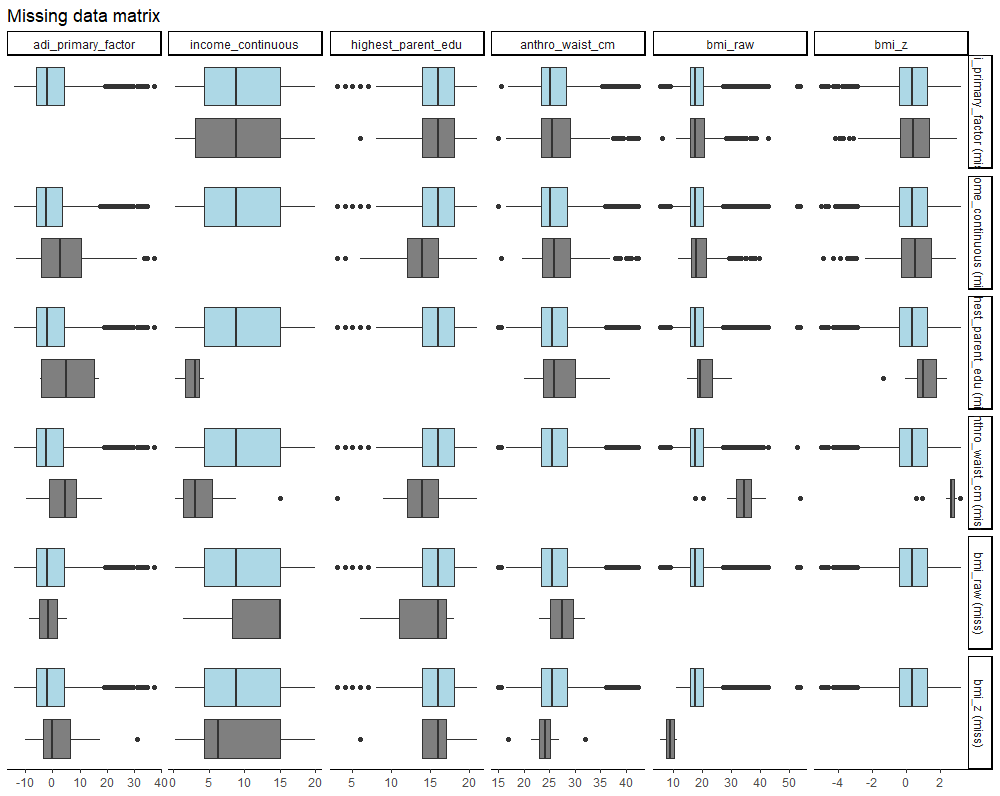

**C) Missingness in SES indicators and cognition**

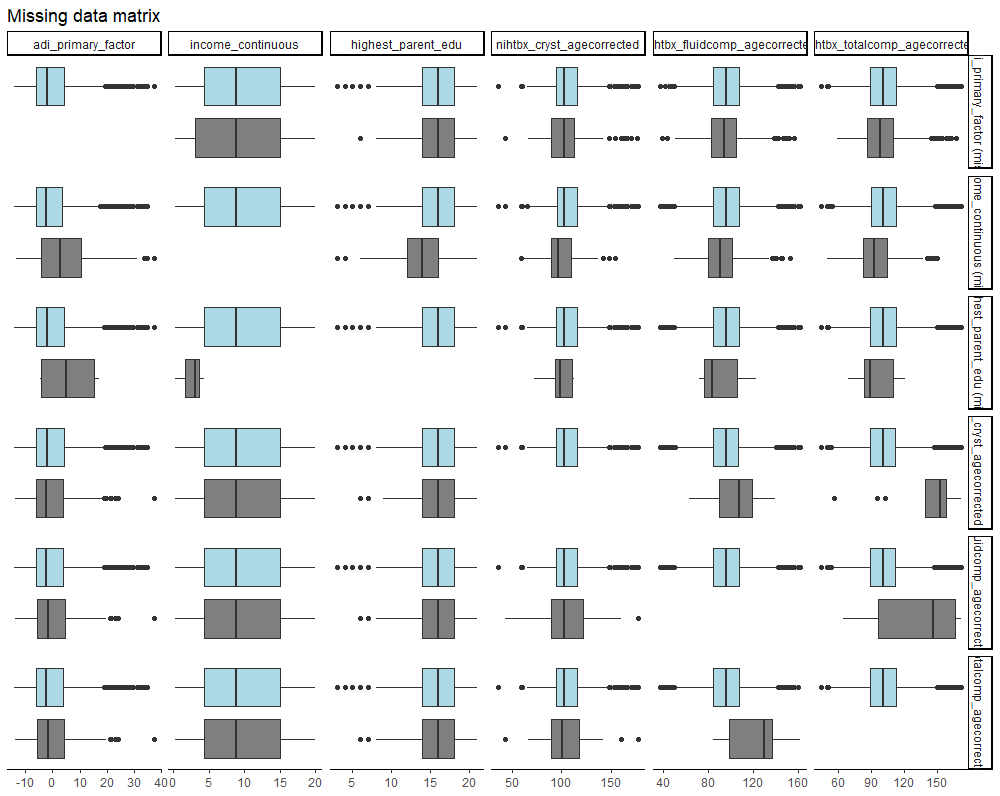

*Note*. While all relevant variables in the current study had at most 8.29% of missing cases (see **eTable 3** in the **Supplement** for tally), selecting only complete data would trim our sample to 6513 participants (a 26% reduction), considerably hampering statistical power. Furthermore, analyses using the “finalfit” package^6^, as shown here, suggested some associations between missingness of socioeconomic status (SES) indicators and demographics. Missingness was represented by grey color. For example, panel A shows that the missingness in neighborhood disadvantage score (“adi_primary_factor”; row 1) seemed more prevalent in Black participants (“race_ethnicity” of 2; column 6), and that the missingness in household income (“income_continouous”; row 2) was greater in participants with higher neighborhood disadvantage (column 1) and of non-White race/ethnicity (column 6). Nonetheless, missingness in obesity-related measures and cognitive performance was unrelated to SES, and so was the case with neuroimaging metrics (not shown here for brevity).

**eTable 3. Missing data and roles of variables in imputation**

| **Variable** | **Missing cases** (out of 8842) | **Imputed?** | **Predictor in imputation?** |
| --- | --- | --- | --- |
| **Age** | 0 (0%) | No | Yes |
| **Sex** | 0 (0%) | No | Yes |
| **Race/ethnicity** | 2 (0.02%) | Yes | Yes |
| **PDS score** | 311 (3.52%) | Yes | Yes |
| **Scanner ID** | 0 (0%) | No | No |
| **Family ID** | 0 (0%) | No | No |
| **Neighborhood disadvantage** | 930 (10.52%) | Yes | Yes |
| **Household income** | 733 (8.29%) | Yes | Yes |
| **Parental education** | 9 (0.10%) | Yes | Yes |
| **White matter microstructure** |  |  |  |
| **RSI-RND** | 1 – 25 (0.01% – 0.28%) | No | Yes |
| **RSI-RNI** | 6 – 59 (0.07% – 0.67%) | No | Yes |
| **DTI-FA** | 1 – 11 (0.01% – 0.12%) | No | Yes |
| **DTI-MD** | 2 – 44 (0.02% – 0.50%) | No | Yes |
| **Mean head motion** | 0 (0%) | No | Yes |
| **ICV** | 0 (0%) | No | Yes |
| **Obesity-related measures** |  |  |  |
| **Waist circumference** | 23 (0.26%) | Yes | Yes |
| **BMI** | 3 (0.03%) | Yes | Yes |
| **BMI *z*-score** | 23 (0.26%) | Yes | Yes |
| **Cognition** |  |  |  |
| **Flanker inhibitory control** | 123 (1.39%) | No | Yes |
| **Picture sequencing** | 117 (1.32%) | No | Yes |
| **List sorting working memory** | 148 (1.67%) | No | Yes |
| **Picture vocabulary** | 129 (1.46%) | No | Yes |
| **Oral reading** | 149 (1.69%) | No | Yes |
| **Dimensional change card sort** | 131 (1.48%) | No | Yes |
| **Pattern comparison** | 132 (1.49%) | No | Yes |
| **Crystallized cognition composite** | 276 (3.12%) | No | Yes |
| **Fluid cognition composite** | 302 (3.42%) | No | Yes |
| **Total cognition composite** | 302 (3.42%) | No | Yes |
| **ACE total number** | 133 (1.50%) | No | No |
| **Common psychiatric diagnoses status** | 0 (0%) | No | No |
| **Preterm birth status** | 92 (1.04%) | No | No |

*Note.* Missing cases are reported as count (frequency). White matter microstructure and neurocognition variables were not imputed as they served as outcome variables in analyses; other variables that were not imputed include those that did not have missing cases and those that were used only in sensitivity analyses. To lower computational cost, Scanner ID and family ID were not modeled as predictors used to impute missing cases. PDS, pubertal development stage; RSI, restriction spectrum imaging; RND, restricted normalized directional; RNI, restricted normalized isotropic; DTI, diffusion tensor imaging; FA, fractional anisotropy; MD, mean diffusivity; BMI, body mass index; ACE, adverse childhood experience

**eTable 4. Distribution of imputed variables before and after imputation of missing data**

| **Variable** | **Original (non-imputed) dataset** | **Multiply imputed datasets** |
| --- | --- | --- |
| ***Demographic variables*** | | |
| **Race/ethnicity** |  |  |
| White | 4738 (53.6%) | 4739 (53.6%) |
| Black | 1212 (13.7%) | 1213 (13.7%) |
| Hispanic | 1805 (20.4%) | 1805 (20.4%) |
| Asian | 183 (2.1%) | 183 (2.1%) |
| Other | 902 (10.2%) | 902 (10.2%) |
| **PDS** |  |  |
| Pre-puberty | 4425 (51.9%) | 4561 (51.6%) |
| Early puberty | 2009 (23.5%) | 2088 (23.6%) |
| Mid-puberty | 1966 (23.0%) | 2054 (23.2%) |
| Late puberty | 124 (1.5%) | 131 (1.5%) |
| Post-puberty | 7 (0.1%) | 7 (0.1%) |
| ***SES indicators*** | | |
| **Neighborhood disadvantage** | 0 ± 8.2 (range: -14.3 to 37.5) | 0 ± 8.3 (range: -14.3 to 37.5) |
| **Household income** | 10.0 ± 6.2 (range: 0.3 to 20.0) | 9.8 ± 6.2 (range: 0.3 to 20.0) |
| **Parental education (years)** | 15.9 ± 2.8 (range: 3.0 to 21.0) | 15.9 ± 2.8 (range: 3.0 to 21.0) |
| ***Obesity-related measures*** | | |
| **Waist circumference (in)** | 26.3 ± 4.0 (range: 15.0 to 42.5) | 26.3 ± 4.0 (range: 15.0 to 42.5) |
| **BMI (kg/m^2^)** | 18.6 ± 4.1 (range: 5.3 to 53.9) | 18.6 ± 4.1 (range: 5.3 to 53.9) |
| **BMI *z*-score** | 0.4 ± 1.2 (range: -5.0 to 3.2) | 0.4 ± 1.2 (range: -5.0 to 3.2) |

*Note*. Distributions of imputed demographic variables, SES indicators, and obesity-related measures were consistent before and after imputation of missing data. Results are shown as mean ± standard deviation for continuous variables and count (frequency) for categorical variables. Variables that were not imputed are not shown here. PDS, pubertal development stage; SES, socioeconomic status; BMI, body mass index

**eTable 5. Pre- and post-harmonization associations between preserved variables and RSI and DTI metrics**

| **Variable (IV)** | **Association with RSI and DTI metrics (DVs)**  **before harmonization, *R*^2^ (%)** | | | | **Association with RSI and DTI metrics (DVs)**  **after harmonization, *R*^2^ (%)** | | | |
| --- | --- | --- | --- | --- | --- | --- | --- | --- |
|  | **Mean** | **SD** | **Min** | **Max** | **Mean** | **SD** | **Min** | **Max** |
| **Age** | 1.33 | 1.04 | 0.01 | 4.61 | 1.84 | 1.36 | -0.01 | 5.63 |
| **Sex** | 0.75 | 1.05 | -0.01 | 3.73 | 0.95 | 1.22 | -0.01 | 4.32 |
| **Race/ethnicity** | 0.67 | 0.45 | 0 | 2.58 | 0.68 | 0.59 | -0.02 | 3.05 |
| **PDS** | 0.54 | 0.65 | -0.04 | 3.16 | 0.74 | 0.87 | -0.04 | 3.51 |
| **Neighborhood disadvantage** | 0.40 | 0.43 | -0.01 | 1.60 | 0.26 | 0.32 | -0.01 | 1.45 |
| **Household income** | 0.18 | 0.17 | -0.01 | 1.00 | 0.31 | 0.24 | -0.01 | 0.94 |
| **Parental education** | 0.22 | 0.22 | -0.01 | 1.21 | 0.30 | 0.29 | -0.01 | 1.20 |

*Note*. Associations between preserved variables and RSI and DTI metrics seemed only limitedly affected by ComBat harmonization. Associations were estimated in linear regression models in which each of the RSI and DTI metrics (as dependent variable (DV)) was regressed onto each of the independent variable (IV). The post-harmonization associations were tested in a randomly chosen dataset out of 50 imputed datasets. RSI, restriction spectrum imaging; DTI, diffusion tensor imaging; SD, standard deviation; PDS, pubertal development stage

**eTable 6. Associations between SES and race/ethnicity**

| **Race/ethnicity (IV)** | **SES indicator (DV)** | | | | | |
| --- | --- | --- | --- | --- | --- | --- |
|  | **Neighborhood disadvantage**  (higher = lower SES) | | **Household income**  (higher = higher SES) | | **Parental education**  (higher = higher SES) | |
|  | ***β*** | **Partial *R^2^*** | ***β*** | **Partial *R^2^*** | ***β*** | **Partial *R^2^*** |
| **Black** | 1.26 *** | 0.17 | -1.07 *** | 0.13 | -0.74 *** | 0.06 |
| **Hispanic** | 0.88 *** | 0.13 | -0.75 *** | 0.10 | -0.64 *** | 0.06 |
| **Asian** | -0.05 | < 0.01 | 0.13 * | < 0.01 | 0.29 *** | < 0.01 |
| **Other** | 0.36 *** | 0.02 | -0.31 *** | 0.01 | -0.20 *** | < 0.01 |

*Note.* Socioeconomic status (SES) was highly entangled with race/ethnicity. Comparing effect sizes across the three SES indicators, it appears that neighborhood disadvantage is most confounded with race/ethnicity, followed by household income, and lastly parental education. This may explain why many white matter microstructure associations with neighborhood disadvantage and household income diminished more in magnitude once analyses controlled for race/ethnicity. Analyses were linear regression models where race/ethnicity was the independent variable (IV) of interest and each of the SES indicator was the dependent variable (DV). Results were in reference to White participants and were standardized. *, *p* < 0.05; ***, *p* < 0.001.

**eTable 7. Sample sizes for models testing associations between SES, RSI and DTI metrics, and neurocognition**

| **Metric/Region** | **Sample size (*n*)** | | | |
| --- | --- | --- | --- | --- |
| **White matter tract** | **RSI-RND** | **RSI-RNI** | **DTI-FA** | **DTI-MD** |
| Fx (right) | 8832 | 8812 | 8840 | 8807 |
| Fx (left) | 8829 | 8793 | 8841 | 8799 |
| CgC (right) | 8837 | 8815 | 8839 | 8832 |
| CgC (left) | 8826 | 8803 | 8839 | 8829 |
| CgH (right) | 8836 | 8819 | 8838 | 8814 |
| CgH (left) | 8828 | 8799 | 8836 | 8818 |
| CST (right) | 8824 | 8825 | 8837 | 8827 |
| CST (left) | 8823 | 8817 | 8837 | 8822 |
| ATR (right) | 8839 | 8789 | 8839 | 8823 |
| ATR (left) | 8833 | 8786 | 8838 | 8821 |
| Unc (right) | 8830 | 8826 | 8833 | 8837 |
| Unc (left) | 8824 | 8826 | 8831 | 8833 |
| ILF (right) | 8830 | 8834 | 8840 | 8839 |
| ILF (left) | 8834 | 8829 | 8838 | 8836 |
| IFOF (right) | 8835 | 8830 | 8838 | 8834 |
| IFOF (left) | 8839 | 8801 | 8838 | 8831 |
| Fmaj | 8822 | 8798 | 8832 | 8815 |
| Fmin | 8824 | 8819 | 8832 | 8814 |
| CC | 8824 | 8820 | 8835 | 8831 |
| SLF (right) | 8830 | 8835 | 8839 | 8840 |
| SLF (left) | 8822 | 8824 | 8838 | 8839 |
| tSLF (right) | 8830 | 8835 | 8840 | 8839 |
| tSLF (left) | 8826 | 8821 | 8839 | 8838 |
| pSLF (right) | 8827 | 8836 | 8839 | 8840 |
| pSLF (left) | 8817 | 8826 | 8835 | 8838 |
| SCS (right) | 8841 | 8830 | 8840 | 8839 |
| SCS (left) | 8839 | 8824 | 8840 | 8838 |
| SIFC (right) | 8838 | 8829 | 8839 | 8830 |
| SIFC (left) | 8840 | 8828 | 8838 | 8818 |
| IFSFC (right) | 8829 | 8833 | 8840 | 8838 |
| IFSFC (left) | 8826 | 8833 | 8839 | 8837 |
| **Cognition** | **Score** | | | |
| Picture vocabulary | 8713 | | | |
| Flanker inhibitory control | 8719 | | | |
| List sorting | 8694 | | | |
| Dimensional card sort | 8711 | | | |
| Pattern comparison | 8710 | | | |
| Picture sequence | 8725 | | | |
| Oral reading | 8693 | | | |
| Fluid cognition | 8540 | | | |
| Crystallized cognition | 8566 | | | |
| Total cognition | 8540 | | | |

*Note*. Sample sizes for models testing associations between socioeconomic status (SES) and RSI/DTI metrics and cognition varied as both imaging and cognitive measures had unimputed missing values. In comparison, models testing associations between SES and obesity-related measures had sample sizes of 8824 due to complete imputation. RSI, restriction spectrum imaging; RND, restricted normalized directional; RNI, restricted normalized isotropic; DTI, diffusion tensor imaging; FA, fractional anisotropy; MD, mean diffusivity. Full names of white matter tracts are shown in the main text.

**eFigure 4. Correlations between SES indicators**

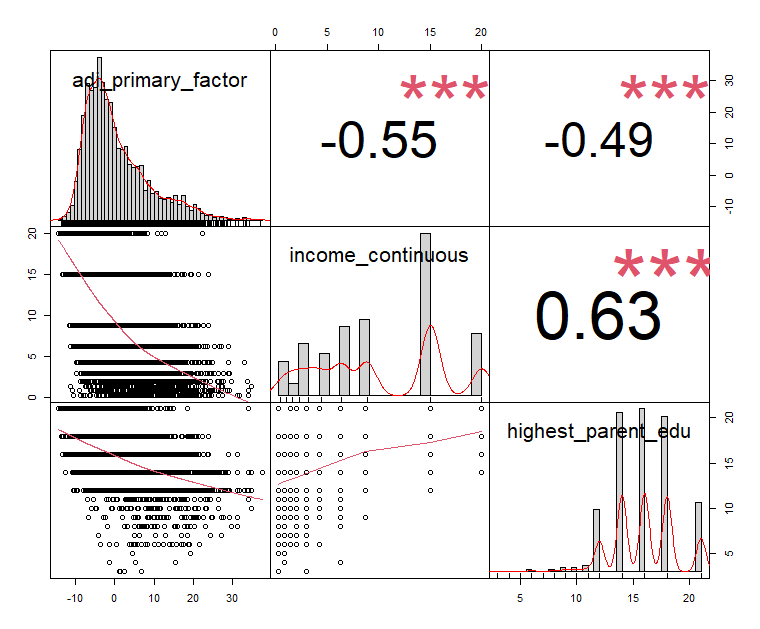

*Note*. Moderate bivariate correlations (Pearson’s *r*) were observed between socioeconomic status (SES) indicators in a randomly selected imputed dataset out of 50. ***, *p* < 0.001.

**eTable 8. Associations between SES and white matter microstructure**

| **White matter tracts** | **SES indicators (IVs)** | | | | | | | | |
| --- | --- | --- | --- | --- | --- | --- | --- | --- | --- |
|  | **Neighborhood disadvantage**  (higher = lower SES) | | | **Household income**  (higher = higher SES) | | | **Parental education**  (higher = higher SES) | | |
|  | ***β* (95% CI)** | ***p*-value**  (nominal) | ***p*-value**  (FDR) | ***β* (95% CI)** | ***p*-value**  (nominal) | ***p*-value**  (FDR) | ***β* (95% CI)** | ***p*-value**  (nominal) | ***p*-value**  (FDR) |
| **A) RSI-RND (DV)** | | | | | | | | | |
| Fx (right) | 0.001 (-0.026 to 0.028) | 0.94 | 0.97 | 0.024 (-0.007 to 0.055) | 0.13 | 0.51 | 0.026 (-0.003 to 0.054) | 0.08 | 0.15 |
| Fx (left) | -0.009 (-0.037 to 0.018) | 0.50 | 0.68 | 0.014 (-0.018 to 0.045) | 0.39 | 0.56 | 0.031 (0.002 to 0.060) | 0.04 | 0.09 |
| CgC (right) | 0.032 (0.005 to 0.059) | 0.02 | 0.11 | -0.033 (-0.064 to -0.003) | 0.03 | 0.39 | 0.010 (-0.018 to 0.038) | 0.48 | 0.55 |
| CgC (left) | 0.017 (-0.010 to 0.044) | 0.21 | 0.45 | -0.023 (-0.053 to 0.007) | 0.13 | 0.51 | -0.013 (-0.041 to 0.015) | 0.37 | 0.50 |
| CgH (right) | -0.013 (-0.040 to 0.014) | 0.34 | 0.52 | -0.010 (-0.040 to 0.021) | 0.54 | 0.73 | 0.033 (0.005 to 0.061) | 0.02 | 0.08 |
| CgH (left) | -0.002 (-0.029 to 0.024) | 0.87 | 0.93 | 0.015 (-0.015 to 0.045) | 0.32 | 0.54 | 0.030 (0.002 to 0.058) | 0.04 | 0.09 |
| CST (right) | 0.012 (-0.014 to 0.039) | 0.35 | 0.52 | 0.013 (-0.017 to 0.043) | 0.39 | 0.56 | 0.042 (0.015 to 0.069) | 0.003 | **0.01** |
| CST (left) | 0.026 (0.000 to 0.053) | 0.05 | 0.21 | 0.007 (-0.023 to 0.037) | 0.64 | 0.83 | 0.049 (0.021 to 0.077) | 0.001 | **0.004** |
| ATR (right) | 0.030 (0.003 to 0.057) | 0.03 | 0.16 | 0.019 (-0.012 to 0.050) | 0.24 | 0.52 | 0.018 (-0.010 to 0.047) | 0.21 | 0.29 |
| ATR (left) | 0.007 (-0.021 to 0.035) | 0.61 | 0.79 | 0.026 (-0.005 to 0.057) | 0.10 | 0.51 | 0.019 (-0.010 to 0.047) | 0.19 | 0.29 |
| Unc (right) | 0.024 (-0.003 to 0.051) | 0.08 | 0.26 | -0.046 (-0.076 to 0.016) | 0.002 | 0.07 | 0.009 (-0.018 to 0.037) | 0.50 | 0.56 |
| Unc (left) | 0.029 (0.001 to 0.056) | 0.04 | 0.18 | -0.032 (-0.062 to -0.002) | 0.04 | 0.39 | 0.010 (-0.018 to 0.038) | 0.47 | 0.55 |
| ILF (right) | 0.002 (-0.025 to 0.029) | 0.87 | 0.93 | -0.016 (-0.046 to 0.014) | 0.30 | 0.54 | 0.021 (-0.007 to 0.048) | 0.15 | 0.24 |
| ILF (left) | 0.019 (-0.008 to 0.046) | 0.17 | 0.44 | -0.001 (-0.031 to 0.029) | 0.94 | 0.98 | 0.020 (-0.008 to 0.047) | 0.16 | 0.25 |
| IFOF (right) | 0.004 (-0.024 to 0.030) | 0.79 | 0.93 | -0.028 (-0.058 to 0.002) | 0.07 | 0.51 | 0.026 (-0.002 to 0.054) | 0.06 | 0.13 |
| IFOF (left) | 0.002 (-0.024 to 0.029) | 0.86 | 0.93 | -0.020 (-0.050 to 0.009) | 0.18 | 0.52 | 0.027 (-0.001 to 0.054) | 0.06 | 0.12 |
| Fmaj | -0.040 (-0.067 to -0.013) | 0.004 | **0.03** | 0.022 (-0.008 to 0.053) | 0.15 | 0.51 | -0.012 (-0.040 to 0.016) | 0.40 | 0.52 |
| Fmin | 0.019 (-0.008 to 0.045) | 0.17 | 0.44 | -0.018 (-0.048 to 0.012) | 0.24 | 0.52 | -0.010 (-0.038 to 0.017) | 0.47 | 0.55 |
| CC | -0.014 (-0.041 to 0.012) | 0.29 | 0.50 | 0.001 (-0.029 to 0.030) | 0.97 | 0.98 | -0.004 (-0.032 to 0.023) | 0.75 | 0.77 |
| SLF (right) | -0.017 (-0.044 to 0.010) | 0.22 | 0.45 | -0.001 (-0.031 to 0.029) | 0.96 | 0.98 | 0.053 (0.025 to 0.080) | < 0.001 | **0.002** |
| SLF (left) | -0.055 (-0.081 to -0.028) | < 0.001 | **0.001** | 0.014 (-0.016 to 0.044) | 0.35 | 0.55 | 0.050 (0.022 to 0.077) | < 0.001 | **0.003** |
| tSLF (right) | -0.018 (-0.045 to 0.009) | 0.19 | 0.44 | 0.000 (-0.031 to 0.030) | 0.98 | 0.98 | 0.044 (0.016 to 0.072) | 0.002 | **0.01** |
| tSLF (left) | -0.056 (-0.083 to -0.030) | < 0.001 | **0.001** | 0.019 (-0.011 to 0.049) | 0.22 | 0.52 | 0.042 (0.015 to 0.070) | 0.003 | **0.01** |
| pSLF (right) | -0.018 (-0.045 to 0.009) | 0.19 | 0.44 | -0.001 (-0.031 to 0.029) | 0.96 | 0.98 | 0.054 (0.026 to 0.081) | < 0.001 | **0.002** |
| pSLF (left) | -0.040 (-0.066 to 0.013) | 0.003 | **0.03** | 0.003 (-0.027 to 0.032) | 0.86 | 0.98 | 0.059 (0.032 to 0.086) | < 0.001 | **0.001** |
| SCS (right) | 0.015 (-0.012 to 0.042) | 0.27 | 0.50 | -0.015 (-0.046 to 0.015) | 0.33 | 0.54 | 0.032 (0.004 to 0.060) | 0.03 | 0.09 |
| SCS (left) | 0.000 (-0.026 to 0.027) | 0.98 | 0.98 | 0.004 (-0.025 to 0.034) | 0.77 | 0.95 | 0.030 (0.002 to 0.058) | 0.03 | 0.09 |
| SIFC (right) | 0.014 (-0.013 to 0.042) | 0.31 | 0.51 | -0.023 (-0.054 to 0.007) | 0.13 | 0.51 | -0.009 (-0.037 to 0.019) | 0.52 | 0.56 |
| SIFC (left) | -0.003 (-0.030 to 0.024) | 0.81 | 0.93 | -0.017 (-0.047 to 0.012) | 0.25 | 0.52 | -0.004 (-0.032 to 0.024) | 0.77 | 0.77 |
| IFSFC (right) | -0.011 (-0.038 to 0.015) | 0.40 | 0.57 | 0.019 (-0.010 to 0.049) | 0.20 | 0.52 | 0.020 (-0.007 to 0.048) | 0.14 | 0.24 |
| IFSFC (left) | -0.014 (-0.040 to 0.012) | 0.29 | 0.50 | 0.015 (-0.015 to 0.044) | 0.33 | 0.54 | 0.021 (-0.006 to 0.049) | 0.12 | 0.22 |

**(continued) eTable 8. Associations between SES and white matter microstructure**

| **White matter tracts** | **SES indicators (IVs)** | | | | | | | | |
| --- | --- | --- | --- | --- | --- | --- | --- | --- | --- |
|  | **Neighborhood disadvantage**  (higher = lower SES) | | | **Household income**  (higher = higher SES) | | | **Parental education**  (higher = higher SES) | | |
|  | ***β* (95% CI)** | ***p*-value**  (nominal) | ***p*-value**  (FDR) | ***β* (95% CI)** | ***p*-value**  (nominal) | ***p*-value**  (FDR) | ***β* (95% CI)** | ***p*-value**  (nominal) | ***p*-value**  (FDR) |
| **B) RSI-RNI (DV)** | | | | | | | | | |
| Fx (right) | 0.046 (0.019 to 0.074) | 0.001 | **0.01** | 0.008 (-0.023 to 0.040) | 0.59 | 0.63 | -0.013 (-0.042 to 0.016) | 0.38 | 0.95 |
| Fx (left) | 0.047 (0.020 to 0.074) | 0.001 | **0.01** | -0.005 (-0.036 to 0.027) | 0.78 | 0.78 | -0.015 (-0.044 to 0.014) | 0.31 | 0.95 |
| CgC (right) | 0.026 (0.000 to 0.053) | 0.05 | 0.18 | -0.033 (-0.063 to -0.003) | 0.03 | **0.05** | -0.012 (-0.041 to 0.016) | 0.39 | 0.95 |
| CgC (left) | 0.019 (-0.009 to 0.046) | 0.18 | 0.52 | -0.036 (-0.067 to -0.005) | 0.02 | **0.04** | -0.018 (-0.046 to 0.011) | 0.23 | 0.95 |
| CgH (right) | 0.061 (0.034 to 0.088) | < 0.001 | **< 0.001** | -0.031 (-0.062 to -0.001) | 0.04 | **0.05** | 0.006 (-0.022 to 0.034) | 0.68 | 0.95 |
| CgH (left) | 0.066 (0.039 to 0.094) | < 0.001 | **< 0.001** | -0.050 (-0.081 to -0.020) | 0.001 | **0.01** | -0.002 (-0.030 to 0.026) | 0.88 | 0.95 |
| CST (right) | 0.037 (0.010 to 0.065) | 0.01 | **0.03** | -0.031 (-0.062 to -0.001) | 0.05 | **0.05** | -0.001 (-0.029 to 0.027) | 0.95 | 0.95 |
| CST (left) | 0.042 (0.015 to 0.069) | 0.002 | **0.01** | -0.044 (-0.074 to -0.013) | 0.01 | **0.01** | -0.003 (-0.031 to 0.025) | 0.83 | 0.95 |
| ATR (right) | 0.045 (0.018 to 0.072) | 0.001 | **0.01** | -0.045 (-0.075 to -0.014) | 0.004 | **0.01** | 0.003 (-0.026 to 0.031) | 0.85 | 0.95 |
| ATR (left) | 0.039 (0.012 to 0.066) | 0.005 | **0.02** | -0.040 (-0.071 to -0.009) | 0.01 | **0.02** | -0.013 (-0.041 to 0.015) | 0.36 | 0.95 |
| Unc (right) | 0.022 (-0.005 to 0.049) | 0.11 | 0.36 | -0.062 (-0.092 to -0.031) | < 0.001 | **0.002** | 0.005 (-0.023 to 0.034) | 0.71 | 0.95 |
| Unc (left) | 0.015 (-0.012 to 0.042) | 0.28 | 0.57 | -0.052 (-0.083 to -0.022) | 0.001 | **0.01** | -0.003 (-0.031 to 0.026) | 0.86 | 0.95 |
| ILF (right) | -0.004 (-0.031 to 0.022) | 0.75 | 0.97 | -0.042 (-0.073 to -0.012) | 0.01 | **0.01** | -0.003 (-0.031 to 0.025) | 0.83 | 0.95 |
| ILF (left) | -0.001 (-0.028 to 0.026) | 0.93 | 0.97 | -0.033 (-0.063 to -0.002) | 0.04 | **0.05** | -0.014 (-0.042 to 0.014) | 0.33 | 0.95 |
| IFOF (right) | 0.001 (-0.026 to 0.028) | 0.94 | 0.97 | -0.056 (-0.086 to -0.026) | < 0.001 | **0.004** | -0.007 (-0.034 to 0.021) | 0.65 | 0.95 |
| IFOF (left) | 0.002 (-0.024 to 0.029) | 0.87 | 0.97 | -0.052 (-0.083 to -0.022) | 0.001 | **0.01** | -0.011 (-0.039 to 0.017) | 0.43 | 0.95 |
| Fmaj | 0.002 (-0.025 to 0.030) | 0.88 | 0.97 | -0.005 (-0.036 to 0.025) | 0.73 | 0.75 | -0.048 (-0.077 to -0.020) | 0.001 | **0.03** |
| Fmin | 0.015 (-0.013 to 0.042) | 0.29 | 0.57 | -0.045 (-0.076 to -0.013) | 0.005 | **0.01** | -0.039 (-0.068 to -0.010) | 0.01 | 0.13 |
| CC | 0.004 (-0.024 to 0.031) | 0.80 | 0.97 | -0.037 (-0.068 to -0.006) | 0.02 | **0.03** | -0.025 (-0.053 to 0.004) | 0.09 | 0.89 |
| SLF (right) | -0.004 (-0.030 to 0.023) | 0.78 | 0.97 | -0.032 (-0.061 to -0.002) | 0.04 | **0.05** | -0.006 (-0.042 to 0.022) | 0.67 | 0.95 |
| SLF (left) | 0.014 (-0.012 to 0.041) | 0.29 | 0.57 | -0.045 (-0.075 to -0.015) | 0.004 | **0.01** | -0.001 (-0.029 to 0.027) | 0.94 | 0.95 |
| tSLF (right) | -0.007 (-0.034 to 0.019) | 0.58 | 0.95 | -0.026 (-0.056 to 0.004) | 0.09 | 0.10 | -0.001 (-0.029 to 0.026) | 0.92 | 0.95 |
| tSLF (left) | 0.013 (-0.014 to 0.040) | 0.33 | 0.61 | -0.044 (-0.074 to -0.014) | 0.004 | **0.01** | 0.002 (-0.026 to 0.029) | 0.91 | 0.95 |
| pSLF (right) | -0.001 (-0.028 to 0.025) | 0.91 | 0.97 | -0.033 (-0.063 to -0.003) | 0.03 | **0.04** | -0.005 (-0.033 to 0.022) | 0.70 | 0.95 |
| pSLF (left) | 0.016 (-0.011 to 0.043) | 0.23 | 0.57 | -0.046 (-0.077 to -0.016) | 0.003 | **0.01** | 0.002 (-0.026 to 0.029) | 0.91 | 0.95 |
| SCS (right) | -0.016 (-0.042 to 0.011) | 0.24 | 0.57 | -0.022 (-0.052 to 0.008) | 0.15 | 0.17 | 0.009 (-0.019 to 0.037) | 0.52 | 0.95 |
| SCS (left) | 0.002 (-0.024 to 0.029) | 0.86 | 0.97 | -0.040 (-0.069 to -0.010) | 0.01 | **0.02** | 0.004 (-0.024 to 0.031) | 0.79 | 0.95 |
| SIFC (right) | -0.008 (-0.035 to 0.019) | 0.57 | 0.95 | -0.045 (-0.076 to -0.014) | 0.004 | **0.01** | -0.007 (-0.035 to 0.022) | 0.65 | 0.95 |
| SIFC (left) | 0.000 (-0.027 to 0.027) | 0.99 | 0.99 | -0.038 (-0.069 to -0.007) | 0.02 | **0.03** | -0.009 (-0.038 to 0.019) | 0.52 | 0.95 |
| IFSFC (right) | 0.003 (-0.024 to 0.029) | 0.85 | 0.97 | -0.046 (-0.077 to -0.016) | 0.003 | **0.01** | 0.008 (-0.019 to 0.036) | 0.55 | 0.95 |
| IFSFC (left) | -0.003 (-0.030 to 0.024) | 0.84 | 0.97 | -0.046 (-0.077 to -0.015) | 0.003 | **0.01** | 0.011 (-0.017 to 0.039) | 0.44 | 0.95 |

**(continued) eTable 8. Associations between SES and white matter microstructure**

| **White matter tracts** | **SES indicators (IVs)** | | | | | | | | |
| --- | --- | --- | --- | --- | --- | --- | --- | --- | --- |
|  | **Neighborhood disadvantage**  (higher = lower SES) | | | **Household income**  (higher = higher SES) | | | **Parental education**  (higher = higher SES) | | |
|  | ***β* (95% CI)** | ***p*-value**  (nominal) | ***p*-value**  (FDR) | ***β* (95% CI)** | ***p*-value**  (nominal) | ***p*-value**  (FDR) | ***β* (95% CI)** | ***p*-value**  (nominal) | ***p*-value**  (FDR) |
| **C) DTI-FA (DV)** | | | | | | | | | |
| Fx (right) | -0.011 (-0.038 to 0.017) | 0.45 | 0.57 | 0.029 (-0.002 to 0.060) | 0.07 | 0.35 | 0.027 (-0.002 to 0.055) | 0.07 | 0.13 |
| Fx (left) | -0.024 (-0.051 to 0.003) | 0.09 | 0.22 | 0.008 (-0.023 to 0.039) | 0.60 | 0.93 | 0.042 (0.013 to 0.070) | 0.004 | **0.01** |
| CgC (right) | 0.028 (0.000 to 0.055) | 0.05 | 0.18 | -0.041 (-0.072 to 0.010) | 0.01 | 0.27 | 0.007 (-0.021 to 0.035) | 0.63 | 0.75 |
| CgC (left) | 0.017 (-0.010 to 0.044) | 0.22 | 0.43 | -0.028 (-0.059 to 0.002) | 0.07 | 0.35 | -0.013 (-0.041 to 0.015) | 0.37 | 0.50 |
| CgH (right) | -0.015 (-0.042 to 0.012) | 0.28 | 0.48 | -0.001 (-0.031 to 0.030) | 0.96 | 0.96 | 0.011 (-0.017 to 0.039) | 0.45 | 0.57 |
| CgH (left) | -0.010 (-0.037 to 0.017) | 0.47 | 0.57 | 0.021 (-0.009 to 0.051) | 0.17 | 0.67 | 0.028 (0.000 to 0.057) | 0.05 | 0.11 |
| CST (right) | 0.012 (-0.014 to 0.039) | 0.36 | 0.56 | -0.003 (-0.033 to 0.027) | 0.86 | 0.96 | 0.045 (0.018 to 0.073) | 0.001 | **0.01** |
| CST (left) | 0.031 (0.005 to 0.058) | 0.02 | 0.10 | -0.001 (-0.032 to 0.029) | 0.92 | 0.96 | 0.047 (0.019 to 0.075) | 0.001 | **0.01** |
| ATR (right) | 0.025 (-0.002 to 0.052) | 0.07 | 0.22 | 0.013 (-0.018 to 0.044) | 0.43 | 0.79 | 0.021 (-0.007 to 0.050) | 0.15 | 0.24 |
| ATR (left) | 0.005 (-0.023 to 0.033) | 0.74 | 0.79 | 0.030 (-0.001 to 0.062) | 0.06 | 0.35 | 0.021 (-0.008 to 0.049) | 0.15 | 0.24 |
| Unc (right) | 0.018 (-0.010 to 0.046) | 0.21 | 0.43 | -0.035 (-0.066 to 0.005) | 0.02 | 0.35 | 0.002 (-0.026 to 0.030) | 0.88 | 0.88 |
| Unc (left) | 0.012 (-0.016 to 0.040) | 0.42 | 0.57 | -0.020 (-0.051 to 0.011) | 0.21 | 0.69 | 0.006 (-0.022 to 0.035) | 0.67 | 0.77 |
| ILF (right) | -0.001 (-0.028 to 0.026) | 0.93 | 0.93 | -0.004 (-0.035 to 0.026) | 0.78 | 0.96 | 0.019 (-0.009 to 0.047) | 0.17 | 0.24 |
| ILF (left) | 0.012 (-0.016 to 0.039) | 0.40 | 0.57 | 0.009 (-0.022 to 0.040) | 0.57 | 0.93 | 0.020 (-0.008 to 0.048) | 0.16 | 0.24 |
| IFOF (right) | 0.005 (-0.022 to 0.032) | 0.73 | 0.79 | -0.025 (-0.056 to 0.005) | 0.10 | 0.44 | 0.031 (0.003 to 0.059) | 0.03 | 0.07 |
| IFOF (left) | 0.010 (-0.017 to 0.037) | 0.48 | 0.57 | -0.007 (-0.037 to 0.023) | 0.63 | 0.93 | 0.028 (0.000 to 0.056) | 0.05 | 0.11 |
| Fmaj | -0.024 (-0.052 to 0.003) | 0.08 | 0.22 | 0.013 (-0.017 to 0.044) | 0.40 | 0.79 | 0.007 (-0.021 to 0.035) | 0.63 | 0.75 |
| Fmin | 0.014 (-0.015 to 0.042) | 0.35 | 0.56 | -0.015 (-0.046 to 0.016) | 0.34 | 0.79 | -0.023 (-0.052 to 0.006) | 0.11 | 0.22 |
| CC | -0.011 (-0.038 to 0.017) | 0.45 | 0.57 | -0.003 (-0.034 to 0.027) | 0.84 | 0.96 | -0.003 (-0.031 to 0.025) | 0.83 | 0.88 |
| SLF (right) | -0.037 (-0.065 to -0.009) | 0.01 | 0.06 | -0.002 (-0.033 to 0.030) | 0.91 | 0.96 | 0.050 (0.021 to 0.078) | 0.001 | **0.005** |
| SLF (left) | -0.066 (-0.094 to -0.039) | < 0.001 | **< 0.001** | 0.015 (-0.016 to 0.046) | 0.34 | 0.79 | 0.050 (0.021 to 0.078) | 0.001 | **0.005** |
| tSLF (right) | -0.035 (-0.063 to -0.007) | 0.01 | 0.07 | -0.006 (-0.038 to 0.025) | 0.69 | 0.93 | 0.042 (0.014 to 0.071) | 0.004 | **0.01** |
| tSLF (left) | -0.065 (-0.093 to -0.038) | < 0.001 | **< 0.001** | 0.019 (-0.012 to 0.050) | 0.22 | 0.69 | 0.042 (0.014 to 0.071) | 0.003 | **0.01** |
| pSLF (right) | -0.037 (-0.064 to -0.009) | 0.01 | 0.06 | 0.001 (-0.030 to 0.032) | 0.94 | 0.96 | 0.050 (0.021 to 0.078) | 0.001 | **0.005** |
| pSLF (left) | -0.056 (-0.083 to -0.028) | < 0.001 | **0.001** | 0.006 (-0.024 to 0.037) | 0.69 | 0.93 | 0.058 (0.030 to 0.087) | < 0.001 | **0.002** |
| SCS (right) | 0.020 (-0.007 to 0.047) | 0.15 | 0.33 | -0.015 (-0.045 to 0.016) | 0.34 | 0.79 | 0.033 (0.004 to 0.061) | 0.02 | 0.07 |
| SCS (left) | 0.006 (-0.012 to 0.044) | 0.27 | 0.75 | 0.003 (-0.027 to 0.033) | 0.85 | 0.96 | 0.039 (0.011 to 0.067) | 0.01 | **0.02** |
| SIFC (right) | 0.016 (-0.012 to 0.044) | 0.27 | 0.48 | -0.032 (-0.063 to -0.002) | 0.04 | 0.35 | 0.002 (-0.026 to 0.031) | 0.87 | 0.88 |
| SIFC (left) | -0.002 (-0.030 to 0.026) | 0.89 | 0.92 | -0.012 (-0.042 to 0.019) | 0.46 | 0.79 | -0.004 (-0.033 to 0.024) | 0.77 | 0.86 |
| IFSFC (right) | -0.023 (-0.050 to 0.005) | 0.10 | 0.25 | 0.014 (-0.017 to 0.044) | 0.39 | 0.79 | 0.022 (-0.007 to 0.050) | 0.13 | 0.24 |
| IFSFC (left) | -0.027 (-0.054 to 0.001) | 0.06 | 0.20 | 0.012 (-0.019 to 0.043) | 0.45 | 0.79 | 0.020 (-0.008 to 0.048) | 0.17 | 0.24 |

**(continued) eTable 8. Associations between SES and white matter microstructure**

| **White matter tracts** | **SES indicators (IVs)** | | | | | | | | |
| --- | --- | --- | --- | --- | --- | --- | --- | --- | --- |
|  | **Neighborhood disadvantage**  (higher = lower SES) | | | **Household income**  (higher = higher SES) | | | **Parental education**  (higher = higher SES) | | |
|  | ***β* (95% CI)** | ***p*-value**  (nominal) | ***p*-value**  (FDR) | ***β* (95% CI)** | ***p*-value**  (nominal) | ***p*-value**  (FDR) | ***β* (95% CI)** | ***p*-value**  (nominal) | ***p*-value**  (FDR) |
| **D) DTI-MD (DV)** | | | | | | | | | |
| Fx (right) | 0.003 (-0.024 to 0.030) | 0.82 | 0.82 | -0.027 (-0.058 to 0.004) | 0.09 | 0.24 | 0.009 (-0.019 to 0.038) | 0.52 | 0.76 |
| Fx (left) | -0.003 (-0.031 to 0.024) | 0.81 | 0.82 | -0.010 (-0.041 to 0.021) | 0.54 | 0.59 | 0.008 (-0.021 to 0.036) | 0.60 | 0.78 |
| CgC (right) | -0.013 (-0.041 to 0.014) | 0.35 | 0.51 | 0.009 (-0.022 to 0.041) | 0.56 | 0.59 | 0.011 (-0.017 to 0.040) | 0.44 | 0.76 |
| CgC (left) | -0.012 (-0.039 to 0.016) | 0.40 | 0.54 | 0.020 (-0.011 to 0.050) | 0.20 | 0.32 | 0.003 (-0.026 to 0.031) | 0.85 | 0.91 |
| CgH (right) | -0.016 (-0.044 to 0.012) | 0.25 | 0.41 | 0.020 (-0.011 to 0.051) | 0.21 | 0.32 | -0.008 (-0.037 to 0.020) | 0.56 | 0.76 |
| CgH (left) | -0.020 (-0.047 to 0.007) | 0.15 | 0.33 | 0.039 (-0.008 to 0.069) | 0.01 | 0.13 | -0.004 (-0.032 to 0.024) | 0.78 | 0.91 |
| CST (right) | -0.022 (-0.050 to 0.005) | 0.11 | 0.32 | 0.035 (-0.004 to 0.066) | 0.03 | 0.13 | -0.007 (-0.035 to 0.022) | 0.65 | 0.81 |
| CST (left) | -0.033 (-0.061 to -0.004) | 0.02 | 0.12 | 0.029 (-0.002 to 0.061) | 0.07 | 0.23 | 0.002 (-0.026 to 0.031) | 0.88 | 0.91 |
| ATR (right) | -0.012 (-0.040 to 0.017) | 0.42 | 0.54 | 0.040 (0.009 to 0.071) | 0.01 | 0.13 | -0.014 (-0.042 to 0.014) | 0.33 | 0.73 |
| ATR (left) | -0.008 (-0.035 to 0.019) | 0.56 | 0.67 | 0.028 (-0.003 to 0.058) | 0.08 | 0.23 | 0.004 (-0.024 to 0.032) | 0.79 | 0.91 |
| Unc (right) | 0.016 (-0.012 to 0.043) | 0.27 | 0.42 | 0.037 (0.005 to 0.068) | 0.02 | 0.13 | -0.009 (-0.037 to 0.020) | 0.54 | 0.76 |
| Unc (left) | 0.004 (-0.024 to 0.032) | 0.78 | 0.82 | 0.027 (-0.005 to 0.058) | 0.09 | 0.24 | 0.001 (-0.027 to 0.030) | 0.92 | 0.92 |
| ILF (right) | 0.022 (-0.005 to 0.049) | 0.10 | 0.32 | 0.018 (-0.013 to 0.048) | 0.25 | 0.33 | -0.016 (-0.044 to 0.012) | 0.27 | 0.73 |
| ILF (left) | 0.020 (-0.006 to 0.047) | 0.14 | 0.32 | 0.009 (-0.022 to 0.040) | 0.57 | 0.59 | -0.016 (-0.044 to 0.012) | 0.26 | 0.73 |
| IFOF (right) | 0.026 (-0.001 to 0.053) | 0.06 | 0.20 | 0.036 (0.006 to 0.066) | 0.02 | 0.13 | -0.011 (-0.039 to 0.016) | 0.42 | 0.76 |
| IFOF (left) | 0.019 (-0.008 to 0.046) | 0.17 | 0.33 | 0.031 (0.001 to 0.062) | 0.04 | 0.20 | -0.009 (-0.037 to 0.018) | 0.50 | 0.76 |
| Fmaj | 0.019 (-0.009 to 0.047) | 0.18 | 0.33 | -0.016 (-0.047 to 0.015) | 0.31 | 0.37 | 0.039 (0.011 to 0.068) | 0.01 | 0.23 |
| Fmin | -0.004 (-0.032 to 0.024) | 0.79 | 0.82 | 0.010 (-0.021 to 0.042) | 0.52 | 0.59 | 0.031 (0.002 to 0.060) | 0.04 | 0.51 |
| CC | 0.021 (-0.006 to 0.049) | 0.13 | 0.32 | 0.006 (-0.025 to 0.037) | 0.71 | 0.71 | 0.021 (-0.007 to 0.050) | 0.14 | 0.71 |
| SLF (right) | 0.034 (0.007 to 0.061) | 0.01 | 0.12 | 0.023 (-0.007 to 0.054) | 0.13 | 0.30 | -0.025 (-0.053 to 0.003) | 0.09 | 0.65 |
| SLF (left) | 0.034 (0.006 to 0.062) | 0.02 | 0.12 | 0.018 (-0.013 to 0.049) | 0.25 | 0.33 | -0.016 (-0.044 to 0.012) | 0.27 | 0.73 |
| tSLF (right) | 0.029 (0.002 to 0.056) | 0.04 | 0.16 | 0.029 (-0.002 to 0.060) | 0.06 | 0.23 | -0.028 (-0.056 to 0.000) | 0.05 | 0.51 |
| tSLF (left) | 0.032 (0.004 to 0.060) | 0.02 | 0.12 | 0.019 (-0.012 to 0.050) | 0.23 | 0.33 | -0.016 (-0.044 to 0.012) | 0.27 | 0.73 |
| pSLF (right) | 0.035 (0.008 to 0.063) | 0.01 | 0.12 | 0.023 (-0.008 to 0.054) | 0.15 | 0.30 | -0.023 (-0.051 to 0.005) | 0.11 | 0.65 |
| pSLF (left) | 0.036 (0.008 to 0.064) | 0.01 | 0.12 | 0.020 (-0.011 to 0.051) | 0.21 | 0.32 | -0.015 (-0.043 to 0.013) | 0.30 | 0.73 |
| SCS (right) | 0.027 (0.000 to 0.055) | 0.05 | 0.19 | 0.024 (-0.007 to 0.055) | 0.13 | 0.30 | -0.015 (-0.043 to 0.013) | 0.29 | 0.73 |
| SCS (left) | 0.008 (-0.019 to 0.036) | 0.56 | 0.67 | 0.019 (-0.011 to 0.050) | 0.22 | 0.32 | -0.010 (-0.038 to 0.019) | 0.50 | 0.76 |
| SIFC (right) | 0.012 (-0.016 to 0.040) | 0.39 | 0.54 | 0.037 (0.007 to 0.068) | 0.02 | 0.13 | -0.015 (-0.043 to 0.014) | 0.31 | 0.73 |
| SIFC (left) | -0.004 (-0.032 to 0.024) | 0.78 | 0.82 | 0.020 (-0.011 to 0.051) | 0.21 | 0.32 | 0.012 (-0.017 to 0.040) | 0.41 | 0.76 |
| IFSFC (right) | 0.018 (-0.009 to 0.046) | 0.19 | 0.33 | 0.018 (-0.013 to 0.049) | 0.26 | 0.33 | -0.012 (-0.041 to 0.016) | 0.39 | 0.76 |
| IFSFC (left) | 0.019 (-0.008 to 0.047) | 0.17 | 0.33 | 0.021 (-0.010 to 0.052) | 0.19 | 0.32 | -0.002 (-0.031 to 0.026) | 0.88 | 0.91 |

*Note*. Linear mixed effects models included all three SES indicators simultaneously as independent variables (IVs) and RSI and DTI metrics in each of the white matter tracts as the dependent variable (DV). Models were adjusted for participant age, sex, PDS, ICV, and mean head motion, and were nested by family. Multiple comparison correction was conducted within each neuroimaging metric and by each SES indicator, giving 31 tests to correct for in each group of models. Effects that survived false discovery rate (FDR)-corrected *p*-value ≤ 0.05 were considered statistically significant and are highlighted. Estimates were standardized *β*’s with 95% confidence intervals (CIs). RSI, restriction spectrum imaging; RND, restricted normalized directional; RNI, restricted normalized isotropic; DTI, diffusion tensor imaging; FA, fractional anisotropy; MD, mean diffusivity. Full names of white matter tracts are shown in the main text.

**eTable 9. Sensitivity analyses on associations between SES and white matter microstructure**

**1) Using only scans with low head motion (mean motion ≤ 2.5 mm)**

| **White matter tracts** | **SES indicators (IVs)** | | | | | | **Sample size (*n*)** |
| --- | --- | --- | --- | --- | --- | --- | --- |
|  | **Neighborhood disadvantage**  (higher = lower SES) | | **Household income**  (higher = higher SES) | | **Parental education**  (higher = higher SES) | |  |
|  | ***β* (95% CI)** | ***p*-value**  (nominal) | ***β* (95% CI)** | ***p*-value**  (nominal) | ***β* (95% CI)** | ***p*-value**  (nominal) |  |
| **A) RSI-RND (DV)** | | | | | | | |
| CST (right) | 0.010 (-0.017 to 0.037) | 0.46 | 0.014 (-0.016 to 0.044) | 0.36 | 0.041 (0.013 to 0.069) | 0.005 | 8618 |
| CST (left) | 0.026 (-0.002 to 0.053) | 0.06 | 0.008 (-0.022 to 0.039) | 0.60 | 0.046 (0.018 to 0.075) | 0.001 | 8616 |
| Fmaj | -0.043 (-0.071 to -0.015) | 0.003 | 0.022 (-0.009 to 0.054) | 0.16 | -0.012 (-0.041 to 0.017) | 0.42 | 8612 |
| SLF (right) | -0.021 (-0.049 to 0.006) | 0.13 | -0.003 (-0.034 to 0.029) | 0.87 | 0.053 (0.024 to 0.082) | < 0.001 | 8623 |
| SLF (left) | -0.064 (-0.091 to -0.036) | < 0.001 | 0.013 (-0.018 to 0.044) | 0.41 | 0.048 (0.020 to 0.077) | 0.001 | 8619 |
| tSLF (right) | -0.021 (-0.049 to 0.006) | 0.13 | -0.003 (-0.034 to 0.028) | 0.85 | 0.045 (0.016 to 0.074) | 0.002 | 8624 |
| tSLF (left) | -0.065 (-0.093 to -0.038) | < 0.001 | 0.018 (-0.013 to 0.049) | 0.26 | 0.040 (0.011 to 0.068) | 0.01 | 8622 |
| pSLF (right) | -0.023 (-0.050 to 0.005) | 0.10 | -0.003 (-0.034 to 0.029) | 0.87 | 0.054 (0.025 to 0.082) | < 0.001 | 8621 |
| pSLF (left) | -0.048 (-0.075 to -0.020) | 0.001 | 0.001 (-0.030 to 0.032) | 0.96 | 0.058 (0.030 to 0.087) | < 0.001 | 8614 |
| **B) RSI-RNI (DV)** | | | | | | | |
| Fx (right) | 0.047 (0.020 to 0.075) | 0.001 | 0.010 (-0.021 to 0.041) | 0.53 | -0.016 (-0.045 to 0.013) | 0.29 | 8601 |
| Fx (left) | 0.049 (0.021 to 0.076) | 0.001 | -0.001 (-0.033 to 0.030) | 0.93 | -0.016 (-0.045 to 0.013) | 0.29 | 8584 |
| CgC (right) | 0.027 (0.000 to 0.054) | 0.05 | -0.034 (-0.065 to -0.004) | 0.03 | -0.014 (-0.042 to 0.015) | 0.35 | 8605 |
| CgC (left) | 0.018 (-0.010 to 0.045) | 0.21 | -0.034 (-0.065 to -0.003) | 0.03 | -0.019 (-0.048 to 0.010) | 0.20 | 8594 |
| CgH (right) | 0.061 (0.033 to 0.088) | < 0.001 | -0.032 (-0.062 to -0.001) | 0.03 | 0.004 (-0.024 to 0.032) | 0.78 | 8608 |
| CgH (left) | 0.065 (0.038 to 0.093) | < 0.001 | -0.051 (-0.082 to -0.021) | 0.001 | -0.005 (-0.034 to 0.023) | 0.71 | 8590 |
| CST (right) | 0.038 (0.010 to 0.065) | 0.01 | -0.032 (-0.063 to -0.001) | 0.05 | -0.002 (-0.031 to 0.026) | 0.87 | 8615 |
| CST (left) | 0.041 (0.014 to 0.068) | 0.003 | -0.045 (-0.076 to -0.013) | 0.01 | -0.004 (-0.032 to 0.025) | 0.79 | 8607 |
| ATR (right) | 0.047 (0.020 to 0.074) | 0.001 | -0.046 (-0.077 to -0.015) | 0.003 | 0.004 (-0.024 to 0.033) | 0.76 | 8579 |
| ATR (left) | 0.040 (0.012 to 0.067) | 0.005 | -0.038 (-0.070 to -0.007) | 0.02 | -0.013 (-0.041 to 0.016) | 0.38 | 8576 |
| Unc (right) | 0.021 (-0.006 to 0.049) | 0.12 | -0.062 (-0.093 to -0.032) | < 0.001 | 0.002 (-0.026 to 0.031) | 0.87 | 8616 |
| Unc (left) | 0.012 (-0.015 to 0.040) | 0.38 | -0.056 (-0.087 to -0.025) | < 0.001 | -0.005 (-0.034 to 0.024) | 0.74 | 8615 |
| ILF (right) | -0.007 (-0.034 to 0.019) | 0.59 | -0.044 (-0.075 to -0.014) | 0.005 | -0.004 (-0.032 to 0.024) | 0.79 | 8623 |
| ILF (left) | -0.005 (-0.032 to 0.022) | 0.74 | -0.034 (-0.065 to -0.003) | 0.03 | -0.015 (-0.043 to 0.013) | 0.30 | 8618 |
| IFOF (right) | 0.001 (-0.025 to 0.028) | 0.92 | -0.055 (-0.085 to -0.025) | < 0.001 | -0.009 (-0.037 to 0.019) | 0.52 | 8619 |
| IFOF (left) | 0.000 (-0.026 to 0.027) | 0.97 | -0.051 (-0.082 to -0.020) | 0.001 | -0.014 (-0.042 to 0.014) | 0.33 | 8591 |
| Fmaj | 0.001 (-0.027 to 0.028) | 0.96 | -0.005 (-0.036 to 0.026) | 0.74 | -0.051 (-0.080 to -0.023) | < 0.001 | 8588 |
| Fmin | 0.013 (-0.014 to 0.041) | 0.34 | -0.045 (-0.076 to 0.013) | 0.01 | -0.039 (-0.068 to -0.010) | 0.01 | 8608 |
| CC | 0.002 (-0.025 to 0.030) | 0.87 | -0.037 (-0.068 to -0.005) | 0.02 | -0.027 (-0.055 to 0.002) | 0.07 | 8609 |
| SLF (right) | -0.005 (-0.031 to 0.022) | 0.73 | -0.031 (-0.061 to -0.001) | 0.05 | -0.007 (-0.035 to 0.021) | 0.63 | 8624 |
| SLF (left) | 0.011 (-0.016 to 0.038) | 0.42 | -0.046 (-0.077 to -0.016) | 0.003 | -0.003 (-0.031 to 0.025) | 0.83 | 8613 |
| tSLF (left) | 0.010 (-0.017 to 0.037) | 0.46 | -0.045 (-0.076 to -0.015) | 0.004 | -0.001 (-0.029 to 0.028) | 0.97 | 8610 |
| pSLF (right) | -0.003 (-0.029 to 0.024) | 0.85 | -0.033 (-0.065 to -0.003) | 0.03 | -0.006 (-0.034 to 0.022) | 0.67 | 8625 |
| pSLF (left) | 0.011 (-0.016 to 0.038) | 0.42 | -0.047 (-0.078 to -0.017) | 0.003 | -0.001 (-0.030 to 0.027) | 0.93 | 8617 |
| SCS (left) | 0.001 (-0.025 to 0.028) | 0.93 | -0.038 (-0.069 to -0.008) | 0.01 | 0.003 (-0.025 to 0.031) | 0.83 | 8614 |
| SIFC (right) | -0.006 (-0.033 to 0.022) | 0.69 | -0.043 (-0.074 to -0.012) | 0.01 | -0.008 (-0.037 to 0.021) | 0.58 | 8618 |
| SIFC (left) | 0.001 (-0.027 to 0.028) | 0.96 | -0.038 (-0.069 to -0.006) | 0.02 | -0.012 (-0.040 to 0.017) | 0.43 | 8619 |
| IFSFC (right) | 0.001 (-0.026 to 0.027) | 0.97 | -0.047 (-0.078 to -0.016) | 0.003 | 0.006 (-0.022 to 0.035) | 0.66 | 8622 |
| IFSFC (left) | -0.006 (-0.033 to 0.021) | 0.66 | -0.047 (-0.078 to -0.016) | 0.003 | 0.007 (-0.021 to 0.036) | 0.61 | 8624 |
| **C) DTI-FA (DV)** | | | | | | | |
| Fx (left) | -0.028 (-0.055 to 0.000) | 0.05 | 0.013 (-0.018 to 0.044) | 0.42 | 0.037 (0.008 to 0.066) | 0.01 | 8630 |
| CST (right) | 0.010 (-0.017 to 0.037) | 0.49 | -0.003 (-0.033 to 0.028) | 0.85 | 0.045 (0.017 to 0.074) | 0.002 | 8626 |
| CST (left) | 0.026 (-0.001 to 0.053) | 0.06 | -0.002 (-0.032 to 0.029) | 0.92 | 0.044 (0.015 to 0.072) | 0.003 | 8627 |
| SLF (right) | -0.042 (-0.070 to -0.014) | 0.004 | -0.004 (-0.036 to 0.028) | 0.80 | 0.051 (0.022 to 0.080) | 0.001 | 8628 |
| SLF (left) | -0.074 (-0.102 to -0.046) | < 0.001 | 0.014 (-0.017 to 0.045) | 0.38 | 0.048 (0.019 to 0.077) | 0.001 | 8627 |
| tSLF (right) | -0.038 (-0.066 to -0.010) | 0.01 | -0.009 (-0.041 to 0.023) | 0.58 | 0.043 (0.014 to 0.072) | 0.004 | 8629 |
| tSLF (left) | -0.072 (-0.100 to -0.044) | < 0.001 | 0.019 (-0.013 to 0.050) | 0.24 | 0.040 (0.012 to 0.069) | 0.01 | 8628 |
| pSLF (right) | -0.042 (-0.070 to -0.014) | 0.004 | -0.001 (-0.033 to 0.031) | 0.95 | 0.050 (0.021 to 0.079) | 0.001 | 8628 |
| pSLF (left) | -0.063 (-0.091 to -0.035) | < 0.001 | 0.005 (-0.027 to 0.036) | 0.77 | 0.056 (0.028 to 0.085) | < 0.001 | 8624 |
| SCS (left) | 0.002 (-0.025 to 0.029) | 0.90 | 0.001 (-0.029 to 0.032) | 0.93 | 0.032 (0.004 to 0.060) | 0.03 | 8629 |

**2) Using only participants without reported adverse childhood experiences (ACEs)**

| **White matter tracts** | **SES indicators (IVs)** | | | | | | **Sample size (*n*)** |
| --- | --- | --- | --- | --- | --- | --- | --- |
|  | **Neighborhood disadvantage**  (higher = lower SES) | | **Household income**  (higher = higher SES) | | **Parental education**  (higher = higher SES) | |  |
|  | ***β* (95% CI)** | ***p*-value**  (nominal) | ***β* (95% CI)** | ***p*-value**  (nominal) | ***β* (95% CI)** | ***p*-value**  (nominal) |  |
| **A) RSI-RND (DV)** | | | | | | | |
| CST (right) | -0.002 (-0.035 to 0.031) | 0.90 | -0.001 (-0.039 to 0.036) | 0.95 | 0.034 (-0.001 to 0.068) | 0.06 | 5670 |
| CST (left) | 0.019 (-0.015 to 0.054) | 0.26 | 0.002 (-0.036 to 0.040) | 0.92 | 0.037 (0.003 to 0.072) | 0.04 | 5668 |
| Fmaj | -0.023 (-0.057 to 0.012) | 0.19 | 0.032 (-0.007 to 0.070) | 0.11 | -0.003 (-0.039 to 0.032) | 0.85 | 5669 |
| SLF (right) | -0.010 (-0.044 to 0.023) | 0.54 | -0.010 (-0.048 to 0.027) | 0.59 | 0.062 (0.027 to 0.097) | < 0.001 | 5674 |
| SLF (left) | -0.051 (-0.084 to -0.018) | 0.003 | 0.011 (-0.026 to 0.049) | 0.55 | 0.053 (0.018 to 0.087) | 0.003 | 5671 |
| tSLF (right) | -0.017 (-0.051 to 0.016) | 0.31 | -0.016 (-0.054 to 0.022) | 0.40 | 0.056 (0.020 to 0.091) | 0.002 | 5675 |
| tSLF (left) | -0.053 (-0.086 to -0.019) | 0.002 | 0.014 (-0.023 to 0.052) | 0.46 | 0.045 (0.010 to 0.079) | 0.01 | 5673 |
| pSLF (right) | -0.010 (-0.043 to 0.023) | 0.54 | -0.008 (-0.046 to 0.029) | 0.66 | 0.063 (0.028 to 0.097) | < 0.001 | 5674 |
| pSLF (left) | -0.035 (-0.068 to -0.002) | 0.04 | 0.002 (-0.035 to 0.039) | 0.91 | 0.062 (0.028 to 0.096) | < 0.001 | 5669 |
| **B) RSI-RNI (DV)** | | | | | | | |
| Fx (right) | 0.051 (0.016 to 0.085) | 0.004 | 0.003 (-0.036 to 0.042) | 0.88 | -0.004 (-0.040 to 0.032) | 0.81 | 5663 |
| Fx (left) | 0.048 (0.013 to 0.082) | 0.01 | -0.015 (-0.054 to 0.024) | 0.46 | -0.007 (-0.043 to 0.029) | 0.69 | 5654 |
| CgC (right) | 0.046 (0.012 to 0.080) | 0.01 | -0.028 (-0.066 to 0.010) | 0.14 | -0.010 (-0.045 to 0.026) | 0.59 | 5669 |
| CgC (left) | 0.035 (0.001 to 0.069) | 0.05 | -0.025 (-0.064 to 0.014) | 0.21 | -0.010 (-0.046 to 0.026) | 0.60 | 5661 |
| CgH (right) | 0.077 (0.043 to 0.110) | < 0.001 | -0.012 (-0.050 to 0.025) | 0.52 | 0.003 (-0.033 to 0.038) | 0.88 | 5669 |
| CgH (left) | 0.070 (0.036 to 0.104) | < 0.001 | -0.029 (-0.066 to 0.009) | 0.14 | -0.014 (-0.049 to 0.021) | 0.43 | 5654 |
| CST (right) | 0.057 (0.023 to 0.091) | 0.001 | -0.017 (-0.055 to 0.021) | 0.38 | 0.012 (-0.024 to 0.047) | 0.51 | 5672 |
| CST (left) | 0.056 (0.022 to 0.090) | 0.001 | -0.033 (-0.071 to 0.005) | 0.09 | 0.009 (-0.026 to 0.044) | 0.61 | 5670 |
| ATR (right) | 0.052 (0.018 to 0.085) | 0.003 | -0.051 (-0.089 to -0.013) | 0.01 | 0.010 (-0.025 to 0.046) | 0.57 | 5647 |
| ATR (left) | 0.051 (0.017 to 0.085) | 0.003 | -0.042 (-0.080 to -0.003) | 0.03 | -0.011 (-0.047 to 0.024) | 0.53 | 5648 |
| Unc (right) | 0.030 (-0.003 to 0.064) | 0.08 | -0.046 (-0.085 to -0.008) | 0.02 | -0.003 (-0.038 to 0.033) | 0.89 | 5672 |
| Unc (left) | 0.028 (-0.025 to 0.062) | 0.10 | -0.037 (-0.076 to 0.001) | 0.06 | -0.013 (-0.049 to 0.022) | 0.46 | 5670 |
| ILF (right) | 0.006 (-0.028 to 0.039) | 0.74 | -0.031 (-0.069 to 0.008) | 0.12 | -0.007 (-0.042 to 0.028) | 0.69 | 5677 |
| ILF (left) | 0.011 (-0.022 to 0.044) | 0.51 | -0.021 (-0.060 to 0.017) | 0.28 | -0.019 (-0.054 to 0.017) | 0.30 | 5673 |
| IFOF (right) | 0.001 (-0.032 to 0.034) | 0.95 | -0.055 (-0.093 to -0.017) | 0.004 | -0.010 (-0.045 to 0.025) | 0.59 | 5675 |
| IFOF (left) | 0.010 (-0.023 to 0.043) | 0.56 | -0.049 (-0.087 to -0.011) | 0.01 | -0.013 (-0.049 to 0.022) | 0.46 | 5655 |
| Fmaj | 0.008 (-0.027 to 0.042) | 0.67 | -0.011 (-0.050 to 0.027) | 0.57 | -0.040 (-0.075 to -0.004) | 0.03 | 5653 |
| Fmin | 0.028 (-0.006 to 0.063) | 0.10 | -0.046 (-0.085 to -0.007) | 0.02 | -0.040 (-0.076 to -0.004) | 0.03 | 5666 |
| CC | 0.017 (-0.017 to 0.051) | 0.32 | -0.036 (-0.075 to 0.003) | 0.07 | -0.025 (-0.061 to 0.011) | 0.17 | 5666 |
| SLF (right) | 0.002 (-0.031 to 0.035) | 0.89 | -0.021 (-0.058 to 0.017) | 0.28 | -0.014 (-0.049 to 0.020) | 0.42 | 5676 |
| SLF (left) | 0.028 (-0.006 to 0.063) | 0.10 | -0.032 (-0.070 to 0.006) | 0.10 | -0.004 (-0.039 to 0.031) | 0.81 | 5671 |
| tSLF (left) | 0.026 (-0.007 to 0.060) | 0.12 | -0.029 (-0.067 to 0.009) | 0.13 | -0.002 (-0.037 to 0.033) | 0.93 | 5670 |
| pSLF (right) | 0.004 (-0.029 to 0.037) | 0.79 | -0.024 (-0.062 to 0.013) | 0.20 | -0.015 (-0.050 to 0.020) | 0.40 | 5677 |
| pSLF (left) | 0.028 (-0.006 to 0.061) | 0.10 | -0.037 (-0.075 to 0.001) | 0.06 | -0.004 (-0.039 to 0.031) | 0.83 | 5673 |
| SCS (left) | 0.011 (-0.022 to 0.044) | 0.51 | -0.027 (-0.065 to 0.010) | 0.15 | -0.001 (-0.035 to 0.034) | 0.97 | 5669 |
| SIFC (right) | 0.000 (-0.034 to 0.034) | 0.99 | -0.034 (-0.072 to 0.004) | 0.08 | -0.016 (-0.052 to 0.020) | 0.38 | 5672 |
| SIFC (left) | 0.020 (-0.014 to 0.053) | 0.25 | -0.023 (-0.061 to 0.016) | 0.25 | -0.019 (-0.055 to 0.016) | 0.29 | 5672 |
| IFSFC (right) | 0.013 (-0.020 to 0.046) | 0.44 | -0.036 (-0.074 to 0.002) | 0.06 | 0.004 (-0.031 to 0.039) | 0.83 | 5675 |
| IFSFC (left) | 0.014 (-0.019 to 0.048) | 0.40 | -0.036 (-0.074 to 0.002) | 0.06 | 0.007 (-0.028 to 0.043) | 0.68 | 5676 |
| **C) DTI-FA (DV)** | | | | | | | |
| Fx (left) | -0.011 (-0.046 to 0.023) | 0.51 | -0.003 (-0.041 to 0.036) | 0.89 | 0.055 (0.019 to 0.090) | 0.003 | 5681 |
| CST (right) | 0.002 (-0.032 to 0.035) | 0.92 | -0.014 (-0.052 to 0.024) | 0.47 | 0.041 (0.006 to 0.076) | 0.02 | 5679 |
| CST (left) | 0.029 (-0.005 to 0.064) | 0.10 | -0.004 (-0.043 to 0.034) | 0.83 | 0.040 (0.005 to 0.075) | 0.02 | 5678 |
| SLF (right) | -0.033 (-0.067 to 0.002) | 0.06 | -0.006 (-0.045 to 0.033) | 0.77 | 0.059 (0.023 to 0.095) | 0.001 | 5680 |
| SLF (left) | -0.064 (-0.099 to -0.030) | < 0.001 | 0.016 (-0.023 to 0.055) | 0.42 | 0.047 (0.012 to 0.082) | 0.01 | 5680 |
| tSLF (right) | -0.035 (-0.069 to -0.001) | 0.05 | -0.018 (-0.057 to 0.021) | 0.36 | 0.051 (0.016 to 0.087) | 0.005 | 5680 |
| tSLF (left) | -0.064 (-0.099 to -0.030) | < 0.001 | 0.017 (-0.022 to 0.055) | 0.40 | 0.040 (0.005 to 0.075) | 0.03 | 5681 |
| pSLF (right) | -0.031 (-0.065 to 0.003) | 0.08 | 0.000 (-0.038 to 0.039) | 0.98 | 0.059 (0.023 to 0.094) | 0.001 | 5680 |
| pSLF (left) | -0.052 (-0.086 to -0.017) | 0.003 | 0.013 (-0.025 to 0.052) | 0.50 | 0.054 (0.019 to 0.090) | 0.003 | 5680 |
| SCS (left) | 0.019 (-0.015 to 0.053) | 0.28 | 0.009 (-0.029 to 0.047) | 0.65 | 0.028 (-0.007 to 0.063) | 0.12 | 5680 |

**3) Using only participants without common psychiatric diagnoses**

| **White matter tracts** | **SES indicators (IVs)** | | | | | | **Sample size (*n*)** |
| --- | --- | --- | --- | --- | --- | --- | --- |
|  | **Neighborhood disadvantage**  (higher = lower SES) | | **Household income**  (higher = higher SES) | | **Parental education**  (higher = higher SES) | |  |
|  | ***β* (95% CI)** | ***p*-value**  (nominal) | ***β* (95% CI)** | ***p*-value**  (nominal) | ***β* (95% CI)** | ***p*-value**  (nominal) |  |
| **A) RSI-RND (DV)** | | | | | | | |
| CST (right) | 0.006 (-0.022 to 0.034) | 0.66 | 0.010 (-0.021 to 0.042) | 0.52 | 0.040 (0.010 to 0.069) | 0.01 | 7672 |
| CST (left) | 0.024 (-0.005 to 0.052) | 0.10 | 0.008 (-0.024 to 0.040) | 0.61 | 0.045 (0.015 to 0.074) | 0.003 | 7672 |
| Fmaj | -0.041 (-0.070 to -0.012) | 0.01 | 0.026 (-0.006 to 0.059) | 0.11 | -0.011 (-0.041 to 0.019) | 0.49 | 7669 |
| SLF (right) | -0.021 (-0.050 to 0.008) | 0.15 | -0.003 (-0.035 to 0.029) | 0.85 | 0.056 (0.026 to 0.086) | < 0.001 | 7677 |
| SLF (left) | -0.056 (-0.084 to -0.028) | < 0.001 | 0.012 (-0.019 to 0.044) | 0.45 | 0.052 (0.022 to 0.081) | 0.001 | 7671 |
| tSLF (right) | -0.018 (-0.047 to 0.011) | 0.21 | -0.004 (-0.036 to 0.028) | 0.81 | 0.053 (0.023 to 0.083) | 0.001 | 7676 |
| tSLF (left) | -0.055 (-0.083 to -0.026) | < 0.001 | 0.017 (-0.015 to 0.049) | 0.30 | 0.046 (0.016 to 0.075) | 0.002 | 7673 |
| pSLF (right) | -0.023 (-0.052 to 0.005) | 0.11 | -0.002 (-0.034 to 0.030) | 0.90 | 0.055 (0.026 to 0.084) | < 0.001 | 7673 |
| pSLF (left) | -0.044 (-0.072 to -0.016) | 0.002 | 0.000 (-0.031 to 0.031) | > 0.99 | 0.059 (0.029 to 0.088) | < 0.001 | 7667 |
| **B) RSI-RNI (DV)** | | | | | | | |
| Fx (right) | 0.054 (0.025 to 0.083) | < 0.001 | 0.005 (-0.028 to 0.038) | 0.75 | -0.001 (-0.031 to 0.030) | 0.96 | 7661 |
| Fx (left) | 0.051 (0.022 to 0.080) | 0.001 | -0.004 (-0.038 to 0.029) | 0.79 | -0.007 (-0.037 to 0.024) | 0.67 | 7643 |
| CgC (right) | 0.032 (0.003 to 0.061) | 0.03 | -0.036 (-0.069 to -0.004) | 0.03 | -0.013 (-0.043 to 0.017) | 0.39 | 7664 |
| CgC (left) | 0.022 (-0.007 to 0.051) | 0.14 | -0.032 (-0.065 to 0.000) | 0.05 | -0.025 (-0.055 to 0.006) | 0.11 | 7653 |
| CgH (right) | 0.061 (0.033 to 0.090) | < 0.001 | -0.035 (-0.067 to -0.003) | 0.03 | 0.008 (-0.022 to 0.038) | 0.61 | 7666 |
| CgH (left) | 0.068 (0.040 to 0.097) | < 0.001 | -0.052 (-0.084 to -0.019) | 0.002 | -0.003 (-0.032 to 0.027) | 0.87 | 7648 |
| CST (right) | 0.040 (0.011 to 0.069) | 0.01 | -0.027 (-0.060 to 0.006) | 0.11 | 0.001 (-0.029 to 0.031) | 0.94 | 7673 |
| CST (left) | 0.048 (0.019 to 0.076) | 0.001 | -0.041 (-0.073 to -0.008) | 0.02 | 0.004 (-0.026 to 0.034) | 0.79 | 7665 |
| ATR (right) | 0.048 (0.019 to 0.077) | 0.001 | -0.051 (-0.083 to -0.018) | 0.002 | 0.009 (-0.021 to 0.039) | 0.55 | 7639 |
| ATR (left) | 0.043 (0.014 to 0.072) | 0.003 | -0.040 (-0.073 to -0.007) | 0.02 | -0.010 (-0.041 to 0.020) | 0.50 | 7640 |
| Unc (right) | 0.027 (-0.002 to 0.056) | 0.07 | -0.064 (-0.097 to -0.032) | < 0.001 | 0.010 (-0.020 to 0.040) | 0.53 | 7674 |
| Unc (left) | 0.018 (-0.011 to 0.047) | 0.22 | -0.051 (-0.083 to -0.018) | 0.003 | -0.002 (-0.032 to 0.029) | 0.91 | 7674 |
| ILF (right) | -0.003 (-0.031 to 0.026) | 0.84 | -0.042 (-0.074 to -0.009) | 0.01 | -0.002 (-0.032 to 0.028) | 0.91 | 7680 |
| ILF (left) | -0.001 (-0.030 to 0.027) | 0.93 | -0.025 (-0.058 to 0.007) | 0.13 | -0.015 (-0.045 to 0.015) | 0.32 | 7678 |
| IFOF (right) | 0.005 (-0.023 to 0.034) | 0.71 | -0.055 (-0.087 to -0.023) | 0.001 | -0.003 (-0.032 to 0.027) | 0.86 | 7677 |
| IFOF (left) | 0.006 (-0.022 to 0.034) | 0.68 | -0.047 (-0.080 to -0.015) | 0.004 | -0.008 (-0.038 to 0.022) | 0.60 | 7652 |
| Fmaj | 0.006 (-0.023 to 0.036) | 0.66 | 0.000 (-0.033 to 0.033) | 0.99 | -0.049 (-0.079 to -0.019) | 0.001 | 7647 |
| Fmin | 0.023 (-0.006 to 0.052) | 0.12 | -0.038 (-0.071 to -0.005) | 0.02 | -0.037 (-0.068 to -0.007) | 0.02 | 7670 |
| CC | 0.007 (-0.022 to 0.036) | 0.64 | -0.029 (-0.062 to 0.004) | 0.09 | -0.028 (-0.058 to 0.002) | 0.07 | 7668 |
| SLF (right) | -0.001 (-0.029 to 0.027) | 0.95 | -0.029 (-0.061 to 0.002) | 0.07 | -0.002 (-0.032 to 0.027) | 0.89 | 7682 |
| SLF (left) | 0.020 (-0.009 to 0.048) | 0.18 | -0.036 (-0.068 to -0.003) | 0.03 | 0.001 (-0.029 to 0.031) | 0.95 | 7672 |
| tSLF (left) | 0.017 (-0.012 to 0.045) | 0.25 | -0.035 (-0.068 to -0.003) | 0.03 | 0.003 (-0.026 to 0.033) | 0.83 | 7668 |
| pSLF (right) | 0.002 (-0.026 to 0.030) | 0.91 | -0.031 (-0.062 to 0.001) | 0.06 | -0.002 (-0.032 to 0.027) | 0.88 | 7683 |
| pSLF (left) | 0.021 (-0.007 to 0.050) | 0.15 | -0.037 (-0.070 to -0.005) | 0.03 | 0.001 (-0.029 to 0.031) | 0.94 | 7674 |
| SCS (left) | 0.008 (-0.019 to 0.036) | 0.56 | -0.034 (-0.066 to -0.002) | 0.04 | 0.007 (-0.022 to 0.036) | 0.64 | 7673 |
| SIFC (right) | 0.001 (-0.028 to 0.030) | 0.94 | -0.046 (-0.078 to -0.013) | 0.01 | -0.003 (-0.033 to 0.027) | 0.84 | 7676 |
| SIFC (left) | 0.008 (-0.020 to 0.037) | 0.57 | -0.035 (-0.068 to -0.002) | 0.04 | -0.007 (-0.038 to 0.023) | 0.63 | 7677 |
| IFSFC (right) | 0.007 (-0.021 to 0.035) | 0.62 | -0.040 (-0.073 to -0.008) | 0.02 | 0.010 (-0.020 to 0.040) | 0.51 | 7680 |
| IFSFC (left) | 0.001 (-0.028 to 0.029) | 0.97 | -0.038 (-0.071 to -0.005) | 0.02 | 0.013 (-0.017 to 0.043) | 0.40 | 7681 |
| **C) DTI-FA (DV)** | | | | | | | |
| Fx (left) | -0.020 (-0.049 to 0.008) | 0.16 | 0.008 (-0.025 to 0.040) | 0.64 | 0.049 (0.018 to 0.079) | 0.002 | 7687 |
| CST (right) | 0.010 (-0.018 to 0.038) | 0.49 | -0.007 (-0.039 to 0.025) | 0.67 | 0.043 (0.013 to 0.073) | 0.004 | 7683 |
| CST (left) | 0.032 (0.003 to 0.061) | 0.03 | 0.001 (-0.031 to 0.033) | 0.96 | 0.041 (0.011 to 0.071) | 0.01 | 7684 |
| SLF (right) | -0.040 (-0.069 to -0.010) | 0.01 | -0.007 (-0.040 to 0.026) | 0.69 | 0.057 (0.027 to 0.087) | < 0.001 | 7685 |
| SLF (left) | -0.063 (-0.092 to -0.034) | < 0.001 | 0.012 (-0.021 to 0.044) | 0.49 | 0.053 (0.023 to 0.084) | 0.001 | 7684 |
| tSLF (right) | -0.034 (-0.064 to -0.005) | 0.02 | -0.013 (-0.046 to 0.020) | 0.45 | 0.054 (0.024 to 0.084) | < 0.001 | 7686 |
| tSLF (left) | -0.060 (-0.089 to -0.031) | < 0.001 | 0.016 (-0.016 to 0.049) | 0.33 | 0.047 (0.016 to 0.077) | 0.002 | 7685 |
| pSLF (right) | -0.040 (-0.070 to -0.011) | 0.01 | -0.033 (-0.036 to 0.030) | 0.88 | 0.054 (0.024 to 0.085) | < 0.001 | 7685 |
| pSLF (left) | -0.056 (-0.085 to -0.027) | < 0.001 | 0.002 (-0.031 to 0.034) | 0.93 | 0.061 (0.031 to 0.091) | < 0.001 | 7682 |
| SCS (left) | 0.009 (-0.020 to 0.038) | 0.55 | -0.004 (-0.036 to 0.028) | 0.79 | 0.046 (0.016 to 0.076) | 0.002 | 7686 |

**4) Using only participants with full-term birth**

| **White matter tracts** | **SES indicators (IVs)** | | | | | | **Sample size (*n*)** |
| --- | --- | --- | --- | --- | --- | --- | --- |
|  | **Neighborhood disadvantage**  (higher = lower SES) | | **Household income**  (higher = higher SES) | | **Parental education**  (higher = higher SES) | |  |
|  | ***β* (95% CI)** | ***p*-value**  (nominal) | ***β* (95% CI)** | ***p*-value**  (nominal) | ***β* (95% CI)** | ***p*-value**  (nominal) |  |
| **A) RSI-RND (DV)** | | | | | | | |
| CST (right) | 0.012 (-0.017 to 0.041) | 0.43 | 0.006 (-0.027 to 0.039) | 0.71 | 0.051 (0.021 to 0.082) | 0.001 | 7147 |
| CST (left) | 0.025 (-0.005 to 0.055) | 0.10 | 0.008 (-0.026 to 0.041) | 0.64 | 0.055 (0.024 to 0.086) | 0.001 | 7145 |
| Fmaj | -0.050 (-0.080 to -0.019) | 0.001 | 0.015 (-0.019 to 0.049) | 0.39 | -0.014 (-0.045 to 0.018) | 0.40 | 7144 |
| SLF (right) | -0.025 (-0.055 to 0.004) | 0.10 | -0.004 (-0.038 to 0.029) | 0.80 | 0.058 (0.027 to 0.088) | < 0.001 | 7150 |
| SLF (left) | -0.064 (-0.094 to -0.035) | < 0.001 | 0.010 (-0.023 to 0.043) | 0.56 | 0.054 (0.023 to 0.085) | 0.001 | 7143 |
| tSLF (right) | -0.025 (-0.055 to 0.005) | 0.11 | -0.002 (-0.036 to 0.032) | 0.92 | 0.051 (0.019 to 0.082) | 0.002 | 7149 |
| tSLF (left) | -0.066 (-0.095 to -0.036) | < 0.001 | 0.014 (-0.019 to 0.048) | 0.39 | 0.045 (0.014 to 0.075) | 0.004 | 7144 |
| pSLF (right) | -0.026 (-0.055 to 0.004) | 0.09 | -0.006 (-0.039 to 0.028) | 0.74 | 0.059 (0.028 to 0.090) | < 0.001 | 7148 |
| pSLF (left) | -0.045 (-0.074 to -0.016) | 0.002 | 0.000 (-0.032 to 0.033) | 0.98 | 0.068 (0.037 to 0.098) | < 0.001 | 7142 |
| **B) RSI-RNI (DV)** | | | | | | | |
| Fx (right) | 0.057 (0.027 to 0.087) | < 0.001 | 0.004 (-0.030 to 0.039) | 0.81 | -0.010 (-0.042 to 0.022) | 0.54 | 7133 |
| Fx (left) | 0.059 (0.029 to 0.089) | < 0.001 | -0.019 (-0.053 to 0.016) | 0.28 | -0.001 (-0.033 to 0.031) | 0.94 | 7118 |
| CgC (right) | 0.029 (0.000 to 0.059) | 0.05 | -0.031 (-0.065 to 0.003) | 0.07 | -0.015 (-0.046 to 0.017) | 0.36 | 7141 |
| CgC (left) | 0.027 (-0.004 to 0.057) | 0.08 | -0.039 (-0.073 to -0.004) | 0.03 | -0.019 (-0.051 to 0.013) | 0.24 | 7133 |
| CgH (right) | 0.066 (0.036 to 0.096) | < 0.001 | -0.020 (-0.053 to 0.014) | 0.26 | 0.006 (-0.026 to 0.037) | 0.71 | 7139 |
| CgH (left) | 0.076 (0.046 to 0.106) | < 0.001 | -0.036 (-0.070 to -0.002) | 0.04 | -0.007 (-0.039 to 0.024) | 0.65 | 7127 |
| CST (right) | 0.053 (0.023 to 0.083) | 0.001 | -0.025 (-0.059 to 0.009) | 0.15 | -0.001 (-0.032 to 0.031) | 0.97 | 7145 |
| CST (left) | 0.055 (0.025 to 0.085) | < 0.001 | -0.041 (-0.075 to -0.007) | 0.02 | -0.001 (-0.033 to 0.030) | 0.94 | 7138 |
| ATR (right) | 0.053 (0.023 to 0.083) | 0.001 | -0.041 (-0.075 to -0.007) | 0.02 | 0.000 (-0.032 to 0.031) | 0.98 | 7115 |
| ATR (left) | 0.054 (0.023 to 0.084) | 0.001 | -0.042 (-0.077 to -0.008) | 0.02 | -0.012 (-0.043 to 0.020) | 0.47 | 7117 |
| Unc (right) | 0.028 (-0.002 to 0.058) | 0.07 | -0.052 (-0.086 to -0.018) | 0.003 | -0.001 (-0.033 to 0.031) | 0.95 | 7141 |
| Unc (left) | 0.019 (-0.011 to 0.050) | 0.20 | -0.047 (-0.081 to -0.013) | 0.01 | -0.006 (-0.038 to 0.025) | 0.69 | 7143 |
| ILF (right) | 0.005 (-0.025 to 0.034) | 0.75 | -0.035 (-0.069 to -0.002) | 0.04 | 0.000 (-0.031 to 0.031) | > 0.99 | 7152 |
| ILF (left) | 0.005 (-0.025 to 0.035) | 0.73 | -0.035 (-0.070 to -0.001) | 0.04 | -0.012 (-0.044 to 0.019) | 0.44 | 7145 |
| IFOF (right) | 0.008 (-0.022 to 0.037) | 0.61 | -0.052 (-0.086 to -0.019) | 0.002 | -0.005 (-0.036 to 0.026) | 0.75 | 7148 |
| IFOF (left) | 0.008 (-0.022 to 0.037) | 0.61 | -0.056 (-0.089 to -0.022) | 0.001 | -0.010 (-0.041 to 0.022) | 0.55 | 7131 |
| Fmaj | 0.014 (-0.017 to 0.044) | 0.37 | -0.007 (-0.041 to 0.027) | 0.69 | -0.044 (-0.076 to -0.012) | 0.01 | 7124 |
| Fmin | 0.021 (-0.010 to 0.051) | 0.19 | -0.042 (-0.077 to -0.008) | 0.02 | -0.047 (-0.079 to -0.015) | 0.004 | 7138 |
| CC | 0.014 (-0.016 to 0.044) | 0.36 | -0.038 (-0.072 to -0.003) | 0.03 | -0.022 (-0.054 to 0.010) | 0.18 | 7138 |
| SLF (right) | 0.000 (-0.030 to 0.029) | 0.98 | -0.031 (-0.064 to 0.002) | 0.06 | -0.007 (-0.038 to 0.024) | 0.67 | 7152 |
| SLF (left) | 0.021 (-0.008 to 0.051) | 0.16 | -0.051 (-0.084 to -0.017) | 0.003 | 0.002 (-0.029 to 0.033) | 0.89 | 7144 |
| tSLF (left) | 0.020 (-0.010 to 0.050) | 0.19 | -0.051 (-0.084 to -0.018) | 0.003 | 0.007 (-0.025 to 0.038) | 0.68 | 7142 |
| pSLF (right) | 0.002 (-0.028 to 0.031) | 0.91 | -0.032 (-0.065 to 0.001) | 0.06 | -0.007 (-0.038 to 0.024) | 0.65 | 7153 |
| pSLF (left) | 0.023 (-0.007 to 0.053) | 0.13 | -0.048 (-0.081 to -0.015) | 0.005 | 0.002 (-0.029 to 0.034) | 0.89 | 7145 |
| SCS (left) | 0.009 (-0.020 to 0.038) | 0.55 | -0.046 (-0.079 to -0.013) | 0.01 | 0.010 (-0.021 to 0.041) | 0.51 | 7144 |
| SIFC (right) | -0.003 (-0.033 to 0.028) | 0.87 | -0.041 (-0.075 to -0.007) | 0.02 | -0.012 (-0.044 to 0.020) | 0.45 | 7145 |
| SIFC (left) | 0.003 (-0.026 to 0.033) | 0.82 | -0.034 (-0.069 to 0.000) | 0.05 | -0.014 (-0.045 to 0.018) | 0.40 | 7146 |
| IFSFC (right) | 0.010 (-0.019 to 0.040) | 0.49 | -0.047 (-0.081 to -0.014) | 0.01 | 0.012 (-0.020 to 0.043) | 0.47 | 7150 |
| IFSFC (left) | 0.003 (-0.027 to 0.033) | 0.84 | -0.046 (-0.079 to -0.012) | 0.01 | 0.012 (-0.020 to 0.043) | 0.47 | 7149 |
| **C) DTI-FA (DV)** | | | | | | | |
| Fx (left) | -0.029 (-0.059 to 0.001) | 0.06 | 0.007 (-0.027 to 0.041) | 0.69 | 0.035 (0.003 to 0.067) | 0.03 | 7156 |
| CST (right) | 0.015 (-0.015 to 0.044) | 0.33 | -0.006 (-0.039 to 0.028) | 0.74 | 0.055 (0.024 to 0.086) | 0.001 | 7155 |
| CST (left) | 0.032 (0.002 to 0.062) | 0.04 | -0.005 (-0.039 to 0.028) | 0.75 | 0.056 (0.024 to 0.087) | < 0.001 | 7154 |
| SLF (right) | -0.040 (-0.071 to -0.009) | 0.01 | -0.002 (-0.036 to 0.032) | 0.90 | 0.060 (0.028 to 0.092) | < 0.001 | 7155 |
| SLF (left) | -0.070 (-0.101 to -0.040) | < 0.001 | 0.017 (-0.017 to 0.051) | 0.33 | 0.052 (0.020 to 0.083) | 0.001 | 7154 |
| tSLF (right) | -0.035 (-0.066 to -0.005) | 0.02 | -0.004 (-0.038 to 0.031) | 0.83 | 0.052 (0.020 to 0.084) | 0.001 | 7155 |
| tSLF (left) | -0.069 (-0.099 to -0.039) | < 0.001 | 0.020 (-0.014 to 0.054) | 0.24 | 0.043 (0.011 to 0.074) | 0.01 | 7155 |
| pSLF (right) | -0.040 (-0.070 to -0.009) | 0.01 | 0.000 (-0.035 to 0.034) | 0.98 | 0.060 (0.028 to 0.092) | < 0.001 | 7155 |
| pSLF (left) | -0.059 (-0.089 to -0.029) | < 0.001 | 0.011 (-0.023 to 0.044) | 0.53 | 0.065 (0.033 to 0.096) | < 0.001 | 7151 |
| SCS (left) | 0.005 (-0.024 to 0.035) | 0.72 | 0.004 (-0.030 to 0.037) | 0.82 | 0.042 (0.010 to 0.073) | 0.01 | 7155 |

*Note*. Sensitivity analyses were conducted on significant associations between socioeconomic status (SES) and white matter microstructure that were observed in the whole study sample. Effects that were significant in the main analyses (**eTable 8** in the **Supplement**) and are nominally significant (*p*-value ≤ 0.05) here in sensitivity analyses were considered robust and are demarcated in green. Effects that were significant in the main analyses, but not nominally significant here, are demarcated in gray. Unhighlighted effects were not significant in the main analyses. Most findings were robust to sensitivity analyses, and there remained spatial overlaps of effects shared between neighborhood and household SES indicators. Findings that failed to replicate mostly involve associations between household income and RSI-RNI in distributed white matter tracts. Research has reported on white matter alterations in children and adolescents who experienced childhood adversity^7,8^, had early-onset psychosis^9^, had major depression^10^, and who were born pre-term^11^. In particular, purported neuroinflammatory phenotypes, which RSI-RNI could potentially reflect, were seen in white matter regions in those who had adverse childhood experiences^7^ or depression^10^. It is possible that these associations were reflected in our main analyses and were excluded in these more restricted subsamples. Furthermore, the subsample of participants without reported adverse childhood experience had about 36% reduction in sample size relative to the whole study sample; it is thus possible that some effects became nonsignificant due to decreased statistical power. Model setup was identical to main analyses; refer to **eTable 8** in the **Supplement** for full specification. RSI, restriction spectrum imaging; RND, restricted normalized directional; RNI, restricted normalized isotropic; DTI, diffusion tensor imaging; FA, fractional anisotropy; MD, mean diffusivity. Full names of white matter tracts are shown in the main text.

**eFigure 5. Associations between SES and white matter DTI-FA**

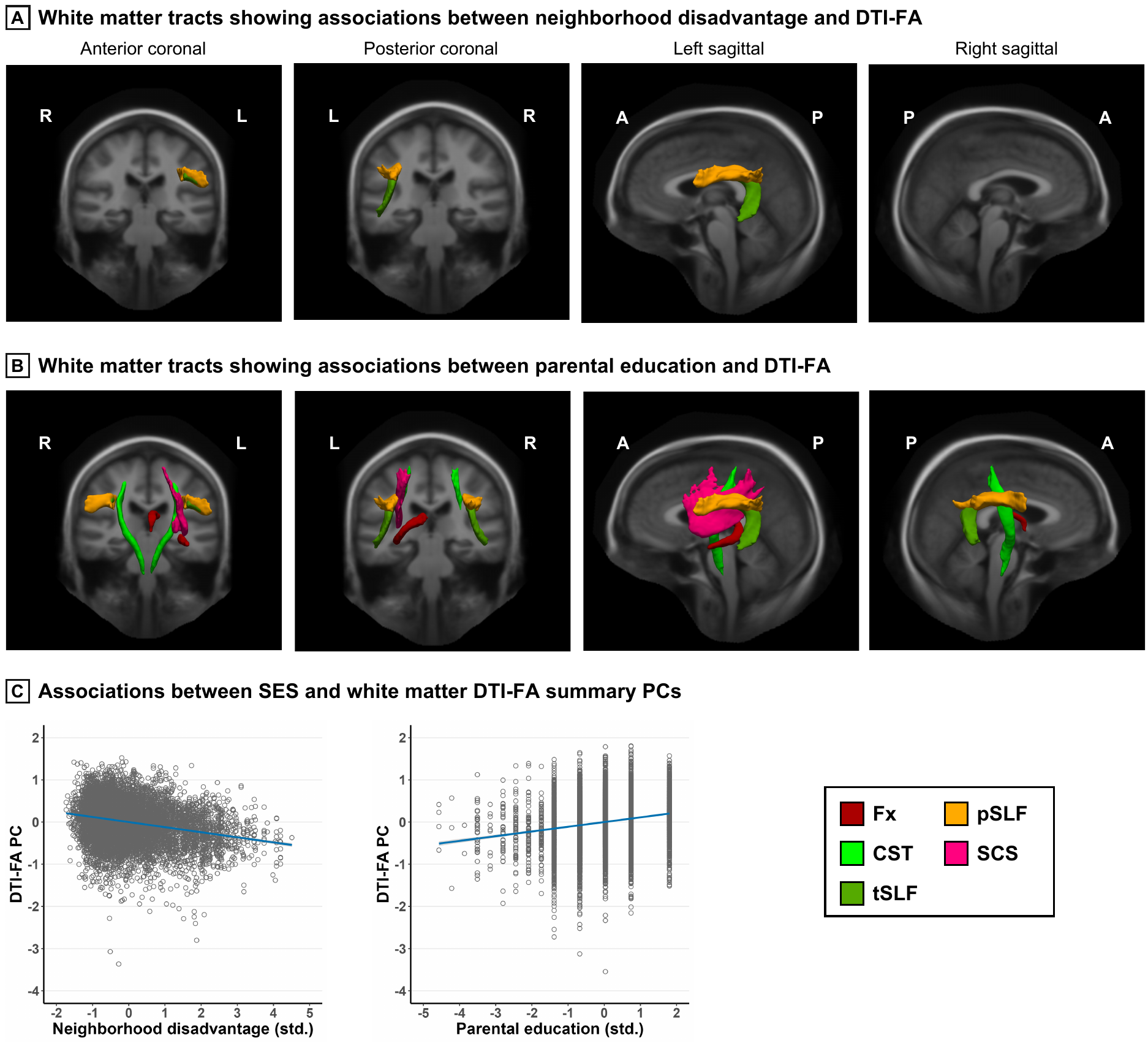

*Note*. Higher neighborhood disadvantage (**A**) and lower parental education (**B**) had independent associations with lower DTI-FA in distributed white matter tracts, overlapping in the superior longitudinal fasciculus (SLF; including the parietal (p) and temporal (t) subregions). Tracts that had significant effects are visualized by color code in coronal and sagittal slices using the AtlasTrack atlas. R, right; L, left; A, anterior; P, posterior; Fx, fornix; CST, corticospinal/pyramidal tract; SCS, superior-corticostriatal tract; PC, principal component. (**C**) Negative associations were seen between SES and white matter DTI-FA PCs. Regression lines were partialled for covariates and flanked by 95% confidence interval shaded in blue. Data points were residuals partialled for covariates extracted from a randomly selected imputed dataset. DTI, diffusion tensor imaging; FA, fractional anisotropy; SES, socioeconomic status

**eTable 10. Associations between SES and white matter microstructure, adjusting for race/ethnicity**

| **White matter tracts** | **SES indicators (IVs)** | | | | | | | | |
| --- | --- | --- | --- | --- | --- | --- | --- | --- | --- |
|  | **Neighborhood disadvantage**  (higher = lower SES) | | | **Household income**  (higher = higher SES) | | | **Parental education**  (higher = higher SES) | | |
|  | ***β* (95% CI)** | ***p*-value**  (nominal) | ***p*-value**  (FDR) | ***β* (95% CI)** | ***p*-value**  (nominal) | ***p*-value**  (FDR) | ***β* (95% CI)** | ***p*-value**  (nominal) | ***p*-value**  (FDR) |
| **A) RSI-RND (DV)** | | | | | | | | | |
| Fx (right) | 0.011 (-0.018 to 0.039) | 0.47 | 0.85 | 0.020 (-0.012 to 0.051) | 0.22 | 0.91 | 0.028 (-0.001 to 0.057) | 0.06 | 0.12 |
| Fx (left) | 0.008 (-0.021 to 0.037) | 0.59 | 0.85 | 0.005 (-0.027 to 0.037) | 0.75 | 0.91 | 0.027 (-0.002 to 0.056) | 0.07 | 0.13 |
| CgC (right) | 0.002 (-0.026 to 0.030) | 0.88 | 0.93 | -0.016 (-0.046 to 0.015) | 0.31 | 0.91 | 0.015 (-0.013 to 0.043) | 0.31 | 0.41 |
| CgC (left) | -0.005 (-0.034 to 0.023) | 0.71 | 0.85 | -0.010 (-0.040 to 0.021) | 0.53 | 0.91 | -0.008 (-0.036 to 0.020) | 0.57 | 0.63 |
| CgH (right) | -0.014 (-0.042 to 0.015) | 0.34 | 0.85 | -0.008 (-0.039 to 0.022) | 0.59 | 0.91 | 0.033 (0.004 to 0.061) | 0.02 | 0.08 |
| CgH (left) | 0.008 (-0.020 to 0.036) | 0.58 | 0.85 | 0.010 (-0.021 to 0.040) | 0.54 | 0.91 | 0.030 (0.002 to 0.058) | 0.04 | 0.10 |
| CST (right) | 0.006 (-0.021 to 0.034) | 0.66 | 0.85 | 0.017 (-0.013 to 0.047) | 0.26 | 0.91 | 0.040 (0.012 to 0.068) | 0.005 | **0.02** |
| CST (left) | 0.016 (-0.012 to 0.044) | 0.26 | 0.85 | 0.013 (-0.017 to 0.044) | 0.39 | 0.91 | 0.049 (0.020 to 0.077) | 0.001 | **0.004** |
| ATR (right) | 0.021 (-0.008 to 0.049) | 0.15 | 0.85 | 0.024 (-0.007 to 0.056) | 0.13 | 0.91 | 0.015 (-0.014 to 0.044) | 0.31 | 0.41 |
| ATR (left) | 0.008 (-0.021 to 0.037) | 0.60 | 0.85 | 0.027 (-0.005 to 0.058) | 0.10 | 0.91 | 0.016 (-0.013 to 0.045) | 0.27 | 0.40 |
| Unc (right) | -0.005 (-0.034 to 0.023) | 0.71 | 0.85 | -0.029 (-0.059 to 0.001) | 0.06 | 0.91 | 0.012 (-0.015 to 0.040) | 0.38 | 0.48 |
| Unc (left) | 0.014 (-0.015 to 0.043) | 0.33 | 0.85 | -0.023 (-0.053 to 0.007) | 0.13 | 0.91 | 0.014 (-0.015 to 0.042) | 0.34 | 0.44 |
| ILF (right) | 0.003 (-0.025 to 0.031) | 0.82 | 0.90 | -0.017 (-0.048 to 0.013) | 0.26 | 0.91 | 0.026 (-0.002 to 0.054) | 0.07 | 0.13 |
| ILF (left) | 0.025 (-0.003 to 0.053) | 0.09 | 0.66 | -0.005 (-0.036 to 0.025) | 0.74 | 0.91 | 0.021 (-0.007 to 0.049) | 0.14 | 0.22 |
| IFOF (right) | -0.020 (-0.048 to 0.008) | 0.17 | 0.85 | -0.013 (-0.044 to 0.017) | 0.38 | 0.91 | 0.031 (0.003 to 0.059) | 0.03 | 0.10 |
| IFOF (left) | -0.013 (-0.041 to 0.015) | 0.38 | 0.85 | -0.009 (-0.039 to 0.020) | 0.54 | 0.91 | 0.027 (0.000 to 0.055) | 0.05 | 0.12 |
| Fmaj | -0.011 (-0.040 to 0.017) | 0.44 | 0.85 | 0.005 (-0.026 to 0.035) | 0.77 | 0.91 | -0.010 (-0.038 to 0.019) | 0.51 | 0.58 |
| Fmin | 0.011 (-0.018 to 0.039) | 0.47 | 0.85 | -0.013 (-0.043 to 0.018) | 0.42 | 0.91 | -0.010 (-0.038 to 0.018) | 0.49 | 0.58 |
| CC | -0.006 (-0.034 to 0.022) | 0.68 | 0.85 | -0.005 (-0.035 to 0.025) | 0.74 | 0.91 | -0.003 (-0.031 to 0.025) | 0.83 | 0.86 |
| SLF (right) | -0.013 (-0.041 to 0.016) | 0.38 | 0.85 | -0.003 (-0.033 to 0.028) | 0.86 | 0.94 | 0.051 (0.024 to 0.079) | < 0.001 | **0.003** |
| SLF (left) | -0.032 (-0.060 to -0.004) | 0.02 | 0.53 | 0.001 (-0.029 to 0.031) | 0.94 | 0.94 | 0.049 (0.021 to 0.076) | 0.001 | **0.004** |
| tSLF (right) | -0.011 (-0.040 to 0.017) | 0.45 | 0.85 | -0.005 (-0.035 to 0.026) | 0.78 | 0.91 | 0.043 (0.014 to 0.071) | 0.003 | **0.02** |
| tSLF (left) | -0.030 (-0.058 to -0.002) | 0.03 | 0.53 | 0.004 (-0.026 to 0.034) | 0.79 | 0.91 | 0.040 (0.013 to 0.068) | 0.004 | **0.02** |
| pSLF (right) | -0.014 (-0.043 to 0.014) | 0.31 | 0.85 | -0.002 (-0.032 to 0.028) | 0.89 | 0.94 | 0.052 (0.024 to 0.080) | < 0.001 | **0.003** |
| pSLF (left) | -0.028 (-0.055 to 0.000) | 0.05 | 0.53 | -0.004 (-0.034 to 0.026) | 0.79 | 0.91 | 0.058 (0.031 to 0.086) | < 0.001 | **0.001** |
| SCS (right) | 0.010 (-0.019 to 0.038) | 0.50 | 0.85 | -0.011 (-0.042 to 0.020) | 0.49 | 0.91 | 0.030 (0.001 to 0.058) | 0.04 | 0.10 |
| SCS (left) | 0.010 (-0.018 to 0.038) | 0.49 | 0.85 | -0.001 (-0.031 to 0.029) | 0.94 | 0.94 | 0.029 (0.001 to 0.056) | 0.04 | 0.11 |
| SIFC (right) | 0.000 (-0.030 to 0.029) | 0.98 | 0.98 | -0.016 (-0.046 to 0.015) | 0.32 | 0.91 | -0.004 (-0.032 to 0.025) | 0.81 | 0.86 |
| SIFC (left) | -0.007 (-0.036 to 0.021) | 0.61 | 0.85 | -0.015 (-0.045 to 0.015) | 0.33 | 0.91 | 0.000 (-0.028 to 0.028) | > 0.99 | > 0.99 |
| IFSFC (right) | -0.004 (-0.032 to 0.024) | 0.76 | 0.88 | 0.013 (-0.017 to 0.043) | 0.39 | 0.91 | 0.023 (-0.004 to 0.051) | 0.10 | 0.17 |
| IFSFC (left) | -0.002 (-0.029 to 0.026) | 0.90 | 0.93 | 0.006 (-0.022 to 0.036) | 0.67 | 0.91 | 0.023 (-0.005 to 0.050) | 0.10 | 0.17 |

**(continued) eTable 10. Associations between SES and white matter microstructure, adjusting for race/ethnicity**

| **White matter tracts** | **SES indicators (IVs)** | | | | | | | | |
| --- | --- | --- | --- | --- | --- | --- | --- | --- | --- |
|  | **Neighborhood disadvantage**  (higher = lower SES) | | | **Household income**  (higher = higher SES) | | | **Parental education**  (higher = higher SES) | | |
|  | ***β* (95% CI)** | ***p*-value**  (nominal) | ***p*-value**  (FDR) | ***β* (95% CI)** | ***p*-value**  (nominal) | ***p*-value**  (FDR) | ***β* (95% CI)** | ***p*-value**  (nominal) | ***p*-value**  (FDR) |
| **B) RSI-RNI (DV)** | | | | | | | | | |
| Fx (right) | 0.029 (0.001 to 0.057) | 0.05 | 0.70 | 0.018 (-0.013 to 0.050) | 0.26 | 0.31 | -0.004 (-0.033 to 0.025) | 0.78 | 0.96 |
| Fx (left) | 0.024 (-0.004 to 0.053) | 0.09 | 0.70 | 0.009 (-0.023 to 0.041) | 0.58 | 0.62 | -0.010 (-0.039 to 0.019) | 0.48 | 0.96 |
| CgC (right) | 0.008 (-0.020 to 0.036) | 0.56 | 0.84 | -0.026 (-0.056 to 0.005) | 0.10 | 0.14 | -0.002 (-0.030 to 0.026) | 0.90 | 0.96 |
| CgC (left) | 0.007 (-0.021 to 0.036) | 0.62 | 0.84 | -0.032 (-0.063 to -0.001) | 0.05 | 0.09 | -0.007 (-0.036 to 0.022) | 0.63 | 0.96 |
| CgH (right) | 0.018 (-0.010 to 0.046) | 0.20 | 0.70 | -0.007 (-0.037 to 0.023) | 0.65 | 0.66 | 0.013 (-0.016 to 0.041) | 0.38 | 0.96 |
| CgH (left) | 0.021 (-0.008 to 0.049) | 0.15 | 0.70 | -0.025 (-0.055 to 0.005) | 0.11 | 0.14 | 0.005 (-0.023 to 0.033) | 0.71 | 0.96 |
| CST (right) | -0.003 (-0.031 to 0.026) | 0.86 | 0.99 | -0.007 (-0.038 to 0.024) | 0.66 | 0.66 | 0.005 (-0.023 to 0.034) | 0.73 | 0.96 |
| CST (left) | -0.007 (-0.035 to 0.021) | 0.62 | 0.84 | -0.015 (-0.046 to 0.016) | 0.34 | 0.39 | 0.001 (-0.027 to 0.029) | 0.94 | 0.96 |
| ATR (right) | 0.018 (-0.010 to 0.046) | 0.21 | 0.70 | -0.031 (-0.062 to -0.001) | 0.05 | 0.09 | 0.012 (-0.017 to 0.040) | 0.42 | 0.96 |
| ATR (left) | 0.011 (-0.018 to 0.039) | 0.45 | 0.83 | -0.026 (-0.057 to 0.006) | 0.11 | 0.14 | -0.001 (-0.029 to 0.028) | 0.96 | 0.96 |
| Unc (right) | -0.006 (-0.035 to 0.022) | 0.66 | 0.85 | -0.046 (-0.077 to -0.015) | 0.003 | **0.05** | 0.015 (-0.014 to 0.043) | 0.31 | 0.96 |
| Unc (left) | -0.004 (-0.032 to 0.025) | 0.79 | 0.94 | -0.042 (-0.073 to -0.011) | 0.01 | **0.05** | 0.006 (-0.023 to 0.034) | 0.70 | 0.96 |
| ILF (right) | -0.019 (-0.047 to 0.009) | 0.19 | 0.70 | -0.034 (-0.065 to -0.003) | 0.03 | 0.08 | 0.003 (-0.025 to 0.031) | 0.82 | 0.96 |
| ILF (left) | -0.007 (-0.036 to 0.021) | 0.61 | 0.84 | -0.029 (-0.060 to 0.002) | 0.06 | 0.12 | -0.008 (-0.036 to 0.021) | 0.59 | 0.96 |
| IFOF (right) | -0.017 (-0.045 to 0.011) | 0.23 | 0.70 | -0.047 (-0.077 to -0.016) | 0.003 | **0.05** | 0.002 (-0.026 to 0.030) | 0.89 | 0.96 |
| IFOF (left) | -0.015 (-0.042 to 0.013) | 0.30 | 0.70 | -0.043 (-0.074 to -0.013) | 0.01 | **0.05** | -0.002 (-0.031 to 0.026) | 0.87 | 0.96 |
| Fmaj | 0.008 (-0.021 to 0.037) | 0.57 | 0.84 | -0.011 (-0.042 to 0.020) | 0.50 | 0.55 | -0.040 (-0.068 to -0.011) | 0.01 | 0.20 |
| Fmin | 0.001 (-0.028 to 0.030) | 0.94 | 0.99 | -0.040 (-0.071 to -0.009) | 0.01 | 0.06 | -0.029 (-0.058 to 0.000) | 0.05 | 0.80 |
| CC | -0.005 (-0.033 to 0.024) | 0.75 | 0.93 | -0.034 (-0.065 to -0.002) | 0.04 | 0.09 | -0.017 (-0.046 to 0.012) | 0.25 | 0.96 |
| SLF (right) | -0.013 (-0.041 to 0.015) | 0.37 | 0.71 | -0.027 (-0.057 to 0.003) | 0.08 | 0.12 | -0.004 (-0.032 to 0.024) | 0.76 | 0.96 |
| SLF (left) | 0.000 (-0.028 to 0.029) | 0.99 | 0.99 | -0.037 (-0.068 to -0.007) | 0.02 | 0.06 | 0.002 (-0.026 to 0.030) | 0.88 | 0.96 |
| tSLF (right) | -0.010 (-0.038 to 0.018) | 0.50 | 0.84 | -0.025 (-0.055 to 0.006) | 0.11 | 0.14 | -0.002 (-0.030 to 0.026) | 0.89 | 0.96 |
| tSLF (left) | 0.001 (-0.028 to 0.029) | 0.95 | 0.99 | -0.037 (-0.068 to -0.007) | 0.02 | 0.06 | 0.004 (-0.024 to 0.032) | 0.79 | 0.96 |
| pSLF (right) | -0.014 (-0.042 to 0.014) | 0.34 | 0.70 | -0.027 (-0.057 to 0.003) | 0.08 | 0.12 | -0.003 (-0.031 to 0.025) | 0.83 | 0.96 |
| pSLF (left) | -0.001 (-0.029 to 0.028) | 0.97 | 0.99 | -0.037 (-0.068 to -0.007) | 0.02 | 0.06 | 0.006 (-0.022 to 0.034) | 0.69 | 0.96 |
| SCS (right) | -0.015 (-0.043 to 0.013) | 0.29 | 0.70 | -0.022 (-0.053 to 0.008) | 0.15 | 0.19 | 0.014 (-0.014 to 0.042) | 0.34 | 0.96 |
| SCS (left) | -0.015 (-0.042 to 0.013) | 0.29 | 0.70 | -0.029 (-0.060 to 0.001) | 0.06 | 0.11 | 0.010 (-0.018 to 0.038) | 0.49 | 0.96 |
| SIFC (right) | -0.014 (-0.043 to 0.015) | 0.34 | 0.70 | -0.043 (-0.074 to -0.012) | 0.01 | **0.05** | 0.001 (-0.028 to 0.030) | 0.94 | 0.96 |
| SIFC (left) | -0.019 (-0.047 to 0.009) | 0.19 | 0.70 | -0.029 (-0.060 to 0.002) | 0.07 | 0.12 | 0.001 (-0.027 to 0.030) | 0.94 | 0.96 |
| IFSFC (right) | -0.017 (-0.045 to 0.011) | 0.23 | 0.70 | -0.035 (-0.066 to -0.004) | 0.03 | 0.08 | 0.014 (-0.015 to 0.042) | 0.34 | 0.96 |
| IFSFC (left) | -0.023 (-0.051 to 0.006) | 0.11 | 0.70 | -0.035 (-0.066 to -0.004) | 0.03 | 0.08 | 0.015 (-0.013 to 0.043) | 0.30 | 0.96 |

**(continued) eTable 10. Associations between SES and white matter microstructure, adjusting for race/ethnicity**

| **White matter tracts** | **SES indicators (IVs)** | | | | | | | | |
| --- | --- | --- | --- | --- | --- | --- | --- | --- | --- |
|  | **Neighborhood disadvantage**  (higher = lower SES) | | | **Household income**  (higher = higher SES) | | | **Parental education**  (higher = higher SES) | | |
|  | ***β* (95% CI)** | ***p*-value**  (nominal) | ***p*-value**  (FDR) | ***β* (95% CI)** | ***p*-value**  (nominal) | ***p*-value**  (FDR) | ***β* (95% CI)** | ***p*-value**  (nominal) | ***p*-value**  (FDR) |
| **C) DTI-FA (DV)** | | | | | | | | | |
| Fx (right) | 0.004 (-0.025 to 0.033) | 0.79 | 0.94 | 0.022 (-0.009 to 0.053) | 0.17 | 0.98 | 0.029 (0.000 to 0.058) | 0.05 | 0.10 |
| Fx (left) | 0.001 (-0.027 to 0.030) | 0.93 | 0.96 | -0.005 (-0.036 to 0.026) | 0.75 | 0.98 | 0.038 (0.009 to 0.066) | 0.01 | **0.03** |
| CgC (right) | 0.000 (-0.029 to 0.028) | 0.98 | 0.98 | -0.024 (-0.055 to 0.007) | 0.13 | 0.98 | 0.010 (-0.018 to 0.039) | 0.49 | 0.61 |
| CgC (left) | -0.006 (-0.035 to 0.023) | 0.68 | 0.94 | -0.015 (-0.045 to 0.016) | 0.35 | 0.98 | -0.008 (-0.036 to 0.021) | 0.60 | 0.69 |
| CgH (right) | -0.010 (-0.039 to 0.018) | 0.47 | 0.94 | -0.003 (-0.033 to 0.028) | 0.86 | 0.98 | 0.011 (-0.017 to 0.040) | 0.43 | 0.58 |
| CgH (left) | 0.004 (-0.025 to 0.032) | 0.80 | 0.94 | 0.014 (-0.017 to 0.044) | 0.38 | 0.98 | 0.031 (0.002 to 0.059) | 0.03 | 0.09 |
| CST (right) | 0.006 (-0.022 to 0.034) | 0.69 | 0.94 | 0.002 (-0.029 to 0.032) | 0.92 | 0.98 | 0.044 (0.016 to 0.072) | 0.002 | **0.01** |
| CST (left) | 0.026 (-0.002 to 0.055) | 0.07 | 0.46 | 0.002 (-0.029 to 0.033) | 0.90 | 0.98 | 0.046 (0.018 to 0.074) | 0.002 | **0.01** |
| ATR (right) | 0.017 (-0.012 to 0.046) | 0.25 | 0.78 | 0.018 (-0.013 to 0.050) | 0.25 | 0.98 | 0.016 (-0.013 to 0.044) | 0.29 | 0.41 |
| ATR (left) | 0.009 (-0.021 to 0.038) | 0.57 | 0.94 | 0.030 (-0.002 to 0.061) | 0.06 | 0.98 | 0.018 (-0.011 to 0.047) | 0.23 | 0.33 |
| Unc (right) | -0.008 (-0.037 to 0.022) | 0.60 | 0.94 | -0.020 (-0.051 to 0.011) | 0.20 | 0.98 | 0.005 (-0.024 to 0.033) | 0.74 | 0.79 |
| Unc (left) | 0.006 (-0.024 to 0.035) | 0.71 | 0.94 | -0.016 (-0.047 to 0.015) | 0.32 | 0.98 | 0.008 (-0.021 to 0.036) | 0.60 | 0.69 |
| ILF (right) | 0.004 (-0.025 to 0.032) | 0.80 | 0.94 | -0.008 (-0.039 to 0.023) | 0.61 | 0.98 | 0.025 (-0.003 to 0.053) | 0.08 | 0.15 |
| ILF (left) | 0.020 (-0.009 to 0.049) | 0.17 | 0.67 | 0.004 (-0.027 to 0.035) | 0.79 | 0.98 | 0.021 (-0.008 to 0.049) | 0.15 | 0.25 |
| IFOF (right) | -0.017 (-0.046 to 0.011) | 0.23 | 0.78 | -0.011 (-0.042 to 0.019) | 0.47 | 0.98 | 0.035 (0.007 to 0.064) | 0.01 | **0.04** |
| IFOF (left) | -0.005 (-0.033 to 0.024) | 0.74 | 0.94 | 0.004 (-0.027 to 0.034) | 0.81 | 0.98 | 0.030 (0.002 to 0.058) | 0.04 | 0.09 |
| Fmaj | -0.003 (-0.032 to 0.026) | 0.82 | 0.94 | 0.000 (-0.031 to 0.031) | 0.99 | 0.99 | 0.010 (-0.018 to 0.039) | 0.47 | 0.61 |
| Fmin | 0.007 (-0.023 to 0.037) | 0.63 | 0.94 | -0.011 (-0.043 to 0.020) | 0.48 | 0.98 | -0.021 (-0.050 to 0.009) | 0.17 | 0.26 |
| CC | -0.003 (-0.031 to 0.026) | 0.85 | 0.95 | -0.009 (-0.040 to 0.022) | 0.57 | 0.98 | 0.002 (-0.027 to 0.030) | 0.91 | 0.94 |
| SLF (right) | -0.025 (-0.055 to 0.004) | 0.09 | 0.46 | -0.008 (-0.040 to 0.024) | 0.62 | 0.98 | 0.049 (0.020 to 0.078) | 0.001 | **0.01** |
| SLF (left) | -0.036 (-0.065 to -0.007) | 0.01 | 0.24 | -0.002 (-0.033 to 0.029) | 0.89 | 0.98 | 0.048 (0.019 to 0.076) | 0.001 | **0.01** |
| tSLF (right) | -0.023 (-0.052 to 0.006) | 0.12 | 0.54 | -0.014 (-0.045 to 0.018) | 0.39 | 0.98 | 0.042 (0.013 to 0.071) | 0.005 | **0.02** |
| tSLF (left) | -0.033 (-0.062 to -0.005) | 0.02 | 0.24 | 0.001 (-0.030 to 0.032) | 0.95 | 0.98 | 0.040 (0.011 to 0.068) | 0.01 | **0.03** |
| pSLF (right) | -0.025 (-0.054 to 0.004) | 0.09 | 0.46 | -0.005 (-0.036 to 0.027) | 0.78 | 0.98 | 0.048 (0.020 to 0.077) | 0.001 | **0.01** |
| pSLF (left) | -0.034 (-0.063 to -0.005) | 0.02 | 0.24 | -0.006 (-0.036 to 0.025) | 0.72 | 0.98 | 0.058 (0.029 to 0.086) | < 0.001 | **0.002** |
| SCS (right) | 0.011 (-0.017 to 0.040) | 0.44 | 0.94 | -0.009 (-0.040 to 0.022) | 0.58 | 0.98 | 0.030 (0.001 to 0.059) | 0.04 | 0.09 |
| SCS (left) | 0.014 (-0.014 to 0.042) | 0.33 | 0.93 | -0.001 (-0.032 to 0.029) | 0.93 | 0.98 | 0.035 (0.007 to 0.064) | 0.01 | **0.04** |
| SIFC (right) | -0.002 (-0.031 to 0.028) | 0.92 | 0.96 | -0.022 (-0.054 to 0.009) | 0.16 | 0.98 | 0.006 (-0.023 to 0.035) | 0.70 | 0.78 |
| SIFC (left) | -0.003 (-0.033 to 0.026) | 0.82 | 0.94 | -0.010 (-0.041 to 0.021) | 0.53 | 0.98 | 0.000 (-0.028 to 0.029) | 0.98 | 0.98 |
| IFSFC (right) | -0.009 (-0.038 to 0.020) | 0.54 | 0.94 | 0.003 (-0.028 to 0.034) | 0.84 | 0.98 | 0.026 (-0.002 to 0.055) | 0.07 | 0.14 |
| IFSFC (left) | -0.005 (-0.033 to 0.024) | 0.75 | 0.94 | -0.002 (-0.033 to 0.029) | 0.88 | 0.98 | 0.023 (-0.005 to 0.052) | 0.11 | 0.19 |

**(continued) eTable 10. Associations between SES and white matter microstructure, adjusting for race/ethnicity**

| **White matter tracts** | **SES indicators (IVs)** | | | | | | | | |
| --- | --- | --- | --- | --- | --- | --- | --- | --- | --- |
|  | **Neighborhood disadvantage**  (higher = lower SES) | | | **Household income**  (higher = higher SES) | | | **Parental education**  (higher = higher SES) | | |
|  | ***β* (95% CI)** | ***p*-value**  (nominal) | ***p*-value**  (FDR) | ***β* (95% CI)** | ***p*-value**  (nominal) | ***p*-value**  (FDR) | ***β* (95% CI)** | ***p*-value**  (nominal) | ***p*-value**  (FDR) |
| **D) DTI-MD (DV)** | | | | | | | | | |
| Fx (right) | -0.002 (-0.030 to 0.027) | 0.92 | 0.93 | -0.024 (-0.055 to 0.007) | 0.12 | 0.26 | 0.008 (-0.021 to 0.037) | 0.59 | 0.76 |
| Fx (left) | -0.005 (-0.034 to 0.023) | 0.71 | 0.93 | -0.009 (-0.041 to 0.022) | 0.55 | 0.64 | 0.008 (-0.021 to 0.037) | 0.58 | 0.76 |
| CgC (right) | -0.005 (-0.034 to 0.024) | 0.74 | 0.93 | 0.005 (-0.026 to 0.037) | 0.74 | 0.77 | 0.004 (-0.025 to 0.033) | 0.79 | 0.87 |
| CgC (left) | -0.005 (-0.035 to 0.024) | 0.71 | 0.93 | 0.017 (-0.014 to 0.048) | 0.27 | 0.43 | -0.003 (-0.032 to 0.025) | 0.82 | 0.87 |
| CgH (right) | 0.002 (-0.027 to 0.032) | 0.87 | 0.93 | 0.011 (-0.020 to 0.042) | 0.50 | 0.62 | -0.015 (-0.043 to 0.014) | 0.32 | 0.59 |
| CgH (left) | 0.004 (-0.025 to 0.033) | 0.78 | 0.93 | 0.026 (-0.005 to 0.057) | 0.10 | 0.24 | -0.011 (-0.040 to 0.017) | 0.43 | 0.61 |
| CST (right) | 0.011 (-0.018 to 0.040) | 0.45 | 0.81 | 0.017 (-0.014 to 0.048) | 0.30 | 0.44 | -0.011 (-0.040 to 0.017) | 0.43 | 0.61 |
| CST (left) | 0.005 (-0.024 to 0.035) | 0.72 | 0.93 | 0.008 (-0.023 to 0.040) | 0.60 | 0.65 | -0.002 (-0.031 to 0.026) | 0.87 | 0.87 |
| ATR (right) | 0.001 (-0.028 to 0.031) | 0.93 | 0.93 | 0.033 (0.002 to 0.065) | 0.04 | 0.22 | -0.017 (-0.046 to 0.011) | 0.23 | 0.59 |
| ATR (left) | 0.005 (-0.024 to 0.034) | 0.73 | 0.93 | 0.021 (-0.010 to 0.052) | 0.19 | 0.34 | -0.003 (-0.031 to 0.026) | 0.84 | 0.87 |
| Unc (right) | 0.029 (0.000 to 0.058) | 0.05 | 0.54 | 0.029 (-0.003 to 0.061) | 0.07 | 0.22 | -0.013 (-0.042 to 0.016) | 0.37 | 0.59 |
| Unc (left) | 0.013 (-0.017 to 0.042) | 0.40 | 0.77 | 0.021 (-0.010 to 0.053) | 0.19 | 0.34 | -0.004 (-0.033 to 0.025) | 0.81 | 0.87 |
| ILF (right) | 0.026 (-0.002 to 0.055) | 0.07 | 0.54 | 0.015 (-0.016 to 0.046) | 0.35 | 0.47 | -0.019 (-0.047 to 0.009) | 0.19 | 0.59 |
| ILF (left) | 0.018 (-0.010 to 0.047) | 0.21 | 0.58 | 0.008 (-0.023 to 0.039) | 0.60 | 0.65 | -0.016 (-0.044 to 0.012) | 0.26 | 0.59 |
| IFOF (right) | 0.029 (0.000 to 0.057) | 0.05 | 0.54 | 0.034 (0.004 to 0.065) | 0.03 | 0.22 | -0.016 (-0.044 to 0.012) | 0.27 | 0.59 |
| IFOF (left) | 0.019 (-0.010 to 0.047) | 0.21 | 0.58 | 0.031 (0.000 to 0.061) | 0.05 | 0.22 | -0.013 (-0.041 to 0.016) | 0.38 | 0.59 |
| Fmaj | -0.002 (-0.031 to 0.027) | 0.90 | 0.93 | -0.002 (-0.033 to 0.029) | 0.90 | 0.90 | 0.031 (0.002 to 0.060) | 0.04 | 0.57 |
| Fmin | -0.003 (-0.033 to 0.027) | 0.86 | 0.93 | 0.010 (-0.022 to 0.042) | 0.54 | 0.64 | 0.031 (0.002 to 0.060) | 0.04 | 0.57 |
| CC | 0.008 (-0.021 to 0.037) | 0.59 | 0.93 | 0.014 (-0.017 to 0.045) | 0.38 | 0.49 | 0.020 (-0.009 to 0.049) | 0.17 | 0.59 |
| SLF (right) | 0.024 (-0.005 to 0.053) | 0.10 | 0.54 | 0.030 (-0.001 to 0.061) | 0.06 | 0.22 | -0.024 (-0.053 to 0.004) | 0.09 | 0.59 |
| SLF (left) | 0.017 (-0.012 to 0.046) | 0.26 | 0.63 | 0.028 (-0.003 to 0.059) | 0.08 | 0.22 | -0.015 (-0.043 to 0.014) | 0.32 | 0.59 |
| tSLF (right) | 0.020 (-0.009 to 0.048) | 0.18 | 0.58 | 0.035 (0.004 to 0.065) | 0.03 | 0.22 | -0.027 (-0.055 to 0.002) | 0.06 | 0.59 |
| tSLF (left) | 0.016 (-0.013 to 0.045) | 0.28 | 0.63 | 0.028 (-0.003 to 0.059) | 0.08 | 0.22 | -0.013 (-0.042 to 0.015) | 0.36 | 0.59 |
| pSLF (right) | 0.026 (-0.003 to 0.054) | 0.08 | 0.54 | 0.029 (-0.002 to 0.060) | 0.07 | 0.22 | -0.023 (-0.051 to 0.005) | 0.11 | 0.59 |
| pSLF (left) | 0.019 (-0.010 to 0.048) | 0.20 | 0.58 | 0.030 (-0.001 to 0.061) | 0.06 | 0.22 | -0.016 (-0.045 to 0.012) | 0.27 | 0.59 |
| SCS (right) | 0.024 (-0.005 to 0.052) | 0.10 | 0.54 | 0.027 (-0.004 to 0.058) | 0.09 | 0.23 | -0.019 (-0.047 to 0.009) | 0.19 | 0.59 |
| SCS (left) | 0.009 (-0.020 to 0.038) | 0.53 | 0.92 | 0.018 (-0.013 to 0.049) | 0.25 | 0.40 | -0.013 (-0.042 to 0.015) | 0.37 | 0.59 |
| SIFC (right) | 0.022 (-0.008 to 0.051) | 0.15 | 0.58 | 0.032 (0.001 to 0.063) | 0.04 | 0.22 | -0.017 (-0.046 to 0.012) | 0.24 | 0.59 |
| SIFC (left) | 0.005 (-0.025 to 0.034) | 0.76 | 0.93 | 0.015 (-0.016 to 0.046) | 0.35 | 0.47 | 0.006 (-0.023 to 0.035) | 0.67 | 0.80 |
| IFSFC (right) | 0.016 (-0.013 to 0.045) | 0.28 | 0.63 | 0.020 (-0.012 to 0.051) | 0.22 | 0.38 | -0.018 (-0.046 to 0.011) | 0.22 | 0.59 |
| IFSFC (left) | 0.014 (-0.016 to 0.043) | 0.36 | 0.74 | 0.024 (-0.007 to 0.056) | 0.13 | 0.26 | -0.007 (-0.036 to 0.022) | 0.64 | 0.80 |

*Note*. Linear mixed effects models included all three SES indicators simultaneously as independent variables (IVs) and RSI and DTI metrics in each of the white matter tracts as the dependent variable (DV). Models were adjusted for participant age, sex, PDS, ICV, mean head motion, and race/ethnicity, and were nested by family. More restricted results were observed here compared to models that did not covary for race/ethnicity, possibly as a reflection of the high confounding between SES and race/ethnicity (see **eTable 6** in the **Supplement**). Multiple comparison correction was conducted within each neuroimaging metric and by each SES indicator, giving 31 tests to correct for in each group of models. Effects that survived false discovery rate (FDR)-corrected *p*-value ≤ 0.05 were considered statistically significant and are highlighted. Estimates were standardized *β*’s with 95% confidence intervals (CIs). RSI, restriction spectrum imaging; RND, restricted normalized directional; RNI, restricted normalized isotropic; DTI, diffusion tensor imaging; FA, fractional anisotropy; MD, mean diffusivity. Full names of white matter tracts are shown in the main text.

**eTable 11. PCA loadings of RSI and DTI metrics that were significantly associated with SES**

**1) Results for tracts significantly associated with neighborhood disadvantage**

| **Metric** | **Principal component (PC)** | |
| --- | --- | --- |
|  | **PC1** | **PC2** |
| **A) RSI-RND** | | |
| Loadings |  |  |
| Fmaj | 0.32 | 0.95 |
| SLF (left) | 0.56 | -0.19 |
| tSLF (left) | 0.55 | -0.17 |
| pSLF (left) | 0.54 | -0.19 |
| Proportion of variance | 0.78 | 0.20 |
| Cumulative proportion | 0.78 | 0.97 |
| **B) RSI-RNI** | | |
| Loadings |  |  |
| Fx (right) | -0.34 | -0.52 |
| Fx (left) | -0.34 | -0.53 |
| CgH (right) | -0.33 | 0.48 |
| CgH (left) | -0.34 | 0.46 |
| CST (right) | -0.36 | 0.06 |
| CST (left) | -0.36 | 0.08 |
| ATR (right) | -0.38 | -0.02 |
| ATR (left) | -0.38 | -0.02 |
| Proportion of variance | 0.59 | 0.13 |
| Cumulative proportion | 0.59 | 0.72 |
| **C) DTI-FA** | | |
| Loadings |  |  |
| SLF (left) | -0.59 | -0.17 |
| tSLF (left) | -0.58 | -0.60 |
| pSLF (left) | -0.56 | 0.78 |
| Proportion of variance | 0.95 | 0.05 |
| Cumulative proportion | 0.95 | 1.00 |

**2) Results for tracts significantly associated with household income**

| **Metric** | **Principal component (PC)** | |
| --- | --- | --- |
|  | **PC1** | **PC2** |
| **A) RSI-RNI** | | |
| Loadings |  |  |
| CgC (right) | -0.17 | 0.04 |
| CgC (left) | -0.17 | 0.01 |
| CgH (right) | -0.16 | 0.16 |
| CgH (left) | -0.16 | 0.14 |
| CST (right) | -0.16 | -0.33 |
| CST (left) | -0.16 | -0.33 |
| ATR (right) | -0.19 | 0.13 |
| ATR (left) | -0.19 | 0.12 |
| Unc (right) | -0.19 | 0.28 |
| Unc (left) | -0.19 | 0.27 |
| ILF (right) | -0.19 | 0.12 |
| ILF (left) | -0.20 | 0.11 |
| IFOF (right) | -0.21 | 0.17 |
| IFOF (left) | -0.21 | 0.15 |
| Fmin | -0.19 | 0.23 |
| CC | -0.21 | 0.03 |
| SLF (right) | -0.20 | -0.21 |
| SLF (left) | -0.21 | -0.21 |
| tSLF (left) | -0.20 | -0.20 |
| pSLF (right) | -0.21 | -0.18 |
| pSLF (left) | -0.20 | -0.26 |
| SCS (left) | -0.21 | -0.23 |
| SIFC (right) | -0.20 | 0.22 |
| SIFC (left) | -0.20 | 0.19 |
| IFSFC (right) | -0.21 | -0.09 |
| IFSFC (left) | -0.21 | -0.07 |
| Proportion of variance | 0.66 | 0.06 |
| Cumulative proportion | 0.66 | 0.72 |

**3) Results for tracts significantly associated with parental education**

| **Metric** | **Principal component (PC)** | |
| --- | --- | --- |
|  | **PC1** | **PC2** |
| **A) RSI-RND** | | |
| Loadings |  |  |
| CST (right) | -0.22 | -0.67 |
| CST (left) | -0.22 | -0.67 |
| SLF (right) | -0.40 | 0.13 |
| SLF (left) | -0.40 | 0.13 |
| tSLF (right) | -0.38 | 0.14 |
| tSLF (left) | -0.39 | 0.14 |
| pSLF (right) | -0.39 | 0.13 |
| pSLF (left) | -0.38 | 0.11 |
| Proportion of variance | 0.70 | 0.17 |
| Cumulative proportion | 0.70 | 0.87 |
| **B) RSI-RNI** | | |
| Loadings |  |  |
| Fmaj | NA | NA |
| Proportion of variance | NA | NA |
| Cumulative proportion | NA | NA |
| **C) DTI-FA** | | |
| Loadings |  |  |
| Fx (left) | -0.12 | 0.21 |
| CST (right) | -0.23 | 0.50 |
| CST (left) | -0.24 | 0.56 |
| SLF (right) | -0.38 | -0.19 |
| SLF (left) | -0.38 | -0.17 |
| tSLF (right) | -0.36 | -0.18 |
| tSLF (left) | -0.37 | -0.17 |
| pSLF (right) | -0.37 | -0.18 |
| pSLF (left) | -0.36 | -0.16 |
| SCS (left) | -0.25 | 0.46 |
| Proportion of variance | 0.58 | 0.17 |
| Cumulative proportion | 0.58 | 0.75 |

*Note.* Principal component analyses (PCA) was conducted with the “psych” package using singular value decomposition^12^. In a randomly selected imputed dataset, the first principal component (i.e., PC1) had strong loadings from all relevant tracts and captured substantial variance (58% to 95%) in RSI or DTI metrics; these PCs would serve as white matter microstructure composite scores in indirect effects models. Some PCs would be reverse-coded to ensure consistent direction of associations with SES indicators (i.e., higher SES, higher RSI-RND, lower RSI-RNI, and higher DTI-FA). Because parental education was associated with RSI-RNI only in Fmaj, no PCA was applied there. RSI, restriction spectrum imaging; RND, restricted normalized directional; RNI, restricted normalized isotropic; DTI, diffusion tensor imaging; FA, fractional anisotropy; MD, mean diffusivity. Full names of white matter tracts are shown in the main text.

**eTable 12. Associations between SES and white matter microstructure PCs**

| **White matter microstructure principal components (PCs)** | **SES indicators (IVs)** | | | | | | **Sample size (*n*)** |
| --- | --- | --- | --- | --- | --- | --- | --- |
|  | **Neighborhood disadvantage (ND)**  (higher = lower SES) | | **Household income (HI)**  (higher = higher SES) | | **Parental education (PE)**  (higher = higher SES) | |  |
|  | ***β* (95% CI)** | ***p*-value**  (nominal) | ***β* (95% CI)** | ***p*-value**  (nominal) | ***β* (95% CI)** | ***p*-value**  (nominal) |  |
| **A) RSI-RND (DV)** | | | | | | | |
| PC1 – ND | **-0.055 (-0.081 to -0.028)** | **< 0.001** | 0.016 (-0.013 to 0.046) | 0.28 | 0.044 (0.017 to 0.071) | 0.002 | 8799 |
| PC2 – ND | 0.011 (-0.017 to 0.039) | 0.43 | -0.018 (-0.049 to 0.014) | 0.27 | 0.045 (0.016 to 0.074) | 0.002 | 8799 |
| PC1 – PE | 0.030 (0.003 to 0.056) | 0.03 | -0.009 (-0.039 to 0.021) | 0.56 | **-0.056 (-0.084 to -0.029)*** | **< 0.001** | 8787 |
| PC2 – PE | -0.048 (-0.075 to -0.021) | 0.001 | -0.010 (-0.041 to 0.020) | 0.50 | -0.019 (-0.047 to 0.010) | 0.20 | 8787 |
| **B) RSI-RNI (DV)** | | | | | | | |
| PC1 – ND | **-0.057 (-0.084 to -0.030)*** | **< 0.001** | 0.038 (0.008 to 0.069) | 0.01 | 0.006 (-0.022 to 0.035) | 0.65 | 8707 |
| PC2 – ND | 0.019 (-0.009 to 0.046) | 0.19 | -0.043 (-0.075 to -0.012) | 0.01 | 0.017 (-0.012 to 0.045) | 0.26 | 8707 |
| PC1 – HI | -0.006 (-0.033 to 0.021) | 0.67 | **0.054 (0.023 to 0.084)*** | **0.001** | 0.006 (-0.022 to 0.035) | 0.65 | 8658 |
| PC2 – HI | 0.008 (-0.020 to 0.036) | 0.57 | -0.017 (-0.048 to 0.014) | 0.28 | -0.015 (-0.044 to 0.014) | 0.30 | 8658 |
| Fmaj – PE | 0.002 (-0.025 to 0.030) | 0.88 | -0.005 (-0.036 to 0.025) | 0.73 | **-0.048 (-0.077 to -0.020)** | **0.001** | 8798 |
| **C) DTI-FA (DV)** | | | | | | | |
| PC1 – ND | **0.064 (0.037 to 0.092)*** | **< 0.001** | -0.015 (-0.046 to 0.016) | 0.34 | -0.051 (-0.079 to -0.022) | < 0.001 | 8835 |
| PC2 – ND | 0.019 (-0.009 to 0.046) | 0.18 | -0.024 (-0.055 to 0.007) | 0.12 | 0.033 (0.005 to 0.062) | 0.02 | 8835 |
| PC1 – PE | 0.042 (0.015 to 0.069) | 0.003 | -0.007 (-0.038 to 0.024) | 0.65 | **-0.059 (-0.087 to -0.030)*** | **< 0.001** | 8826 |
| PC2 – PE | 0.056 (0.029 to 0.083) | < 0.001 | -0.003 (-0.033 to 0.027) | 0.85 | 0.018 (-0.010 to 0.046) | 0.20 | 8826 |

*Note*. The first PCs (i.e., PC1) demonstrated strong associations to specific SES indicators consistent with individual tracts from which the PCs were derived (see bolded text). In comparison, the second PCs (i.e., PC2) did not demonstrate appreciable associations with SES. PC1s were therefore designated as white matter microstructure PCs used in indirect effects models. Associations here were examined with linear mixed effects models with all three SES indicators as simultaneous independent variables (IVs) and each of the PCs as dependent variable (DV). Covariates included participant age, sex, PDS, ICV, and mean head motion as fixed effects, and family as random effect. * denotes PCs that would be reverse-coded to ensure consistent direction of associations with SES indicators (i.e., higher SES, higher RSI-RND, lower RSI-RNI, and higher DTI-FA); original unreversed PCs are shown here for accurate data representation. Estimates were standardized *β*’s with 95% confidence intervals (CIs). Results were pooled across all imputed datasets. RSI, restriction spectrum imaging; RND, restricted normalized directional; RNI, restricted normalized isotropic; DTI, diffusion tensor imaging; FA, fractional anisotropy; MD, mean diffusivity. Full names of white matter tracts are shown in the main text.

**eTable 13. Associations between SES and obesity-related measures**

| **Measures (DV)** | **SES indicators (IVs)** | | | | | |
| --- | --- | --- | --- | --- | --- | --- |
|  | **Neighborhood disadvantage**  (higher = lower SES) | | **Household income**  (higher = higher SES) | | **Parental education**  (higher = higher SES) | |
|  | ***β* (95% CI)** | ***p*-value**  (FDR) | ***β* (95% CI)** | ***p*-value**  (FDR) | ***β* (95% CI)** | ***p*-value**  (FDR) |
| BMI | 0.103 (0.076 to 0.129) | < 0.001 | -0.067 (-0.097 to -0.037) | < 0.001 | -0.106 (-0.133 to -0.078) | < 0.001 |
| Waist circumference | 0.082 (0.057 to 0.108) | < 0.001 | -0.054 (-0.083 to -0.025) | < 0.001 | -0.093 (-0.120 to -0.065) | < 0.001 |
| BMI *z*-score | 0.081 (0.055 to 0.107) | < 0.001 | -0.062 (-0.091 to -0.033) | < 0.001 | -0.089 (-0.116 to -0.061) | < 0.001 |

*Note*. Linear mixed effects models included all three SES indicators simultaneously as independent variables (IVs) and RSI and DTI metrics in each of the obesity-related measures as the dependent variable (DV). Models were adjusted for participant age, sex, and PDS, and were nested by family. Multiple comparison was corrected for three tests. Effects that survived false discovery rate (FDR)-corrected *p*-value ≤ 0.05 were considered statistically significant. Estimates were standardized *β*’s with 95% confidence intervals (CIs). SES, socioeconomic status; BMI, body mass index

**eTable 14. Indirect effects of obesity-related measures on associations between SES and white matter microstructure**

| **Model** | | **Path a**  **(IV 🡪 mediator)** | | **Path b**  **(Mediator 🡪 DV,**  **controlling for IV)** | | **Path a × b**  **(Indirect effect)** | | | | **Path c’**  **(IV 🡪 DV,**  **controlling for mediator)** | | **Proportion mediated** |
| --- | --- | --- | --- | --- | --- | --- | --- | --- | --- | --- | --- | --- |
| **IV** | **DV** | ***β* (SE)** | ***p*-value**  (nominal) | ***β* (SE)** | ***p*-value**  (nominal) | **Estimate (SE)** | ***p*-value**  (nominal) | ***p*-value**  (FDR) | **95% CI** | ***β* (SE)** | ***p*-value**  (nominal) | ***%*** |
| ***BMI as the mediator*** | | | | | | | | | | | | |
| Neighborhood disadvantage | RSI-RND PC | 0.108 (0.012) | < 0.001 | -0.035 (0.011) | 0.001 | -0.004 (0.001) | 0.002 | **0.003** | [-0.006, -0.001] | -0.049 (0.013) | < 0.001 | 7.55 |
|  | RSI-RNI PC | 0.104 (0.012) | < 0.001 | 0.148 (0.011) | < 0.001 | 0.015 (0.002) | < 0.001 | **< 0.001** | [0.011, 0.020] | 0.042 (0.013) | 0.001 | 26.75 |
|  | DTI-FA PC | 0.107 (0.012) | < 0.001 | -0.001 (0.011) | 0.911 | 0.000 (0.001) | 0.908 | 0.908 | [-0.003, 0.002] | -0.065 (0.013) | < 0.001 | 0.21 |
| Household income | RSI-RNI PC | -0.062 (0.014) | < 0.001 | 0.124 (0.011) | < 0.001 | -0.008 (0.002) | < 0.001 | **< 0.001** | [-0.011, -0.004] | -0.049 (0.015) | 0.001 | 13.75 |
| Parental education | RSI-RND PC | -0.103 (0.013) | < 0.001 | -0.047 (0.011) | < 0.001 | 0.005 (0.001) | < 0.001 | **0.007** | [0.003, 0.008] | 0.053 (0.014) | < 0.001 | 8.44 |
|  | RSI-RNI Fmaj | -0.106 (0.013) | < 0.001 | 0.082 (0.011) | < 0.001 | -0.009 (0.002) | < 0.001 | **< 0.001** | [-0.012, -0.006] | -0.040 (0.014) | 0.005 | 17.98 |
|  | DTI-FA PC | -0.105 (0.013) | < 0.001 | -0.011 (0.011) | 0.340 | 0.001 (0.001) | 0.338 | 0.386 | [-0.001, 0.004] | 0.059 (0.014) | < 0.001 | 1.89 |
| ***Waist circumference as the mediator*** | | | | | | | | | | | | |
| Neighborhood disadvantage | RSI-RND PC | 0.087 (0.012) | < 0.001 | -0.031 (0.011) | 0.005 | -0.003 (0.001) | 0.009 | **0.013** | [-0.005, -0.001] | -0.050 (0.013) | < 0.001 | 5.07 |
|  | RSI-RNI PC | 0.083 (0.013) | < 0.001 | 0.133 (0.011) | < 0.001 | 0.011 (0.002) | < 0.001 | **< 0.001** | [0.008, 0.015] | 0.046 (0.013) | < 0.001 | 19.30 |
|  | DTI-FA PC | 0.086 (0.012) | < 0.001 | 0.007 (0.011) | 0.519 | 0.001 (0.001) | 0.526 | 0.581 | [-0.001, 0.003] | -0.065 (0.013) | < 0.001 | -0.96 |
| Household income | RSI-RNI PC | -0.048 (0.014) | 0.001 | 0.115 (0.011) | < 0.001 | -0.006 (0.002) | 0.001 | **0.002** | [-0.009, -0.002] | -0.051 (0.015) | 0.001 | 9.89 |
| Parental education | RSI-RND PC | -0.091 (0.013) | < 0.001 | -0.040 (0.011) | < 0.001 | 0.004 (0.001) | 0.001 | **0.002** | [0.002, 0.006] | 0.054 (0.013) | < 0.001 | 6.40 |
|  | RSI-RNI Fmaj | -0.094 (0.013) | < 0.001 | 0.079 (0.011) | < 0.001 | -0.007 (0.001) | < 0.001 | **< 0.001** | [-0.011, -0.005] | -0.041 (0.014) | 0.004 | 15.40 |
|  | DTI-FA PC | -0.094 (0.013) | < 0.001 | -0.003 (0.011) | 0.816 | 0.000 (0.001) | 0.812 | 0.853 | [-0.002, 0.002] | 0.060 (0.014) | < 0.001 | 0.41 |
| ***BMI z-score as the mediator*** | | | | | | | | | | | | |
| Neighborhood disadvantage | RSI-RND PC | 0.085 (0.013) | < 0.001 | -0.033 (0.011) | 0.002 | -0.003 (0.001) | 0.004 | **0.006** | [-0.005, -0.001] | -0.050 (0.013) | < 0.001 | 5.28 |
|  | RSI-RNI PC | 0.081 (0.013) | < 0.001 | 0.139 (0.011) | < 0.001 | 0.011 (0.002) | < 0.001 | **< 0.001** | [0.007, 0.015] | 0.046 (0.013) | < 0.001 | 19.64 |
|  | DTI-FA PC | 0.084 (0.013) | < 0.001 | -0.011 (0.011) | 0.340 | -0.001 (0.001) | 0.322 | 0.386 | [-0.003, 0.001] | -0.064 (0.013) | < 0.001 | 1.39 |
| Household income | RSI-RNI PC | -0.060 (0.014) | < 0.001 | 0.101 (0.011) | < 0.001 | -0.006 (0.002) | < 0.001 | **< 0.001** | [-0.009, -0.003] | -0.050 (0.015) | 0.001 | 10.89 |
| Parental education | RSI-RND PC | -0.088 (0.013) | < 0.001 | -0.050 (0.011) | < 0.001 | 0.004 (0.001) | < 0.001 | **< 0.001** | [0.002, 0.007] | 0.053 (0.013) | < 0.001 | 7.60 |
|  | RSI-RNI Fmaj | -0.090 (0.013) | < 0.001 | 0.075 (0.011) | < 0.001 | -0.007 (0.001) | < 0.001 | **< 0.001** | [-0.010, -0.004] | -0.041 (0.014) | 0.003 | 14.02 |
|  | DTI-FA PC | -0.090 (0.013) | < 0.001 | -0.023 (0.011) | 0.038 | 0.002 (0.001) | 0.045 | 0.059 | [0.000, 0.004] | 0.058 (0.014) | < 0.001 | 3.45 |

*Note*. Higher levels of obesity-related measures had indirect effects on the associations between lower socioeconomic status (SES) and lower RSI-RND and greater RSI-RNI. In each indirect effects model, one of the three SES indicators was the independent variable (IV), one obesity-related measure was the mediator, and each of the white matter microstructure PCs that were associated with the SES indicator (i.e., IV) was the dependent variable (DV). Models were adjusted for the other two SES indicators (so that independent indirect effects between SES indicators could be assessed), age, sex, PDS, ICV, and mean head motion. False discovery rate (FDR) correction for multiple comparison was applied to all 21 tested indirect effects; results that survived the FDR-corrected threshold of *p*-value ≤ 0.05 are highlighted. Estimates were standardized *β*’s with standard errors (SEs); 95% confidence intervals (CIs) were estimated using 20000 Monte Carlo simulations. Results were pooled across all imputed datasets. Refer to **eTable 12** in the **Supplement** for sample sizes in individual models. BMI, body mass index; RSI, restriction spectrum imaging; RND, restricted normalized directional; RNI, restricted normalized isotropic; DTI, diffusion tensor imaging; FA, fractional anisotropy; MD, mean diffusivity

**eTable 15. Associations between SES and cognitive performance**

| **Measures (DV)** | **SES indicators (IVs)** | | | | | | |
| --- | --- | --- | --- | --- | --- | --- | --- |
|  | **Neighborhood disadvantage**  (higher = lower SES) | | **Household income**  (higher = higher SES) | | **Parental education**  (higher = higher SES) | | |
|  | ***β* (95% CI)** | ***p*-value**  (nominal) | ***β* (95% CI)** | ***p*-value**  (nominal) | ***β* (95% CI)** | | ***p*-value**  (nominal) |
| Total cognition | -0.124 (-0.150 to -0.099) | < 0.001 | 0.142 (0.114 to 0.171) | < 0.001 | | 0.242 (0.215 to 0.268) | < 0.001 |
| Crystallized cognition | -0.110 (-0.136 to -0.085) | < 0.001 | 0.127 (0.098 to 0.156) | < 0.001 | | 0.261 (0.234 to 0.287) | < 0.001 |
| Fluid cognition | -0.099 (-0.125 to -0.072) | < 0.001 | 0.110 (0.080 to 0.140) | < 0.001 | | 0.139 (0.111 to 0.167) | < 0.001 |
| Picture vocabulary | -0.131 (-0.156 to -0.106) | < 0.001 | 0.132 (0.104 to 0.161) | < 0.001 | | 0.233 (0.207 to 0.260) | < 0.001 |
| Flanker inhibitory control | -0.055 (-0.082 to -0.027) | < 0.001 | 0.073 (0.042 to 0.103) | < 0.001 | | 0.063 (0.034 to 0.091) | < 0.001 |
| List sorting | -0.100 (-0.126 to -0.074) | < 0.001 | 0.113 (0.084 to 0.142) | < 0.001 | | 0.155 (0.128 to 0.182) | < 0.001 |
| Dimensional card sort | -0.079 (-0.106 to -0.053) | < 0.001 | 0.072 (0.042 to 0.102) | < 0.001 | | 0.086 (0.058 to 0.114) | < 0.001 |
| Pattern recognition | -0.052 (-0.079 to -0.025) | < 0.001 | 0.033 (0.002 to 0.063) | 0.04 | | 0.060 (0.032 to 0.089) | < 0.001 |
| Picture sequencing | -0.043 (-0.070 to -0.017) | 0.001 | 0.097 (0.066 to 0.127) | < 0.001 | | 0.087 (0.059 to 0.115) | < 0.001 |
| Oral reading | -0.060 (-0.087 to -0.034) | < 0.001 | 0.094 (0.064 to 0.124) | < 0.001 | | 0.220 (0.192 to 0.248) | < 0.001 |

*Note*. Higher total cognition composite score (as the dependent variable (DV)) on the NIH Toolbox Cognition Batteries was independently associated with higher socioeconomic status (SES) as indexed by each of the three SES indicators (as independent variables (IVs)). Individual task and crystallized/fluid cognition composite scores were similarly associated with SES in exploratory analyses. Estimates are standardized *β*’s with 95% confidence intervals (CIs). Linear mixed effects models were adjusted for participant age, sex, and PDS and were nested by family. Because we only had one *a* *priori* hypothesis concerning total cognition and analyses with other cognitive measures were exploratory, multiple comparison correction did not apply.

**eTable 16.** **Indirect effects of total cognition score on associations between SES and white matter microstructure**

| **Model** | | **Path a**  **(IV 🡪 mediator)** | | **Path b**  **(Mediator 🡪 DV,**  **controlling for IV)** | | **Path a × b**  **(Indirect effect)** | | | | **Path c’**  **(IV 🡪 DV,**  **controlling for mediator)** | | **Proportion mediated** |
| --- | --- | --- | --- | --- | --- | --- | --- | --- | --- | --- | --- | --- |
| **IV** | **DV** | ***β* (SE)** | ***p*-value**  (nominal) | ***β* (SE)** | ***p*-value**  (nominal) | **Estimate (SE)** | ***p*-value**  (nominal) | ***p*-value**  (FDR) | **95% CI** | ***β* (SE)** | ***p*-value**  (nominal) | ***%*** |
| ***Total cognition score as the mediator*** | | | | | | | | | | | | |
| Neighborhood disadvantage | RSI-RND PC  (*n* = 8498) | -0.126 (0.012) | < 0.001 | 0.095 (0.012) | < 0.001 | -0.012 (0.002) | < 0.001 | **< 0.001** | [-0.016, -0.009] | -0.039 (0.013) | 0.002 | 23.59 |
|  | DTI-FA PC  (*n* = 8533) | -0.126 (0.012) | < 0.001 | 0.082 (0.012) | < 0.001 | -0.010 (0.002) | < 0.001 | **< 0.001** | [-0.014, -0.007] | -0.054 (0.013) | < 0.001 | 16.04 |
| Parental education | RSI-RND PC  (*n* = 8489) | 0.238 (0.013) | < 0.001 | 0.080 (0.012) | < 0.001 | 0.019 (0.003) | < 0.001 | **< 0.001** | [0.013, 0.025] | 0.038 (0.014) | 0.006 | 33.34 |
|  | DTI-FA PC  (*n* = 8524) | 0.238 (0.013) | < 0.001 | 0.067 (0.012) | < 0.001 | 0.016 (0.003) | < 0.001 | **< 0.001** | [0.010, 0.022] | 0.042 (0.014) | 0.004 | 27.58 |

*Note*. Greater total cognition had indirect effects on the associations between higher socioeconomic status (SES) and greater RSI-RND and DTI-FA. In each indirect effects model, one of the three SES indicators was the independent variable (IV), total cognition score was the mediator, and each of the white matter microstructure PCs that were associated with the SES indicator (i.e., IV) was the dependent variable (DV). Models were adjusted for the other two SES indicators (so that independent indirect effects between SES indicators could be assessed), age, sex, PDS, ICV, and mean head motion. False discovery rate (FDR) correction for multiple comparison was applied to all 4 tested indirect effects; results that survived the FDR-corrected threshold of *p*-value ≤ 0.05 are highlighted. Estimates were standardized *β*’s with standard errors (SEs); 95% confidence intervals (CIs) were estimated using 20000 Monte Carlo simulations. Results were pooled across all imputed datasets. Sample sizes are noted here as both SES indicators and white matter microstructure PCs had missing values. RSI, restriction spectrum imaging; RND, restricted normalized directional; RNI, restricted normalized isotropic; DTI, diffusion tensor imaging; FA, fractional anisotropy; MD, mean diffusivity

**eTable 17. Fit indices for main indirect effects models.**

| **Model** | | **Path a × b**  **(Indirect effect)** | | |
| --- | --- | --- | --- | --- |
| **IV** | **DV** | **CFI** | **RMSEA** | **SRMR** |
| ***BMI as the mediator*** | | | | |
| Neighborhood disadvantage | RSI-RND PC | 0.974 | 0.061 | 0.010 |
|  | RSI-RNI PC | 0.968 | 0.065 | 0.010 |
|  | DTI-FA PC | 0.964 | 0.063 | 0.010 |
| Household income | RSI-RNI PC | 0.968 | 0.066 | 0.010 |
| Parental education | RSI-RND PC | 0.975 | 0.060 | 0.009 |
|  | RSI-RNI Fmaj | 0.962 | 0.064 | 0.010 |
|  | DTI-FA PC | 0.965 | 0.064 | 0.010 |
| ***Waist circumference as the mediator*** | | | | |
| Neighborhood disadvantage | RSI-RND PC | 0.962 | 0.073 | 0.011 |
|  | RSI-RNI PC | 0.954 | 0.076 | 0.011 |
|  | DTI-FA PC | 0.949 | 0.074 | 0.011 |
| Household income | RSI-RNI PC | 0.955 | 0.077 | 0.011 |
| Parental education | RSI-RND PC | 0.964 | 0.073 | 0.010 |
|  | RSI-RNI Fmaj | 0.945 | 0.076 | 0.011 |
|  | DTI-FA PC | 0.952 | 0.075 | 0.011 |
| ***BMI z-score as the mediator*** | | | | |
| Neighborhood disadvantage | RSI-RND PC | 0.953 | 0.080 | 0.011 |
|  | RSI-RNI PC | 0.946 | 0.081 | 0.011 |
|  | DTI-FA PC | 0.938 | 0.079 | 0.011 |
| Household income | RSI-RNI PC | 0.945 | 0.082 | 0.012 |
| Parental education | RSI-RND PC | 0.955 | 0.079 | 0.011 |
|  | RSI-RNI Fmaj | 0.934 | 0.081 | 0.011 |
|  | DTI-FA PC | 0.941 | 0.080 | 0.011 |
| ***Total cognition composite score as the mediator*** | | | | |
| Neighborhood disadvantage | RSI-RND PC | 0.967 | 0.079 | 0.012 |
|  | DTI-FA PC | 0.959 | 0.079 | 0.012 |
| Parental education | RSI-RND PC | 0.968 | 0.079 | 0.012 |
|  | DTI-FA PC | 0.961 | 0.079 | 0.012 |

*Note*. Model fit indices included comparative fit index (CFI), root mean square error of approximation (RMSEA), and standardized root mean square residual (SRMR). Models demonstrated good fit^13–15^ that are comparable to those in reported models^16^. See **eTable 14** and **16** in the **Supplement** for detailed indirect effects estimates

**eTable 18. Indirect effects of cognitive task scores on associations between SES and white matter microstructure**

| **Model** | | **Path a**  **(IV 🡪 mediator)** | | **Path b**  **(Mediator 🡪 DV,**  **controlling for IV)** | | **Path a × b**  **(Indirect effect)** | | **Path c’**  **(IV 🡪 DV,**  **controlling for mediator)** | | **Proportion mediated** |
| --- | --- | --- | --- | --- | --- | --- | --- | --- | --- | --- |
| **IV** | **DV** | ***β* (SE)** | ***p*-value**  (nominal) | ***β* (SE)** | ***p*-value**  (nominal) | **Estimate (SE)** | ***p*-value**  (nominal) | ***β* (SE)** | ***p*-value**  (nominal) | ***%*** |
| ***Crystallized cognition composite score as the mediator*** | | | | | | | | | | |
| Neighborhood disadvantage | RSI-RND PC | -0.099 (0.013) | < 0.001 | 0.055 (0.011) | < 0.001 | -0.005 (0.001) | < 0.001 | -0.044 (0.013) | 0.001 | 11.07 |
|  | DTI-FA PC | -0.098 (0.013) | < 0.001 | 0.051 (0.012) | < 0.001 | -0.005 (0.001) | < 0.001 | -0.058 (0.014) | < 0.001 | 8.01 |
| Parental education | RSI-RND PC | 0.246 (0.013) | < 0.001 | 0.040 (0.011) | < 0.001 | 0.010 (0.003) | 0.001 | 0.046 (0.014) | 0.001 | 17.40 |
|  | DTI-FA PC | 0.246 (0.013) | < 0.001 | 0.033 (0.012) | 0.005 | 0.008 (0.003) | 0.005 | 0.049 (0.014) | 0.001 | 14.32 |
| ***Fluid cognition composite score as the mediator*** | | | | | | | | | | |
| Neighborhood disadvantage | RSI-RND PC | -0.091 (0.013) | < 0.001 | 0.092 (0.011) | < 0.001 | -0.008 (0.002) | < 0.001 | -0.042 (0.013) | 0.002 | 16.73 |
|  | DTI-FA PC | -0.092 (0.013) | < 0.001 | 0.075 (0.011) | < 0.001 | -0.007 (0.001) | < 0.001 | -0.057 (0.014) | < 0.001 | 10.78 |
| Parental education | RSI-RND PC | 0.132 (0.014) | < 0.001 | 0.082 (0.011) | < 0.001 | 0.011 (0.002) | < 0.001 | 0.046 (0.014) | 0.001 | 19.14 |
|  | DTI-FA PC | 0.131 (0.014) | < 0.001 | 0.071 (0.011) | < 0.001 | 0.009 (0.002) | < 0.001 | 0.049 (0.014) | 0.001 | 16.02 |
| ***Picture vocabulary task score (assessing verbal knowledge) as the mediator*** | | | | | | | | | | |
| Neighborhood disadvantage | RSI-RND PC | -0.122 (0.013) | < 0.001 | 0.063 (0.011) | < 0.001 | -0.008 (0.002) | < 0.001 | -0.042 (0.013) | 0.001 | 15.35 |
|  | DTI-FA PC | -0.120 (0.013) | < 0.001 | 0.061 (0.012) | < 0.001 | -0.007 (0.002) | < 0.001 | -0.055 (0.014) | < 0.001 | 11.72 |
| Parental education | RSI-RND PC | 0.218 (0.013) | < 0.001 | 0.046 (0.011) | < 0.001 | 0.010 (0.003) | < 0.001 | 0.048 (0.014) | < 0.001 | 17.32 |
|  | DTI-FA PC | 0.218 (0.013) | < 0.001 | 0.047 (0.012) | < 0.001 | 0.010 (0.003) | < 0.001 | 0.049 (0.014) | 0.001 | 17.36 |
| ***Flanker inhibitory control task score (assessing attention and executive functioning) as the mediator*** | | | | | | | | | | |
| Neighborhood disadvantage | RSI-RND PC | -0.050 (0.014) | < 0.001 | 0.042 (0.010) | < 0.001 | -0.002 (0.001) | 0.006 | -0.049 (0.013) | < 0.001 | 4.20 |
|  | DTI-FA PC | -0.051 (0.014) | < 0.001 | 0.028 (0.011) | 0.007 | -0.001 (0.001) | 0.029 | -0.062 (0.014) | < 0.001 | 2.31 |
| Parental education | RSI-RND PC | 0.058 (0.014) | < 0.001 | 0.034 (0.010) | 0.001 | 0.002 (0.001) | 0.009 | 0.056 (0.013) | < 0.001 | 3.47 |
|  | DTI-FA PC | 0.059 (0.014) | < 0.001 | 0.023 (0.010) | 0.025 | 0.001 (0.001) | 0.049 | 0.058 (0.014) | < 0.001 | 2.32 |
| ***List sorting task score (assessing working memory) as the mediator*** | | | | | | | | | | |
| Neighborhood disadvantage | RSI-RND PC | -0.089 (0.013) | < 0.001 | 0.051 (0.011) | < 0.001 | -0.005 (0.001) | < 0.001 | -0.046 (0.013) | < 0.001 | 8.92 |
|  | DTI-FA PC | -0.088 (0.013) | < 0.001 | 0.050 (0.011) | < 0.001 | -0.004 (0.001) | < 0.001 | -0.059 (0.014) | < 0.001 | 7.02 |
| Parental education | RSI-RND PC | 0.146 (0.014) | < 0.001 | 0.049 (0.011) | < 0.001 | 0.007 (0.002) | < 0.001 | 0.050 (0.014) | < 0.001 | 12.57 |
|  | DTI-FA PC | 0.146 (0.014) | < 0.001 | 0.044 (0.011) | < 0.001 | 0.006 (0.002) | < 0.001 | 0.052 (0.014) | < 0.001 | 10.90 |
| ***Dimensional change card sort task score (assessing executive functioning) as the mediator*** | | | | | | | | | | |
| Neighborhood disadvantage | RSI-RND PC | -0.074 (0.013) | < 0.001 | 0.061 (0.010) | < 0.001 | -0.005 (0.001) | < 0.001 | -0.046 (0.013) | < 0.001 | 9.04 |
|  | DTI-FA PC | -0.075 (0.013) | < 0.001 | 0.051 (0.011) | < 0.001 | -0.004 (0.001) | < 0.001 | -0.059 (0.014) | < 0.001 | 6.07 |
| Parental education | RSI-RND PC | 0.081 (0.014) | < 0.001 | 0.057 (0.010) | < 0.001 | 0.005 (0.001) | < 0.001 | 0.053 (0.013) | < 0.001 | 8.03 |
|  | DTI-FA PC | 0.081 (0.014) | < 0.001 | 0.051 (0.011) | < 0.001 | 0.004 (0.001) | < 0.001 | 0.054 (0.014) | < 0.001 | 7.08 |
| ***Pattern comparison task score (assessing processing speed) as the mediator*** | | | | | | | | | | |
| Neighborhood disadvantage | RSI-RND PC | -0.048 (0.014) | < 0.001 | 0.072 (0.010) | < 0.001 | -0.003 (0.001) | 0.002 | -0.048 (0.013) | < 0.001 | 6.87 |
|  | DTI-FA PC | -0.049 (0.014) | < 0.001 | 0.048 (0.011) | < 0.001 | -0.002 (0.001) | 0.004 | -0.061 (0.014) | < 0.001 | 3.79 |
| Parental education | RSI-RND PC | 0.059 (0.014) | < 0.001 | 0.066 (0.010) | < 0.001 | 0.004 (0.001) | < 0.001 | 0.054 (0.013) | < 0.001 | 6.73 |
|  | DTI-FA PC | 0.057 (0.014) | < 0.001 | 0.047 (0.011) | < 0.001 | 0.003 (0.001) | 0.003 | 0.056 (0.014) | < 0.001 | 4.53 |
| ***Picture sequence task score (assessing episodic memory) as the mediator*** | | | | | | | | | | |
| Neighborhood disadvantage | RSI-RND PC | -0.040 (0.013) | 0.003 | 0.049 (0.010) | < 0.001 | -0.002 (0.001) | 0.011 | -0.049 (0.013) | < 0.001 | 3.94 |
|  | DTI-FA PC | -0.040 (0.013) | 0.003 | 0.047 (0.011) | < 0.001 | -0.002 (0.001) | 0.014 | -0.062 (0.014) | < 0.001 | 2.95 |
| Parental education | RSI-RND PC | 0.086 (0.014) | < 0.001 | 0.039 (0.010) | < 0.001 | 0.003 (0.001) | 0.001 | 0.055 (0.013) | < 0.001 | 5.78 |
|  | DTI-FA PC | 0.085 (0.014) | < 0.001 | 0.044 (0.011) | < 0.001 | 0.004 (0.001) | 0.001 | 0.056 (0.014) | < 0.001 | 6.24 |
| ***Oral reading task score (assessing reading ability) as the mediator*** | | | | | | | | | | |
| Neighborhood disadvantage | RSI-RND PC | -0.050 (0.013) | < 0.001 | 0.033 (0.011) | 0.002 | -0.002 (0.001) | 0.016 | -0.050 (0.013) | < 0.001 | 3.24 |
|  | DTI-FA PC | -0.050 (0.013) | < 0.001 | 0.029 (0.011) | 0.009 | -0.001 (0.001) | 0.032 | -0.063 (0.014) | < 0.001 | 2.22 |
| Parental education | RSI-RND PC | 0.211 (0.014) | < 0.001 | 0.022 (0.011) | 0.038 | 0.005 (0.002) | 0.039 | 0.054 (0.014) | < 0.001 | 8.01 |
|  | DTI-FA PC | 0.211 (0.014) | < 0.001 | 0.012 (0.011) | 0.289 | 0.002 (0.002) | 0.290 | 0.057 (0.014) | < 0.001 | 4.15 |

*Note*. In exploratory analyses, the independent associations between higher socioeconomic status (SES) indicators and greater RSI-RND and DTI-FA had indirect effects via cognitive performance that appeared to be general rather than domain-specific, as similar effects were observed across individual task and fluid/crystallized composite cognitive measures. In each indirect effects model, one of the three SES indicators was the independent variable (IV), cognition was the mediator, and each of the white matter microstructure PCs that were associated with the SES indicator (i.e., IV) was the dependent variable (DV). Models were adjusted for the other two SES indicators (so that independent indirect effects between SES indicators could be assessed), age, sex, PDS, ICV, and mean head motion. We did not apply multiple comparison correction due to the explorative nature of analyses. Estimates were standardized *β*’s with standard errors (SEs). Results were pooled across all imputed datasets. RSI, restriction spectrum imaging; RND, restricted normalized directional; RNI, restricted normalized isotropic; DTI, diffusion tensor imaging; FA, fractional anisotropy; MD, mean diffusivity

**eTable 19. Indirect effects of white matter microstructure on associations between SES and cognitive performance**

**1) Indirect effects on total cognition score**

| **Model** | | **Path a**  **(IV 🡪 mediator)** | | **Path b**  **(Mediator 🡪 DV,**  **controlling for IV)** | | **Path a × b**  **(Indirect effect)** | | | | **Path c’**  **(IV 🡪 DV,**  **controlling for mediator)** | | **Proportion mediated** |
| --- | --- | --- | --- | --- | --- | --- | --- | --- | --- | --- | --- | --- |
| **IV** | **Mediator** | ***β* (SE)** | ***p*-value**  (nominal) | ***β* (SE)** | ***p*-value**  (nominal) | **Estimate (SE)** | ***p*-value**  (nominal) | ***p*-value**  (FDR) | **95% CI** | ***β* (SE)** | ***p*-value**  (nominal) | ***%*** |
| ***Total cognition composite score as the DV*** | | | | | | | | | | | | |
| Neighborhood disadvantage | RSI-RND PC  (*n* = 8498) | -0.050 (0.013) | < 0.001 | 0.085 (0.010) | < 0.001 | -0.004 (0.001) | < 0.001 | **< 0.001** | [-0.007, -0.002] | -0.109 (0.012) | < 0.001 | 3.74 |
|  | DTI-FA PC  (*n* = 8533) | -0.064 (0.013) | < 0.001 | 0.068 (0.010) | < 0.001 | -0.004 (0.001) | < 0.001 | **< 0.001** | [-0.007, -0.002] | -0.109 (0.012) | < 0.001 | 3.82 |
| Parental education | RSI-RND PC  (*n* = 8489) | 0.056 (0.014) | < 0.001 | 0.071 (0.010) | < 0.001 | 0.004 (0.001) | < 0.001 | **< 0.001** | [0.002, 0.006] | 0.224 (0.013) | < 0.001 | 1.76 |
|  | DTI-FA PC  (*n* = 8524) | 0.057 (0.014) | < 0.001 | 0.057 (0.010) | < 0.001 | 0.003 (0.001) | 0.001 | **0.001** | [0.002, 0.005] | 0.224 (0.013) | < 0.001 | 1.42 |

**2) Indirect effects on individual task and fluid/crystallized cognition scores**

| **Model** | | **Path a**  **(IV 🡪 mediator)** | | **Path b**  **(Mediator 🡪 DV,**  **controlling for IV)** | | **Path a × b**  **(Indirect effect)** | | **Path c’**  **(IV 🡪 DV,**  **controlling for mediator)** | | **Proportion mediated** |
| --- | --- | --- | --- | --- | --- | --- | --- | --- | --- | --- |
| **IV** | **Mediator** | ***β* (SE)** | ***p*-value**  (nominal) | ***β* (SE)** | ***p*-value**  (nominal) | **Estimate (SE)** | ***p*-value**  (nominal) | ***β* (SE)** | ***p*-value**  (nominal) | ***%*** |
| ***Crystallized cognition composite score as the DV*** | | | | | | | | | | |
| Neighborhood disadvantage | RSI-RND PC | -0.049 (0.013) | < 0.001 | 0.050 (0.010) | < 0.001 | -0.002 (0.001) | 0.003 | -0.097 (0.012) | < 0.001 | 2.46 |
|  | DTI-FA PC | -0.063 (0.013) | < 0.001 | 0.043 (0.010) | < 0.001 | -0.003 (0.001) | 0.002 | -0.095 (0.012) | < 0.001 | 2.78 |
| Parental education | RSI-RND PC | 0.056 (0.014) | < 0.001 | 0.036 (0.010) | 0.001 | 0.002 (0.001) | 0.008 | 0.244 (0.013) | < 0.001 | 0.83 |
|  | DTI-FA PC | 0.057 (0.014) | < 0.001 | 0.028 (0.010) | 0.005 | 0.002 (0.001) | 0.022 | 0.245 (0.013) | < 0.001 | 0.65 |
| ***Fluid cognition composite score as the DV*** | | | | | | | | | | |
| Neighborhood disadvantage | RSI-RND PC | -0.050 (0.013) | < 0.001 | 0.092 (0.011) | < 0.001 | -0.005 (0.001) | < 0.001 | -0.086 (0.013) | < 0.001 | 5.09 |
|  | DTI-FA PC | -0.064 (0.013) | < 0.001 | 0.070 (0.011) | < 0.001 | -0.004 (0.001) | < 0.001 | -0.087 (0.013) | < 0.001 | 4.89 |
| Parental education | RSI-RND PC | 0.057 (0.014) | < 0.001 | 0.084 (0.011) | < 0.001 | 0.005 (0.001) | < 0.001 | 0.127 (0.014) | < 0.001 | 3.62 |
|  | DTI-FA PC | 0.058 (0.014) | < 0.001 | 0.068 (0.011) | < 0.001 | 0.004 (0.001) | 0.001 | 0.127 (0.014) | < 0.001 | 3.00 |
| ***Picture vocabulary task score (assessing verbal knowledge) as the DV*** | | | | | | | | | | |
| Neighborhood disadvantage | RSI-RND PC | -0.050 (0.013) | < 0.001 | 0.057 (0.010) | < 0.001 | -0.003 (0.001) | 0.001 | -0.119 (0.012) | < 0.001 | 2.36 |
|  | DTI-FA PC | -0.063 (0.013) | < 0.001 | 0.051 (0.010) | < 0.001 | -0.003 (0.001) | < 0.001 | -0.117 (0.012) | < 0.001 | 2.68 |
| Parental education | RSI-RND PC | 0.058 (0.014) | < 0.001 | 0.042 (0.010) | < 0.001 | 0.002 (0.001) | 0.003 | 0.216 (0.013) | < 0.001 | 1.13 |
|  | DTI-FA PC | 0.059 (0.014) | < 0.001 | 0.041 (0.010) | < 0.001 | 0.002 (0.001) | 0.003 | 0.216 (0.013) | < 0.001 | 1.10 |
| ***Flanker inhibitory control task score (assessing attention and executive functioning) as the DV*** | | | | | | | | | | |
| Neighborhood disadvantage | RSI-RND PC | -0.051 (0.013) | < 0.001 | 0.047 (0.011) | < 0.001 | -0.002 (0.001) | 0.005 | -0.048 (0.013) | < 0.001 | 4.73 |
|  | DTI-FA PC | -0.063 (0.013) | < 0.001 | 0.029 (0.011) | 0.008 | -0.002 (0.001) | 0.022 | -0.049 (0.013) | < 0.001 | 3.66 |
| Parental education | RSI-RND PC | 0.058 (0.014) | < 0.001 | 0.038 (0.011) | 0.001 | 0.002 (0.001) | 0.009 | 0.056 (0.014) | < 0.001 | 3.81 |
|  | DTI-FA PC | 0.059 (0.014) | < 0.001 | 0.025 (0.011) | 0.027 | 0.001 (0.001) | 0.051 | 0.057 (0.014) | < 0.001 | 2.50 |
| ***List sorting task score (assessing working memory) as the DV*** | | | | | | | | | | |
| Neighborhood disadvantage | RSI-RND PC | -0.051 (0.013) | < 0.001 | 0.051 (0.011) | < 0.001 | -0.003 (0.001) | 0.003 | -0.087 (0.013) | < 0.001 | 2.90 |
|  | DTI-FA PC | -0.064 (0.013) | < 0.001 | 0.047 (0.010) | < 0.001 | -0.003 (0.001) | 0.001 | -0.085 (0.013) | < 0.001 | 3.40 |
| Parental education | RSI-RND PC | 0.057 (0.014) | < 0.001 | 0.050 (0.011) | < 0.001 | 0.003 (0.001) | 0.002 | 0.143 (0.014) | < 0.001 | 1.95 |
|  | DTI-FA PC | 0.059 (0.014) | < 0.001 | 0.042 (0.011) | < 0.001 | 0.002 (0.001) | 0.004 | 0.143 (0.014) | < 0.001 | 1.69 |
| ***Dimensional change card sort task score (assessing executive functioning) as the DV*** | | | | | | | | | | |
| Neighborhood disadvantage | RSI-RND PC | -0.050 (0.013) | < 0.001 | 0.066 (0.011) | < 0.001 | -0.003 (0.001) | 0.001 | -0.071 (0.013) | < 0.001 | 4.45 |
|  | DTI-FA PC | -0.063 (0.013) | < 0.001 | 0.051 (0.011) | < 0.001 | -0.003 (0.001) | 0.001 | -0.072 (0.013) | < 0.001 | 4.28 |
| Parental education | RSI-RND PC | 0.057 (0.014) | < 0.001 | 0.062 (0.011) | < 0.001 | 0.004 (0.001) | 0.001 | 0.077 (0.014) | < 0.001 | 4.41 |
|  | DTI-FA PC | 0.058 (0.014) | < 0.001 | 0.052 (0.011) | < 0.001 | 0.003 (0.001) | 0.002 | 0.078 (0.014) | < 0.001 | 3.76 |
| ***Pattern comparison task score (assessing processing speed) as the DV*** | | | | | | | | | | |
| Neighborhood disadvantage | RSI-RND PC | -0.051 (0.013) | < 0.001 | 0.079 (0.011) | < 0.001 | -0.004 (0.001) | 0.001 | -0.044 (0.013) | 0.001 | 8.33 |
|  | DTI-FA PC | -0.063 (0.013) | < 0.001 | 0.049 (0.011) | < 0.001 | -0.003 (0.001) | 0.001 | -0.046 (0.013) | < 0.001 | 6.37 |
| Parental education | RSI-RND PC | 0.058 (0.014) | < 0.001 | 0.073 (0.011) | < 0.001 | 0.004 (0.001) | < 0.001 | 0.055 (0.014) | < 0.001 | 7.19 |
|  | DTI-FA PC | 0.059 (0.014) | < 0.001 | 0.048 (0.011) | < 0.001 | 0.003 (0.001) | 0.002 | 0.054 (0.014) | < 0.001 | 4.99 |
| ***Picture sequence task score (assessing episodic memory) as the DV*** | | | | | | | | | | |
| Neighborhood disadvantage | RSI-RND PC | -0.051 (0.013) | < 0.001 | 0.053 (0.011) | < 0.001 | -0.003 (0.001) | 0.003 | -0.038 (0.013) | 0.004 | 6.74 |
|  | DTI-FA PC | -0.063 (0.013) | < 0.001 | 0.047 (0.011) | < 0.001 | -0.003 (0.001) | 0.001 | -0.037 (0.013) | 0.005 | 7.50 |
| Parental education | RSI-RND PC | 0.059 (0.014) | < 0.001 | 0.043 (0.011) | < 0.001 | 0.003 (0.001) | 0.004 | 0.083 (0.014) | < 0.001 | 2.94 |
|  | DTI-FA PC | 0.060 (0.014) | < 0.001 | 0.045 (0.011) | < 0.001 | 0.003 (0.001) | 0.003 | 0.083 (0.014) | < 0.001 | 3.13 |
| ***Oral reading task score (assessing reading ability) as the DV*** | | | | | | | | | | |
| Neighborhood disadvantage | RSI-RND PC | -0.052 (0.013) | < 0.001 | 0.033 (0.011) | 0.002 | -0.002 (0.001) | 0.016 | -0.049 (0.013) | < 0.001 | 3.48 |
|  | DTI-FA PC | -0.065 (0.013) | < 0.001 | 0.027 (0.010) | 0.010 | -0.002 (0.001) | 0.026 | -0.048 (0.013) | < 0.001 | 3.55 |
| Parental education | RSI-RND PC | 0.059 (0.014) | < 0.001 | 0.023 (0.010) | 0.040 | 0.001 (0.001) | 0.066 | 0.210 (0.014) | < 0.001 | 0.62 |
|  | DTI-FA PC | 0.059 (0.014) | < 0.001 | 0.011 (0.011) | 0.294 | 0.001 (0.001) | 0.314 | 0.210 (0.014) | < 0.001 | 0.31 |

*Note*. The independent associations between higher socioeconomic status (SES) and better cognitive performance had indirect effects via greater RSI-RND and DTI-FA. Cognitive performance was primarily assessed using the total cognition composite score (**1**), supplemented by exploratory analyses on individual task scores (**2**). Effects again appear general rather than domain-specific. In each indirect effects model, one of the three SES indicators was the independent variable (IV), each of the white matter microstructure PCs that were associated with the SES indicator (i.e., IV) was the mediator, and cognition was the dependent variable (DV).. Models were adjusted for the other two SES indicators (so that independent indirect effects between SES indicators could be assessed), age, sex, PDS, ICV, and mean head motion. False discovery rate (FDR) correction for multiple comparison was applied to 4 tested indirect effects involving the total cognition composite score. Results that survived the FDR-corrected threshold of *p*-value ≤ 0.05 are highlighted. Estimates were standardized *β*’s with standard errors (SEs); 95% confidence intervals (CIs) were estimated using 20000 Monte Carlo simulations. Results were pooled across all imputed datasets. Sample sizes are noted here as both SES indicators and white matter microstructure PCs had missing values. RSI, restriction spectrum imaging; RND, restricted normalized directional; RNI, restricted normalized isotropic; DTI, diffusion tensor imaging; FA, fractional anisotropy; MD, mean diffusivity **eTable 20. Indirect effects model testing in random subsample with one participant per family**

**1) Indirect effects of obesity-related measures**

| **Model** | | **Path a**  **(IV 🡪 mediator)** | | **Path b**  **(Mediator 🡪 DV,**  **controlling for IV)** | | **Path a × b**  **(Indirect effect)** | | | | **Path c’**  **(IV 🡪 DV,**  **controlling for mediator)** | | **Proportion mediated** |
| --- | --- | --- | --- | --- | --- | --- | --- | --- | --- | --- | --- | --- |
| **IV** | **DV** | ***β* (SE)** | ***p*-value**  (nominal) | ***β* (SE)** | ***p*-value**  (nominal) | **Estimate (SE)** | ***p*-value**  (nominal) | ***p*-value**  (FDR) | **95% CI** | ***β* (SE)** | ***p*-value**  (nominal) | ***%*** |
| ***BMI as the mediator*** | | | | | | | | | | | | |
| Neighborhood disadvantage | RSI-RND PC  (*n* = 7598) | 0.107 (0.013) | < 0.001 | -0.030 (0.012) | 0.010 | -0.003 (0.001) | 0.010 | **0.017** | [-0.006, -0.001] | -0.051 (0.013) | < 0.001 | 5.95 |
|  | RSI-RNI PC  (*n* = 7526) | 0.104 (0.013) | < 0.001 | 0.149 (0.012) | < 0.001 | 0.015 (0.002) | < 0.001 | **< 0.001** | [0.011, 0.020] | 0.041 (0.014) | 0.003 | 27.52 |
|  | DTI-FA PC  (*n* = 7626) | 0.106 (0.013) | < 0.001 | 0.004 (0.012) | 0.768 | 0.000 (0.001) | 0.758 | 0.796 | [-0.002, 0.003] | -0.066 (0.014) | < 0.001 | -0.57 |
| Household income | RSI-RNI PC  (*n* = 7488) | -0.069 (0.015) | < 0.001 | 0.127 (0.012) | < 0.001 | -0.009 (0.002) | < 0.001 | **< 0.001** | [-0.013, -0.005] | -0.042 (0.016) | 0.008 | 17.41 |
| Parental education | RSI-RND PC  (*n* = 7592) | -0.099 (0.014) | < 0.001 | -0.042 (0.012) | < 0.001 | 0.004 (0.001) | 0.001 | **0.002** | [0.002, 0.007] | 0.053 (0.014) | < 0.001 | 7.26 |
|  | RSI-RNI Fmaj  (*n* = 7601) | -0.101 (0.014) | < 0.001 | 0.088 (0.012) | < 0.001 | -0.009 (0.002) | < 0.001 | **< 0.001** | [-0.013, -0.006] | -0.039 (0.015) | 0.010 | 18.82 |
|  | DTI-FA PC  (*n* = 7618) | -0.101 (0.014) | < 0.001 | -0.006 (0.012) | 0.602 | 0.001 (0.001) | 0.592 | 0.654 | [-0.002, 0.003] | 0.062 (0.015) | < 0.001 | 1.01 |
| ***Waist circumference as the mediator*** | | | | | | | | | | | | |
| Neighborhood disadvantage | RSI-RND PC  (*n* = 7598) | 0.088 (0.013) | < 0.001 | -0.029 (0.012) | 0.013 | -0.003 (0.001) | 0.020 | **0.028** | [-0.005, -0.001] | -0.051 (0.013) | < 0.001 | 4.72 |
|  | RSI-RNI PC  (*n* = 7526) | 0.084 (0.013) | < 0.001 | 0.135 (0.012) | < 0.001 | 0.011 (0.002) | < 0.001 | **< 0.001** | [0.008, 0.016] | 0.044 (0.014) | 0.001 | 20.52 |
|  | DTI-FA PC  (*n* = 7626) | 0.087 (0.013) | < 0.001 | 0.009 (0.012) | 0.441 | 0.001 (0.001) | 0.448 | 0.554 | [-0.001, 0.003] | -0.067 (0.013) | < 0.001 | -1.22 |
| Household income | RSI-RNI PC  (*n* = 7488) | -0.053 (0.015) | 0.001 | 0.118 (0.012) | < 0.001 | -0.006 (0.002) | 0.001 | **0.002** | [-0.010, -0.003] | -0.044 (0.016) | 0.005 | 12.51 |
| Parental education | RSI-RND PC  (*n* = 7592) | -0.089 (0.014) | < 0.001 | -0.039 (0.012) | 0.001 | 0.003 (0.001) | 0.002 | **0.004** | [0.001, 0.006] | 0.053 (0.014) | < 0.001 | 6.15 |
|  | RSI-RNI Fmaj  (*n* = 7601) | -0.091 (0.014) | < 0.001 | 0.083 (0.012) | < 0.001 | -0.008 (0.002) | < 0.001 | **< 0.001** | [-0.011, -0.005] | -0.040 (0.015) | 0.008 | 15.99 |
|  | DTI-FA PC  (*n* = 7618) | -0.091 (0.014) | < 0.001 | -0.002 (0.012) | 0.848 | 0.000 (0.001) | 0.844 | 0.844 | [-0.002, 0.002] | 0.062 (0.015) | < 0.001 | 0.33 |
| ***BMI z-score as the mediator*** | | | | | | | | | | | | |
| Neighborhood disadvantage | RSI-RND PC  (*n* = 7598) | 0.083 (0.013) | < 0.001 | -0.029 (0.012) | 0.013 | -0.002 (0.001) | 0.015 | **0.023** | [-0.005, -0.001] | -0.052 (0.013) | < 0.001 | 4.42 |
|  | RSI-RNI PC  (*n* = 7526) | 0.080 (0.014) | < 0.001 | 0.140 (0.012) | < 0.001 | 0.011 (0.002) | < 0.001 | **< 0.001** | [0.007, 0.016] | 0.044 (0.014) | 0.001 | 20.15 |
|  | DTI-FA PC  (*n* = 7626) | 0.083 (0.013) | < 0.001 | -0.006 (0.012) | 0.587 | -0.001 (0.001) | 0.566 | 0.654 | [-0.003, 0.002] | -0.065 (0.014) | < 0.001 | 0.82 |
| Household income | RSI-RNI PC  (*n* = 7488) | -0.067 (0.015) | < 0.001 | 0.104 (0.012) | < 0.001 | -0.007 (0.002) | < 0.001 | **< 0.001** | [-0.011, -0.004] | -0.043 (0.016) | 0.006 | 13.99 |
| Parental education | RSI-RND PC  (*n* = 7592) | -0.082 (0.014) | < 0.001 | -0.045 (0.012) | < 0.001 | 0.004 (0.001) | 0.001 | **0.002** | [0.002, 0.007] | 0.053 (0.014) | < 0.001 | 6.47 |
|  | RSI-RNI Fmaj  (*n* = 7601) | -0.082 (0.014) | < 0.001 | 0.082 (0.012) | < 0.001 | -0.007 (0.002) | < 0.001 | **< 0.001** | [-0.010, -0.004] | -0.041 (0.015) | 0.007 | 14.35 |
|  | DTI-FA PC  (*n* = 7618) | -0.083 (0.014) | < 0.001 | -0.019 (0.012) | 0.110 | 0.002 (0.001) | 0.119 | 0.157 | [0.000, 0.004] | 0.061 (0.015) | < 0.001 | 2.53 |

**2) Indirect effects of total cognition score**

| **Model** | | **Path a**  **(IV 🡪 mediator)** | | **Path b**  **(Mediator 🡪 DV,**  **controlling for IV)** | | **Path a × b**  **(Indirect effect)** | | | | **Path c’**  **(IV 🡪 DV,**  **controlling for mediator)** | | **Proportion mediated** |
| --- | --- | --- | --- | --- | --- | --- | --- | --- | --- | --- | --- | --- |
| **IV** | **DV** | ***β* (SE)** | ***p*-value**  (nominal) | ***β* (SE)** | ***p*-value**  (nominal) | **Estimate (SE)** | ***p*-value**  (nominal) | ***p*-value**  (FDR) | **95% CI** | ***β* (SE)** | ***p*-value**  (nominal) | ***%*** |
| ***Total cognition score as the mediator*** | | | | | | | | | | | | |
| Neighborhood disadvantage | RSI-RND PC  (*n* = 7327) | -0.129 (0.013) | < 0.001 | 0.098 (0.012) | < 0.001 | -0.013 (0.002) | < 0.001 | **< 0.001** | [-0.017, -0.009] | -0.039 (0.014) | 0.004 | 24.46 |
|  | DTI-FA PC  (*n* = 7354) | -0.129 (0.013) | < 0.001 | 0.086 (0.013) | < 0.001 | -0.011 (0.002) | < 0.001 | **< 0.001** | [-0.015, -0.008] | -0.054 (0.014) | < 0.001 | 17.07 |
| Parental education | RSI-RND PC  (*n* = 7324) | 0.242 (0.014) | < 0.001 | 0.083 (0.012) | < 0.001 | 0.020 (0.003) | < 0.001 | **< 0.001** | [0.014, 0.027] | 0.036 (0.015) | 0.017 | 36.23 |
|  | DTI-FA PC  (*n* = 7346) | 0.242 (0.014) | < 0.001 | 0.073 (0.013) | < 0.001 | 0.018 (0.003) | < 0.001 | **< 0.001** | [0.012, 0.024] | 0.042 (0.015) | 0.006 | 29.86 |

*Note*. Both (**1**) obesity-related measures and (**2**) total cognition had indirect effects on the associations between socioeconomic status (SES) and white matter microstructure. These findings here are all in line with those in main analyses (see **eTable 14** and **16** in the **Supplement**), ruling out confounding from family structure. In each indirect effects model, one of the three SES indicators was the independent variable (IV), one obesity-related measure or total cognition score was the mediator, and each of the white matter microstructure PCs that were associated with the SES indicator (i.e., IV) was the dependent variable (DV). Models were adjusted for the other two SES indicators (so that independent indirect effects between SES indicators could be assessed), age, sex, PDS, ICV, and mean head motion. False discovery rate (FDR) correction for multiple comparison was applied to each group of 21 tests (**1**) or 4 tests (**2**). Results that survived the FDR-corrected threshold of *p*-value ≤ 0.05 are highlighted. Estimates were standardized *β*’s with standard errors (SEs); 95% confidence intervals (CIs) were estimated using 20000 Monte Carlo simulations. Results were pooled across all imputed datasets. BMI, body mass index; RSI, restriction spectrum imaging; RND, restricted normalized directional; RNI, restricted normalized isotropic; DTI, diffusion tensor imaging; FA, fractional anisotropy; MD, mean diffusivity.
